# Triangulating population, individual, and genetic evidence for priority setting in structurally heterogeneous blocs: metabolic risk factors for stroke in the Shanghai Cooperation Organisation

**DOI:** 10.64898/2026.09.30.26364374

**Authors:** Xinyu Wang, Mu Li, Junzhao Zhang, Hongqi Zhang, Yongjie Ma

## Abstract

**BACKGROUND:** Regional stroke-prevention priorities usually rest on what a bloc’s members share. Where members span multiple Global Burden of Disease (GBD) super-regions and genetic ancestry groups, population, individual, and genetic evidence face different transportability limits, and earlier combined studies did not prespecify what counts as disagreement.

**METHODS:** We studied the Shanghai Cooperation Organisation, which spans four super-regions and five ancestry groups and is land-contiguous. For five metabolic risk factors we triangulated GBD 2023 attributable fractions of stroke DALYs in ten states, incident stroke among 7896 adults in the China Health and Retirement Longitudinal Study (2011–18; SHRs per 1 SD), and subtype-specific Mendelian randomisation in three ancestries. An explicit rule set, with one pre-submission amendment, classified convergence and divergence by whether a listed mechanism accounted for it.

**FINDINGS:** High systolic blood pressure converged: 53–71% of attributable DALYs, the strongest cohort exposure (SHR 1·37, 95% CI 1·26–1·48), and genetically supported in both ancestries. LDL cholesterol (GBD attribution restricted to ischaemic stroke; European genetic evidence for that subtype; no cohort association) and body-mass index (last in attribution but second in the cohort, with a 1·80-fold [1·48–2·18] gradient inside the 20–25 kg/m² minimum-risk window) showed structural discordance. The cohort lacked subtype data to establish LDL dilution. Fasting glucose and kidney dysfunction diverged without such a mechanism.

**INTERPRETATION:** Systolic blood pressure had the strongest cross-stream support. LDL cholesterol and body-mass index require qualified interpretation; fasting glucose and kidney dysfunction could not be ranked reliably. The framework identifies where priorities require scrutiny.

**FUNDING:** None.

## Introduction

Stroke is a leading cause of death and disability worldwide, its burden concentrated in low-income and middle-income countries and largely attributable to five modifiable metabolic exposures: high systolic blood pressure (SBP), high LDL cholesterol (LDL), high body-mass index (BMI), high fasting glucose (FG), and kidney dysfunction (KD).^1–3^ Regional analyses conventionally take an intergovernmental bloc as the unit and draw common priorities from what its member states share, which is sound where they resemble one another in the respects the evidence depends on. Two such respects can be counted, and we take them together as a bloc’s evidence-relevant structural heterogeneity: how many of the seven Global Burden of Disease super-regions—epidemiological, not purely geographic—it spans,^2^ and how many genetic ancestry groups it holds. Structural heterogeneity and the capacity to act on a common finding tend to be inversely distributed.

Where a bloc is heterogeneous in both respects, the difficulty is not the ecological fallacy alone but the simultaneous failure of three distinct extrapolations. Population-level attribution cannot be carried down to individuals. Individual-level estimates from one member state cannot be carried across to the others. Genetic estimates cannot be carried between ancestral lineages. Compounding these, each state’s attributable fraction in the Global Burden of Disease Study is computed from relative risks pooled across populations,^2^ so the population estimate already presumes a transportability that such heterogeneity renders uncertain.

Triangulation here is qualitative and decision-oriented across complementary estimands, not synthesis of a common causal effect, and does not assume source independence.^4^ Each failure maps to a matched component: population-to-individual extrapolation to the cohort, between-state epidemiological transportability to the population stream, and cross-ancestry transportability to genetic estimates. Other vulnerabilities are shared: attribution and cohort estimates rest on the same observational relative risks, so agreement between them is partly built in. Agreement counts only where the agreeing source was in a position to disagree, which makes divergence the more informative signal. Treating it as the object of inference requires stating in advance what counts as a divergence and which explanations are admissible: none may rest on a claim that no source can test.

A bloc on which this reasoning can be tested must satisfy a conjunction rather than a compromise: heterogeneous enough for the extrapolations to fail, and institutionally capable enough for a common finding to be acted on. Of thirteen intergovernmental organisations characterised against both criteria, the Shanghai Cooperation Organisation (SCO) satisfies them most clearly (Methods): its ten member states span four of the seven super-regions and five genetic ancestry groups, of which only European, east Asian, and south Asian have stroke genome-wide association data at scale.

Studies combining Global Burden of Disease estimates with Mendelian randomisation have been reported for a region and for a single country, the second adding survey data.^5,6^ Neither states in advance what would count as a disagreement or which explanations for one are admissible; where the sources did disagree, the divergence was attributed to pathways none of them could test, leaving it unfalsifiable and the conclusion undisturbed. We asked instead which of the five exposures no source contradicted when in a position to do so, triangulating attribution across the ten states, stroke incidence in a Chinese cohort, and genetic estimates in three ancestries, so that burden-based priorities reflect evidence strength and transportability.

## Methods

### Study design

We did a triangulation study integrating three sources of evidence with partly distinct biases—ecological GBD estimates, Chinese individual-level cohort data, and genetic evidence from multi-ancestry MR—to assess the contribution of five metabolic risk factors (high SBP, high BMI, high LDL, high FG, and KD) to stroke and its subtypes (IS, ICH, SAH) across the ten SCO member states, membership taken as at 2026. The three components are reported according to GATHER,^7^ STROBE,^8^ and STROBE-MR,^9^ respectively.

### The study bloc and its comparators

The SCO was fixed as the study bloc at the outset; the criteria below were specified afterwards, to locate it among the blocs that might have been chosen and to define the conditions under which the design transfers. Each is a count taken from an external source, and the comparison returns organisations exceeding the SCO on two of the three dimensions.

Thirteen intergovernmental organisations meeting a stated eligibility rule were characterised on three dimensions (appendix 1.1). Evidence-relevant structural heterogeneity was indexed by the number of GBD super-regions spanned and the number of genetic ancestry groups represented, these being the dimensions along which cause-of-death patterns, pooled relative risks, and GWAS reference panels fail to transport; developmental heterogeneity by the range of the Socio-demographic Index, reported for context only; and geographical coherence by the number of connected components formed by shared land borders (tables S1–S3, figure 1).

**Figure 1:**
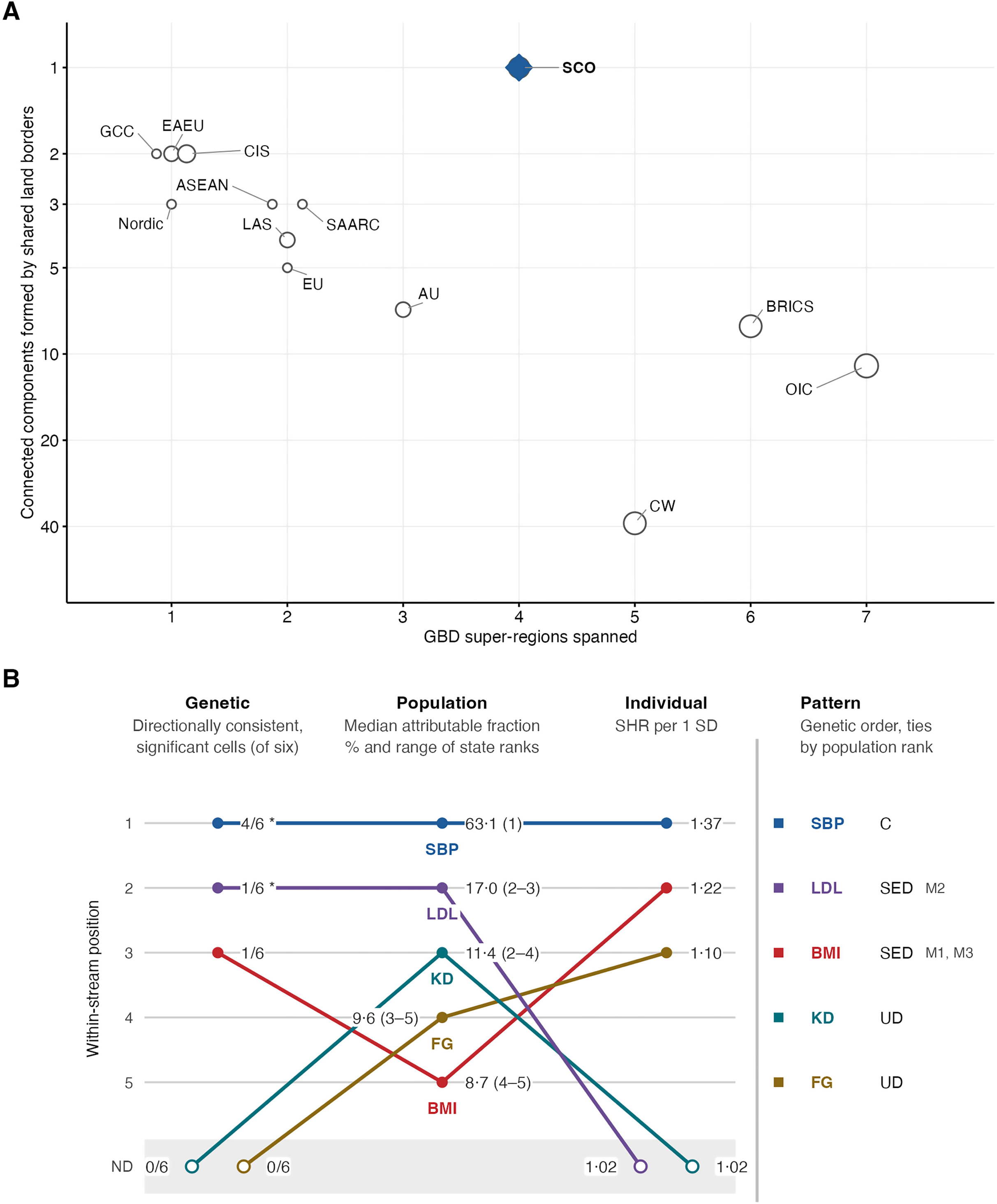
The study bloc among thirteen intergovernmental organisations, and the position of five metabolic risk factors within each evidence stream. (A) Structural heterogeneity and geographical coherence of thirteen intergovernmental organisations. The horizontal axis is the number of GBD super-regions spanned; the vertical axis is the number of connected components formed by treating member states as vertices and land borders between two member states of the same organisation as edges, on a reversed logarithmic scale, so that organisations higher in the panel are more geographically coherent. Borders with non-member states and maritime links create no edge. Point area is proportional to the number of genetic ancestry groups represented, groups matched by no current large-scale reference panel being counted alongside the rest. The Shanghai Cooperation Organisation (filled diamond) is the only organisation forming a single component. Points sharing identical coordinates are offset horizontally for legibility. The range of the Socio-demographic Index, the third dimension on which the organisations were characterised, is tabulated rather than plotted. Eligibility criteria and assignment rules are in appendix 1.1, the values for all thirteen organisations in tables S1–S2, and the assignment of individual countries to ancestry groups in table S3. (B) Within-stream ordering of the five exposures. The Genetic column reports the number of directionally consistent, FDR-significant ancestry-by-subtype cells out of six; Population reports the median attributable fraction (%) and range of within-state ranks; and Individual reports the mutually adjusted SHR per 1 SD. ND marks factors flagged not detected in that stream. Lines connect each factor across stream positions, and the adjacent numbers give the stream-specific values. The Pattern column reports the final classification: C=convergence; SED=structurally explicable divergence; UD=unexplained divergence; M1=threshold counterfactual against a continuous exposure–response; M2=outcome aggregation against subtype specificity; and M3=mediation through other measured exposures. Its display order is genetic order, with ties by population rank; this is only a display convention, not the genetic-stream tie-breaking rule, which is defined in appendix 3.1 by significant-cell count, MVMR persistence, and minimum FDR-corrected p-value. The LDL label reflects the revised qualitative M2 criterion: GBD supports an IS-specific attribution and European MR detects LDL–IS while no haemorrhagic estimate survives correction; C1 has a materially lower IS share than C2; the cohort does not resolve incident stroke by subtype, so the contribution of aggregation to the cohort null cannot be quantified (appendix 3.2). **Abbreviations** ASEAN=Association of Southeast Asian Nations. AU=African Union. BMI=body-mass index. C=convergence. CIS=Commonwealth of Independent States. CW=Commonwealth of Nations. DALY=disability-adjusted life-year. EAEU=Eurasian Economic Union. eGFR=estimated glomerular filtration rate. EU=European Union. FDR=false discovery rate. FG=fasting plasma glucose. GBD=Global Burden of Diseases, Injuries, and Risk Factors Study. GCC=Gulf Cooperation Council. KD=kidney dysfunction. LAS=League of Arab States. LDL=low-density lipoprotein cholesterol. ND=not detected. Nordic=Nordic Council. OIC=Organisation of Islamic Cooperation. SAARC=South Asian Association for Regional Cooperation. SBP=systolic blood pressure. SCO=Shanghai Cooperation Organisation. SD=standard deviation. SDI=Socio-demographic Index. SED=structurally explicable divergence. SHR=subdistribution hazard ratio. UD=unexplained divergence. BRICS is a proper name and not an initialism of its current membership.

Contiguity matters because proximity stands for shared exposure environment, health system, and population structure only where members are joined by land; it conditions the interpretability rather than the computability of the spatial analyses (appendix 1.4).

The SCO was the only organisation of the thirteen to form a single connected component. The Commonwealth, BRICS, and the Organisation of Islamic Cooperation matched or exceeded it on structural heterogeneity but fragmented into 39, eight, and 11 components; every other organisation forming fewer than five components spanned at most two super-regions. The CIS, whose membership overlaps the SCO’s most closely, spans a single super-region, so the SCO’s structural heterogeneity derives from its south and east Asian members rather than from its Eurasian core.

Four member states—Kazakhstan, Kyrgyzstan, Tajikistan, and Uzbekistan—belong to an ancestry group represented by no current large-scale reference panel, and Iran to one without stroke association data at the scale instrument selection requires, so the absence of genetic evidence for these five is a structural property of the bloc rather than a gap in study design (appendix 1.3).

Two features bear on the timeliness rather than the validity of the analysis: SCO health ministers convene through a standing mechanism, joined in February, 2026, by a centre for cooperation on metabolic disease, and a search of its published instruments to July 24, 2026, returned no bloc-wide target for non-communicable diseases or their risk factors, unlike the European Union, ASEAN, and the African Union (table S2), so evidence on priority ordering bears on an initial rather than a revised agenda.

### Data sources

For the ecological component we used national age-standardised GBD 2023 estimates of stroke incidence, mortality, and DALYs from 2001—the year the SCO was founded—to 2023, quantifying each risk factor’s contribution as the PAF of stroke DALYs (table S6).^2^ For the Chinese individual-level component we used CHARLS,^10^ a nationally representative cohort of adults aged 45 years or older across four waves (2011–2018), with venous blood sampled at waves 1 and 3. For the genetic component we did two-sample univariable and multivariable MR using public GWAS summary statistics in three ancestries, each mapped to the SCO members of predominantly that ancestry: European (Russia, Belarus), east Asian (China), and south Asian (India, Pakistan), the other five contributing no genetic evidence (tables S7–S8). Kidney function was represented by eGFR in the cohort and MR components,^11^ inversely to the GBD risk factor KD.

### GBD component

After describing stroke burden and its age and sex structure, we did three hierarchical clustering analyses of the ten states (Ward’s method on Euclidean distances; number of clusters by maximum mean silhouette width; the cluster containing China designated C1). We clustered states, in turn, on (1) the temporal dynamics of total-stroke burden; (2) the 2023 PAFs of the five risk factors; and (3) the isometric-log-ratio-transformed proportional composition of the three subtypes, fixing the cluster number to match the attribution clustering, to test whether the attribution pattern was underpinned by differences in stroke type. Geographical structure and cluster agreement were assessed by Mantel tests (primary), with join-count statistics and the adjusted Rand index as auxiliary measures, and stability by bootstrap resampling; secondary SDI, age-specific, and sex-specific analyses are in appendix 4. Because GBD estimates for data-sparse locations borrow strength within region and super-region, spatial concordance could reflect the estimation structure rather than exposure. We therefore repeated the Mantel test controlling for a hierarchy distance, tested the geographic gradient within the single super-region containing six member states, and repeated the primary test omitting each state in turn and omitting states whose cause-of-death data are rated two stars or fewer (appendix 2).

### CHARLS component

We included participants aged 45 years or older, alive and stroke-free at wave 1 (2011) with complete baseline metabolic measures; the five exposures were analysed per 1-SD increment. The outcome was incident stroke over waves 2–4 (to 2018), ascertained by self-report of a physician diagnosis without adjudication and without differentiation by subtype, with non-stroke death as a competing event and interval-censored event times imputed at wave midpoints (appendix 5.4). The primary analysis was a Fine-Gray subdistribution hazard model in three specifications—each exposure alone (Model 1), all five simultaneously (Model 2, main), and Model 2 plus antihypertensive, antidiabetic, and lipid-lowering medication (Model 3)—all adjusted for sociodemographic, behavioural, and clinical covariates (table S9).^12^ We examined nonlinearity with four-knot restricted cubic splines on the subdistribution hazard scale, referenced to values within the GBD TMREL window where one is defined on the same continuous scale, and tested proportional subdistribution hazards with exposure-by-log-time interactions. Prespecified subgroups (age <70 vs ≥70 years, sex, residence), three sensitivity analyses, and an auxiliary gradient-boosted (XGBoost–SHAP) importance analysis are described in appendix 5. CHARLS was approved by the Biomedical Ethics Review Committee of Peking University (IRB00001052-11015 for the household survey; IRB00001052-11014 for biospecimen collection), and all participants gave written informed consent.

### Multi-ancestry MR component

For each ancestry, the five exposures were instrumented from ancestry-matched GWAS (genome-wide significant, LD-clumped, per-variant F≥10; the threshold was relaxed and analyses treated as exploratory for south Asian ancestry), with IS, ICH, and SAH as the primary outcomes and total stroke as an ancestry-internal auxiliary outcome,^13,14^ because the available total-stroke phenotypes are not comparable across ancestries (appendix 6.1). The primary estimator was IVW MR, with MR-Egger and weighted-median sensitivity analyses and heterogeneity, pleiotropy, and influence diagnostics;^15,16^ MVMR (MVMR-IVW primary) estimated each exposure’s mutually adjusted direct effect,^17^ with conditional F and Q statistics (appendix 6.4). Within each outcome, IVW p values were FDR-corrected across the five exposures.^18^ Because no exposure study reports its units and instrument sets, linkage disequilibrium structure, and outcome definitions differ between ancestries, ancestries were compared by direction of effect and significance only. Where an estimate was compatible with the null, we report the smallest effect that analysis could have detected, so that a null is not read as evidence of absence (table S14).

### Evidence licensing

The three streams differ not only in their biases but in what each can be asked, so we state the conclusions each was taken to license before the rules by which they are compared.

The population stream is licensed to characterise differences between member states in attributed burden, conditional on the relative risks and the counterfactual exposure distributions that the Global Burden of Disease Study applies. It is not licensed to support a claim about individuals, and it cannot register the absence of an effect in a particular state: once a risk–outcome pair is included the attributable fraction is positive in every location by construction, so absence enters only as a global decision not to pair the two.

The individual stream is licensed to establish whether an exposure is associated with incident stroke across the range observed in the cohort, and the shape of that association; it is not licensed to supply an effect size for any other member state. For body-mass index the question is not how large the effect is anywhere in the bloc, but whether a theoretical-minimum-risk window falls inside an observed exposure range and whether risk varies within it. That is a property of the attribution procedure rather than of any particular population: a threshold counterfactual assigns no burden inside its window wherever it is applied, so an observed within-window gradient identifies a way attribution can miss variation relevant to the individual stream. It does not show that the cohort hazard ratio equals the relative risk used by GBD. What it licenses no inference about is set out in appendix 5.7.

The genetic stream is licensed to establish whether an effect operates on a given stroke subtype, within the ancestry in which it was estimated. A null in one ancestry does not transfer to another, nor are ancestries compared in magnitude (appendix 3.4); for the five member states without ancestry-matched stroke association data the stream is silent rather than negative.

### Evidence triangulation and decision rules

The original rules were specified before the streams were compared and framed on the streams’ structural properties rather than on this bloc’s results. After internal review but before submission, we amended M2 and report the amendment here: it requires a subtype-specific signal, aggregation of subtypes in the individual stream, and population-stream evidence of material heterogeneity in subtype composition, with genetic evidence supporting the subtype restriction. It does not require the unobserved CHARLS subtype proportion to be inferred.

Each stream was ordered internally by a criterion appropriate to what it is licensed to estimate: the population stream by median attributable fraction across the ten member states; the individual stream by subdistribution hazard ratio per 1 SD in the mutually adjusted model; and the genetic stream by the number of ancestry-by-subtype comparisons showing a directionally consistent, FDR-corrected association, ties broken by persistence in multivariable analysis and, where still tied, by a third criterion given in appendix 3.1. A factor was additionally flagged not detected in a stream when that stream’s estimate was compatible with the null after correction; the population stream has no such flag, for the reason given above. Positions are internal to each stream and carry no quantitative equivalence between streams.

Structurally explicable divergence required that a mechanism from a prespecified list (table S16) predict both the presence and the direction of the divergence observed. For M2, the revised precondition is that the subtype-specific signal is restricted to one subtype, the individual stream aggregates subtypes, and population-stream evidence supports material heterogeneity in subtype composition and genetic evidence supports the subtype restriction; it does not license a quantitative claim about the unobserved CHARLS subtype mix. Where no listed mechanism did so, the factor was classified as unexplained and no post-hoc explanation was offered. The derivation for each factor is given in table S17.

### Role of the funding source

This study received no funding.

## Results

### GBD component

#### Stroke burden, its temporal and demographic structure

Across the ten member states, the 2001–2023 trajectories of ASIR, ASMR, and ASDR formed three descriptive groups (mean silhouette width 0·22; figure 2, tables S19–S20): a large group with high-to-intermediate, steadily declining burden, a second with high but steeply declining burden, and Pakistan alone, persistently low and flat; incidence rose only in India. This temporal structure did not track geography, for total stroke or any subtype (Mantel and join-count both non-significant, except IS on the Mantel test; figures S1–S4, tables S20– S21). Burden rose log-linearly with age in both sexes, with small and inconsistent sex differences, and followed an inverted-U association with SDI (figures S5, S7–S8; tables S19 and S22).

**Figure 2:**
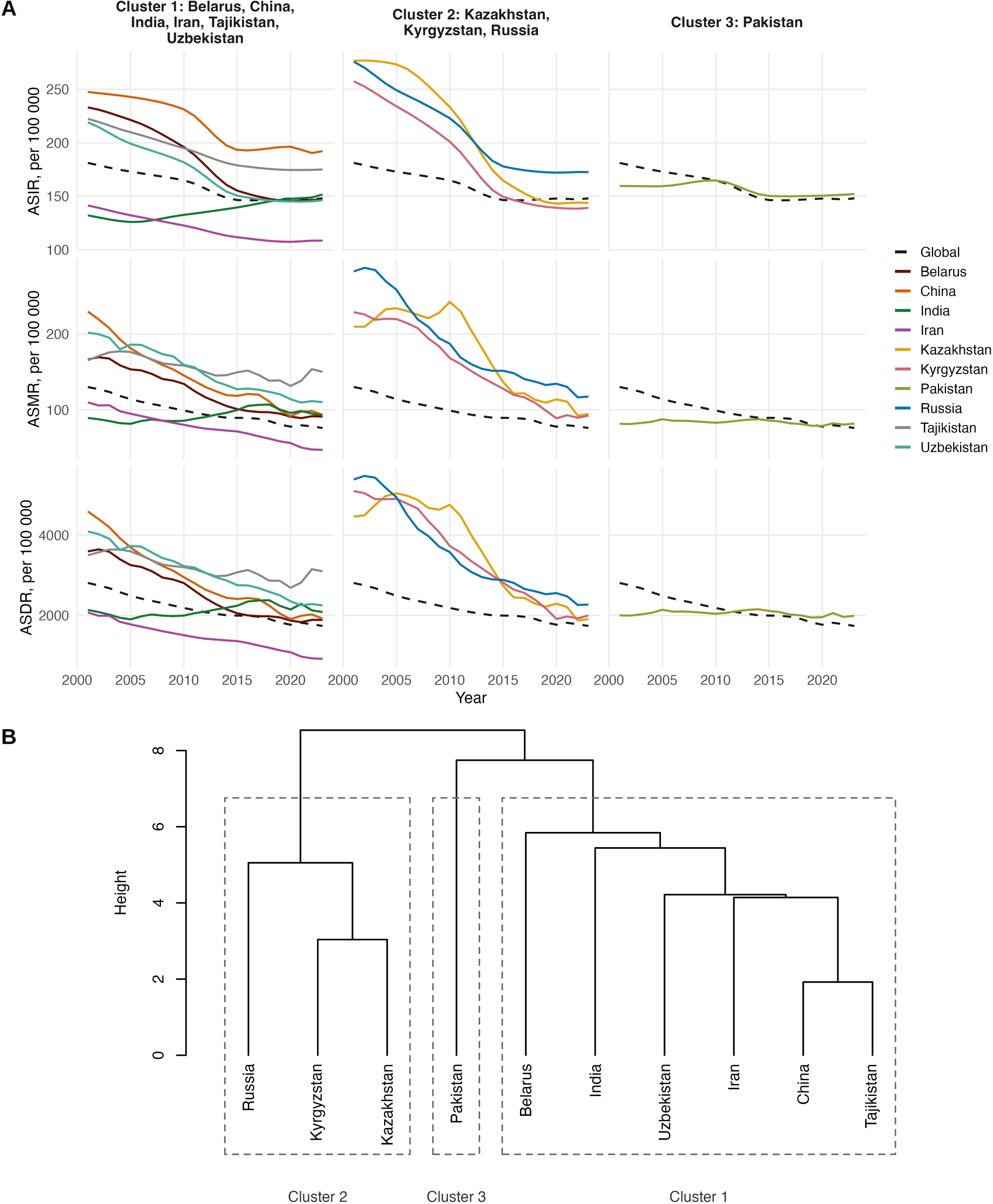
Temporal burden trajectories of total stroke in the ten SCO member states, 2001–2023. (A) Annual rates per 100 000 by trajectory cluster: rows are ASIR, ASMR, and ASDR and columns are the three clusters. Coloured lines are individual member states and the dashed black line is the global rate. The y-axis scale is shared across clusters within a row and differs between rows. (B) Dendrogram of the same clustering, in which the ten states were clustered by Ward’s linkage on five standardised temporal features of each rate series—linear slope, total percentage change, volatility, trend acceleration, and mean level—derived separately for each of the three rates, with the number of clusters chosen by maximum mean silhouette width. Dashed boxes delimit the three clusters, each labelled beneath, and height is the Ward merge distance; the tip order follows the clustering and not the left-to-right order of the columns in (A). Cluster 1 comprises Belarus, China, India, Iran, Tajikistan, and Uzbekistan; cluster 2 Kazakhstan, Kyrgyzstan, and Russia; and cluster 3 Pakistan alone. Cluster numbers are labels and do not order the clusters by burden. **Abbreviations** ASDR=age-standardised DALY rate. ASIR=age-standardised incidence rate. ASMR=age-standardised mortality rate. DALY=disability-adjusted life-year. SCO=Shanghai Cooperation Organisation.

#### Metabolic attribution clusters geographically

In 2023, high SBP dominated metabolic attribution in every state (about 53–71% of attributable stroke DALYs), ahead of high LDL (11–24%), KD (8·8–14·0%), high FG (8·1– 11·5%), and high BMI (1·6–11·0%; figure 3A, table 1). Clustering the states on these five PAFs gave two groups (mean silhouette width 0·34; bootstrap Jaccard 0·88 and 0·95; figure 3B, tables S24–S25): C1 (China, India, Pakistan), with generally lower attribution—most conspicuously for LDL (10·6–12·9%)—and C2 (the other seven), with roughly two-fold higher LDL attribution (17–24%, Tajikistan excepted). Unlike the temporal structure, this attribution pattern tracked geography closely (Mantel r=0·50, p=0·0063), forming a south-east (C1) versus north-west (C2) division; subtype-specific patterns were broadly similar (Mantel p≤0·03; figures S9–S10, table S26).

**Figure 3:**
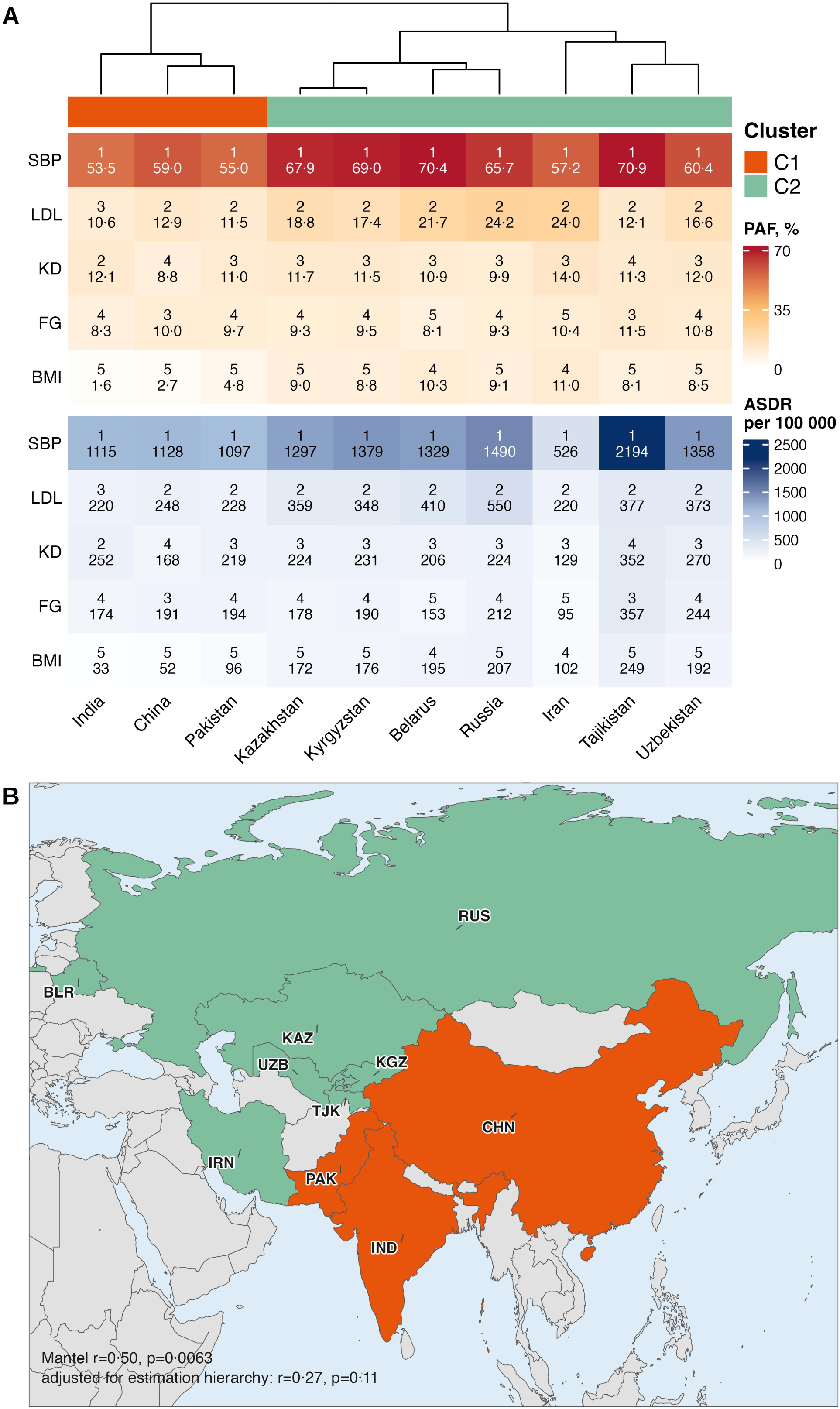
Metabolic risk attribution for stroke and its geographical clustering, 2023. (A) PAFs, as a percentage of stroke DALYs (upper), and attributable ASDR per 100 000 (lower) for the five metabolic risk factors, on independent colour scales; each cell gives the within-country rank of the risk factor above its value. Columns are ordered by the dendrogram above, obtained by Ward’s-linkage clustering of the five PAFs standardised across countries within each risk factor, and the colour bar beneath it denotes cluster membership. (B) The same clusters mapped, C1 being the cluster containing China; countries outside the SCO are grey. Annotated on the map are the Pearson Mantel correlation between PAF feature distances and great-circle distances separating national capitals, and beneath it the same correlation with the estimation hierarchy held fixed, each on 99 999 permutations. GBD estimates for locations with limited input data are informed by other locations in the same region and super-region, so member states sharing a position in that hierarchy may resemble one another for reasons internal to the modelling rather than to any real distribution of exposure. To make that alternative testable we defined a hierarchy distance between each pair of member states—0 where both belong to the same GBD region, 1 where they belong to the same super-region but different regions, and 2 otherwise—and held it fixed in a partial Mantel test. Attenuation therefore shows that geographical concordance is partly aligned with the estimation hierarchy; it does not separate concordance generated by borrowing from concordance with an independently constructed epidemiological typology, and appendix 2.2 sets out why that ambiguity is narrowed rather than removed (table S13). **Abbreviations** Member states are identified on the map by three-letter ISO country code—BLR=Belarus, CHN=China, IND=India, IRN=Iran, KAZ=Kazakhstan, KGZ=Kyrgyzstan, PAK=Pakistan, RUS=Russia, TJK=Tajikistan, UZB=Uzbekistan—and the same codes are used in supplementary figures. ASDR=age-standardised DALY rate. BMI=body-mass index. C1, C2=metabolic-attribution clusters 1 and 2, C1 being the cluster containing China. DALY=disability-adjusted life-year. FG=fasting plasma glucose. GBD=Global Burden of Diseases, Injuries, and Risk Factors Study. KD=kidney dysfunction. LDL=low-density lipoprotein cholesterol. PAF=population attributable fraction. SBP=systolic blood pressure. SCO=Shanghai Cooperation Organisation.

**Table 1:** Metabolic-risk population attributable fractions for stroke DALYs by member state and cluster, 2023. Values are the percentage of age-standardised total-stroke DALYs attributable to each risk factor, both sexes combined. Fractions are not additive across risk factors because the exposures overlap. C1 is the cluster containing China (figure 3). **Abbreviations** BMI=body-mass index. C1, C2=metabolic-attribution clusters 1 and 2. DALY=disability-adjusted life-year. FG=fasting plasma glucose. KD=kidney dysfunction. LDL=low-density lipoprotein cholesterol. SBP=systolic blood pressure.

| Cluster | Member state | SBP | LDL | KD | FG | BMI |
| --- | --- | --- | --- | --- | --- | --- |
| C1 | China | 59.0 | 12.9 | 8.8 | 10.0 | 2.7 |
| C1 | India | 53.5 | 10.6 | 12.1 | 8.3 | 1.6 |
| C1 | Pakistan | 55.0 | 11.5 | 11.0 | 9.7 | 4.8 |
| C2 | Belarus | 70.4 | 21.7 | 10.9 | 8.1 | 10.3 |
| C2 | Iran | 57.2 | 24.0 | 14.0 | 10.4 | 11.0 |
| C2 | Kazakhstan | 67.9 | 18.8 | 11.7 | 9.3 | 9.0 |
| C2 | Kyrgyzstan | 69.0 | 17.4 | 11.5 | 9.5 | 8.8 |
| C2 | Russia | 65.7 | 24.2 | 9.9 | 9.3 | 9.1 |
| C2 | Tajikistan | 70.9 | 12.1 | 11.3 | 11.5 | 8.1 |
| C2 | Uzbekistan | 60.4 | 16.6 | 12.0 | 10.8 | 8.5 |

Attribution profiles were aligned with the GBD estimation hierarchy at least as closely as with geographic distance in every outcome (r=0·57–0·76, all p≤0·0016), and adjustment for the hierarchy removed the geographic association for total stroke and subarachnoid haemorrhage while leaving it in place for ischaemic stroke and intracerebral haemorrhage, where it would not survive correction across the four outcomes (partial Mantel r=0·03–0·44). Within the single super-region containing six member states the geographic gradient persisted (r=0·58, p=0·01), and omitting each state in turn left the total-stroke association between 0·41 and 0·59; omitting the two states whose cause-of-death data are rated two stars or fewer raised it in all four outcomes (0·63–0·68). The pattern is therefore not an artefact of sparse input data, but geography and the estimation hierarchy cannot be separated with ten locations (appendix 2.2).

#### Subtype composition, and what it does and does not explain

The 2023 subtype composition had a distance structure associated with the attribution profile, although exact cluster membership agreed only modestly (Mantel r=0·68, p<0·0001; adjusted Rand index 0·29, p=0·17; figure 4, table S27). The ischaemia-predominant cluster had a high IS share of DALYs (58–75%); in the three C1 states the share was 39–42% and ICH exceeded IS. Because attribution for total stroke is the subtype-weighted sum of subtype-specific attribution, this correspondence is partly arithmetic, and the informative comparison is within subtype.

**Figure 4:**
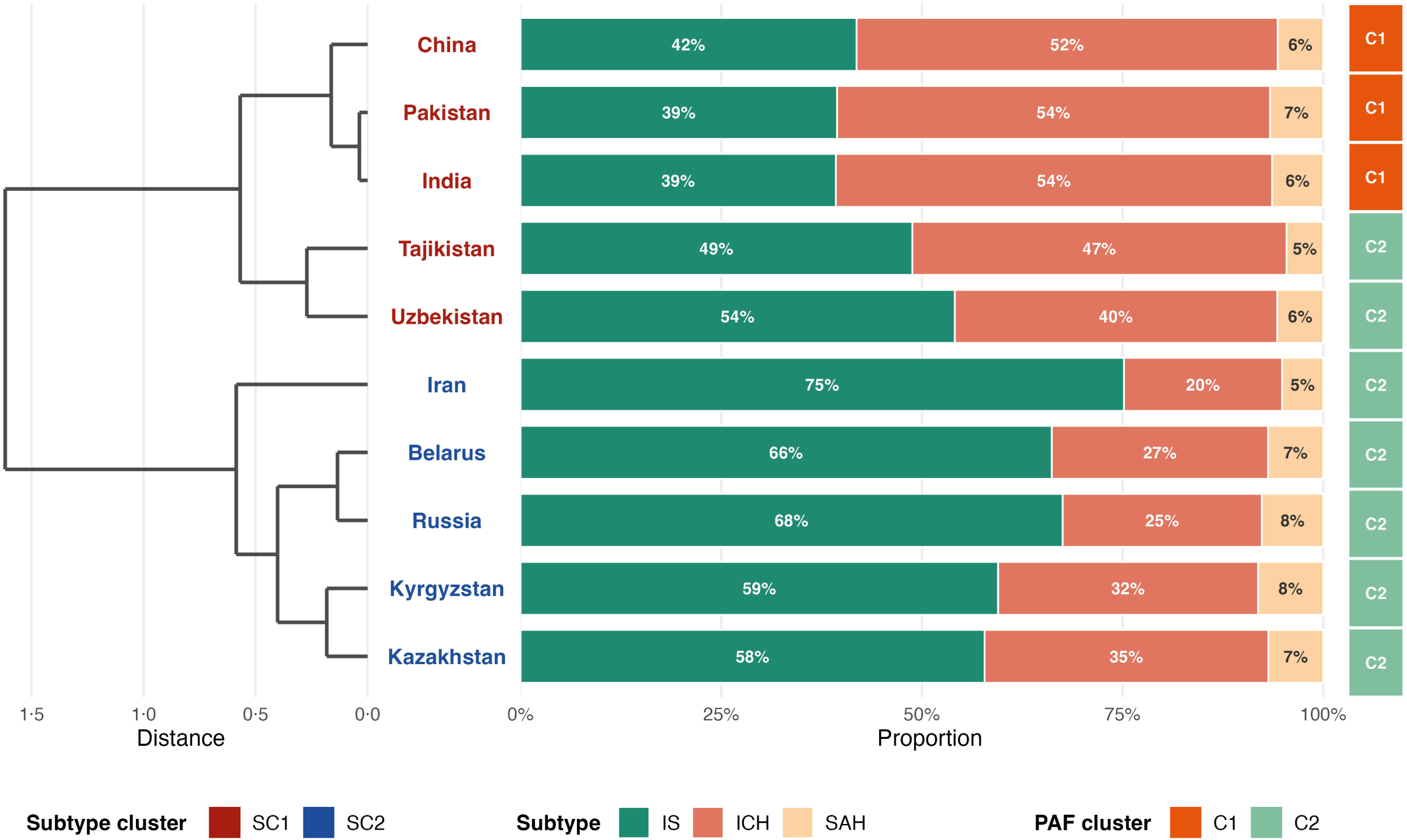
Stroke-subtype composition and its alignment with the metabolic-attribution clusters, 2023. Left, dendrogram of the ten member states clustered on the isometric-log-ratio-transformed proportions of IS, ICH, and SAH in ASDR (Ward’s linkage, with k fixed at 2 to match the attribution clustering); the axis is the Ward merge distance. Centre, stacked bars of the three subtype proportions, labelled where the proportion is at least 3%. Right, colour strip giving each state’s metabolic-attribution cluster (C1, C2; figure 3). Country-label colour denotes the subtype-composition cluster (SC1, SC2); SC1 and C1 are the clusters containing China. **Abbreviations** ASDR=age-standardised DALY rate. C1, C2=metabolic-attribution clusters 1 and 2. DALY=disability-adjusted life-year. ICH=intracerebral haemorrhage. IS=ischaemic stroke. SAH=subarachnoid haemorrhage. SC1, SC2=stroke subtype-composition clusters 1 and 2, SC1 and C1 both being the clusters containing China.

Made within subtype, it separates the two contrasts that look alike at the level of total stroke. LDL attribution to IS differed far less between the same clusters (C1 26·7–31·0% vs C2 24·8– 35·9%), so the two-fold contrast in total-stroke attribution is largely a composition effect: LDL is paired with ischaemic stroke alone, and the C1 states have the lower ischaemic share. BMI behaves differently. Its attribution was 2·5–5·4% in the C1 states against 9·5–11·7% elsewhere for IS, 0·9–4·3% against 6·6–9·5% for ICH, and 1·6–5·3% against 7·5–9·9% for SAH: the ranges do not overlap in any subtype, and the near three-fold contrast therefore cannot be attributed to composition. The difference in estimated BMI-attributable burden is not explained by subtype composition (figure S9).

### CHARLS component

#### Cohort and competing-risk outcomes

Of 25 586 CHARLS wave-1 participants, 8012 were stroke-free with complete baseline data and entered the analysis (figure S17); the 68·7% excluded were older and had more comorbidity, so selection may affect the associations, with no established direction of bias. Over a mean 6·3 years, 573 had an incident stroke and 682 died without stroke. Baseline differences were judged by the maximum pairwise SMD; the strongest gradients were with older age (1·05) and, among the exposures, higher SBP, higher FG, and lower eGFR. BMI was higher in those who had a stroke but lower in those who died (0·58), and LDL differed least (0·17; table 2, table S28).

**Table 2:** Baseline characteristics of the CHARLS analysis cohort by follow-up outcome. Values are mean ± SD or n (%); percentages are of non-missing values and missing counts refer to the overall cohort. Units are mmHg for SBP, kg/m² for BMI, mg/dL for FG and LDL, and mL/min per 1·73 m² for eGFR. The SMD column is the largest of the three pairwise standardised mean differences; *0·2–0·5, **0·5–0·8, ***>0·8. Full pairwise p values and SMDs are in table S28. Medication categories follow the CHARLS questionnaire wording. **Abbreviations** BMI=body-mass index. CHARLS=China Health and Retirement Longitudinal Study. eGFR=estimated glomerular filtration rate. FG=fasting plasma glucose. LDL=low-density lipoprotein cholesterol. SBP=systolic blood pressure. SD=standard deviation. SMD=standardised mean difference.

| Characteristic | Overall<br>(N=8012) | No event<br>(n=6757) | Stroke<br>(n=573) | Death<br>(n=682) | SMD | Missing, n |
| --- | --- | --- | --- | --- | --- | --- |
| **Metabolic exposures** |  |  |  |  |  |  |
| SBP | 129.21 ± 21.19 | 127.73 ± 20.34 | 138.02 ± 23.54 | 136.50 ± 23.89 | 0.468* | 0 |
| BMI | 23.48 ± 3.85 | 23.52 ± 3.80 | 24.48 ± 4.12 | 22.22 ± 3.69 | 0.578** | 0 |
| FG | 109.24 ± 32.57 | 108.12 ± 30.31 | 115.49 ± 39.02 | 115.00 ± 44.78 | 0.211* | 0 |
| LDL | 117.98 ± 34.32 | 118.13 ± 33.99 | 120.40 ± 35.31 | 114.48 ± 36.48 | 0.165 | 0 |
| eGFR | 95.78 ± 14.02 | 96.72 ± 13.52 | 93.88 ± 13.85 | 88.00 ± 16.24 | 0.583** | 0 |
| **Demographics** |  |  |  |  |  |  |
| Age (year) | 59.19 ± 9.29 | 58.15 ± 8.77 | 61.05 ± 8.44 | 67.99 ± 10.01 | 1.045*** | 0 |
| Sex |  |  |  |  | 0.326* | 0 |
| male | 3719 (46.4%) | 3025 (44.8%) | 279 (48.7%) | 415 (60.9%) |  |  |
| female | 4293 (53.6%) | 3732 (55.2%) | 294 (51.3%) | 267 (39.1%) |  |  |
| Marital Status |  |  |  |  | 0.409* | 0 |
| married/partnered | 7015 (87.6%) | 6028 (89.2%) | 484 (84.5%) | 503 (73.8%) |  |  |
| separated/divorced | 88 (1.1%) | 71 (1.1%) | 5 (0.9%) | 12 (1.8%) |  |  |
| widowed | 853 (10.6%) | 618 (9.1%) | 80 (14.0%) | 155 (22.7%) |  |  |
| never married | 56 (0.7%) | 40 (0.6%) | 4 (0.7%) | 12 (1.8%) |  |  |
| Education |  |  |  |  | 0.256* | 0 |
| less than lower secondary | 7257 (90·6%) | 6078 (90·0%) | 522 (91·1%) | 657 (96·3%) |  |  |
| upper secondary/vocational | 669 (8·3%) | 600 (8·9%) | 48 (8·4%) | 21 (3·1%) |  |  |
| tertiary | 86 (1·1%) | 79 (1·2%) | 3 (0·5%) | 4 (0·6%) |  |  |
| **Residence** |  |  |  |  | 0·164 | 0 |
| urban | 2826 (35·3%) | 2412 (35·7%) | 213 (37·2%) | 201 (29·5%) |  |  |
| rural | 5186 (64·7%) | 4345 (64·3%) | 360 (62·8%) | 481 (70·5%) |  |  |
| **Lifestyle and comorbidities** |  |  |  |  |  |  |
| **Smoking** |  |  |  |  | 0·339* | 1 |
| no | 4882 (60·9%) | 4241 (62·8%) | 327 (57·1%) | 314 (46·1%) |  |  |
| yes | 3129 (39·1%) | 2516 (37·2%) | 246 (42·9%) | 367 (53·9%) |  |  |
| **Drinking** |  |  |  |  | 0·179 | 5 |
| no | 4870 (60·8%) | 4181 (61·9%) | 327 (57·2%) | 362 (53·1%) |  |  |
| yes | 3137 (39·2%) | 2572 (38·1%) | 245 (42·8%) | 320 (46·9%) |  |  |
| **Heart disease history** |  |  |  |  | 0·277* | 29 |
| no | 7043 (88·2%) | 6022 (89·5%) | 455 (79·5%) | 566 (83·1%) |  |  |
| yes | 940 (11·8%) | 708 (10·5%) | 117 (20·5%) | 115 (16·9%) |  |  |
| **Depression** |  |  |  |  | 0·187 | 82 |
| no | 4945 (62·4%) | 4259 (63·5%) | 327 (57·7%) | 359 (54·4%) |  |  |
| yes | 2985 (37·6%) | 2444 (36·5%) | 240 (42·3%) | 301 (45·6%) |  |  |
| **Medication use** |  |  |  |  |  |  |
| **For high blood pressure** |  |  |  |  | 0·501** | 40 |
| No | 6450 (80·9%) | 5588 (83·1%) | 351 (61·4%) | 511 (75·3%) |  |  |
| Yes | 1522 (19·1%) | 1133 (16·9%) | 221 (38·6%) | 168 (24·7%) |  |  |
| **For diabetes** |  |  |  |  | 0·165 | 76 |
| No | 7626 (96·1%) | 6463 (96·6%) | 529 (93·0%) | 634 (93·5%) |  |  |
| Yes | 310 (3·9%) | 226 (3·4%) | 40 (7·0%) | 44 (6·5%) |  |  |
| **For dyslipidaemia** |  |  |  |  | 0·195 | 165 |
| No | 7452 (95·0%) | 6301 (95·3%) | 509 (90·4%) | 642 (95·4%) |  |  |
| Yes | 395 (5·0%) | 310 (4·7%) | 54 (9·6%) | 31 (4·6%) |  |  |
| **Follow-up time (year)** | 6·33 ± 1·42 | 6·71 ± 0·98 | 4·67 ± 1·44 | 3·93 ± 1·72 | 1·991*** | 0 |

#### Associations with incident stroke

Cumulative incidence stratified by clinical cut-offs diverged for SBP, BMI, and FG but not for eGFR or LDL (figure S18). In the mutually adjusted competing-risk model (Model 2; 7896 participants, 565 strokes), SBP was the strongest independent risk factor (SHR per 1 SD 1·37, 95% CI 1·26–1·48), followed by BMI (1·22, 1·14–1·30) and FG (1·10, 1·04–1·17); LDL (1·02, 0·94–1·11) and eGFR (1·02, 0·93–1·12) were not associated (figure 5). Estimates were materially unchanged in the single-exposure models and after further adjustment for medication, which attenuated all associations modestly but preserved their ranking, FG becoming marginal (figure 5, table S29).

**Figure 5:**
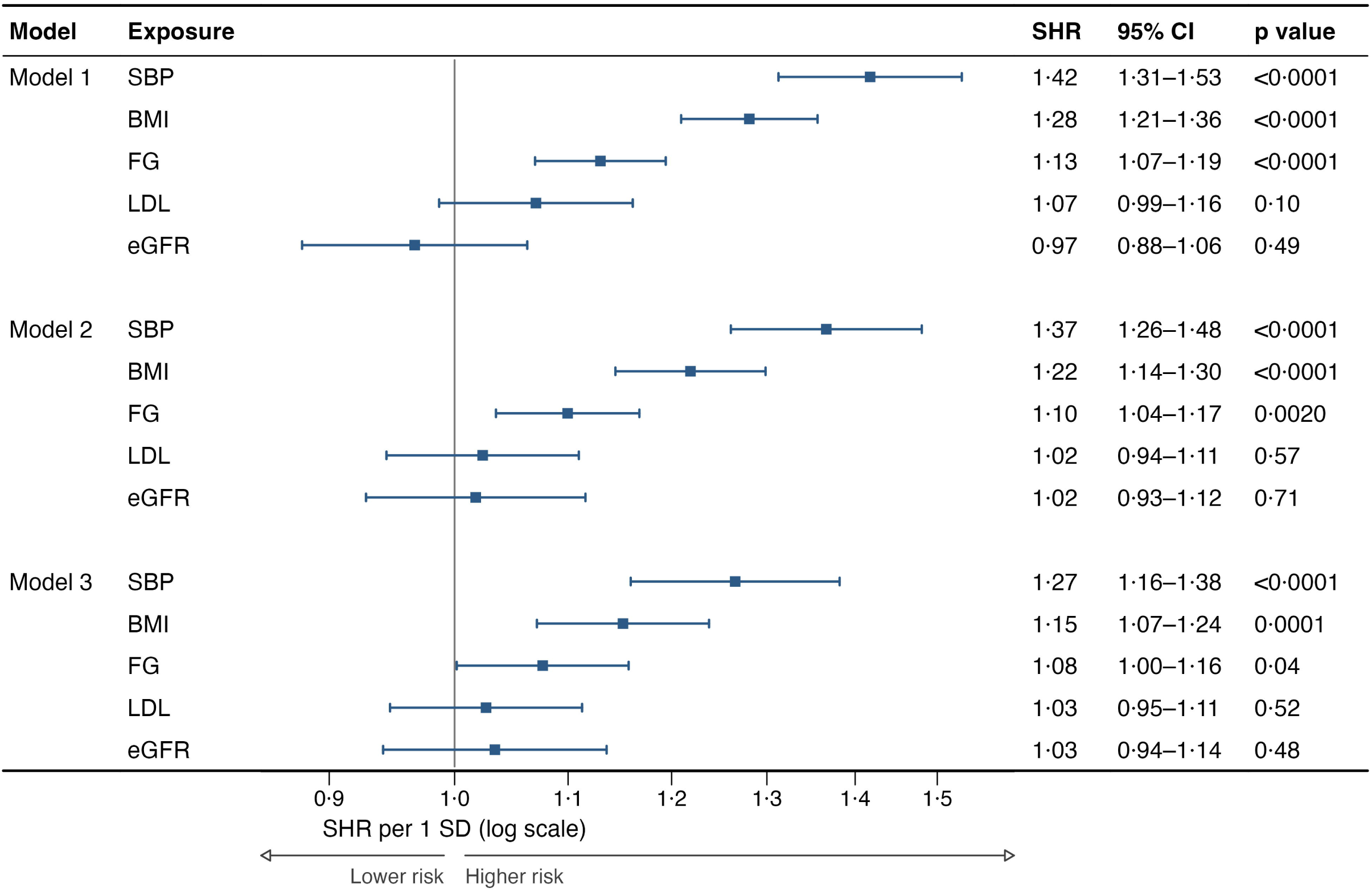
Associations of five metabolic risk factors with incident stroke in CHARLS. Fine-Gray subdistribution hazard ratios per 1 SD increment with 95% CIs, non-stroke death being the competing event; squares are point estimates, horizontal lines 95% CIs, and the vertical line the null. The x axis is on a log scale. Model 1 enters each exposure separately, Model 2 adjusts all five mutually and is the main model, and Model 3 adds antihypertensive, antidiabetic, and lipid-lowering medication; covariate sets are given in table S9 and full model output in table S29. Models 1 and 2 were fitted in 7896 participants (565 incident strokes) and Model 3, which additionally requires medication data, in 7710 (556). The five exposures were prespecified and mutually adjusted, so p values are not corrected for multiplicity. **Abbreviations** BMI=body-mass index. CHARLS=China Health and Retirement Longitudinal Study. CI=confidence interval. eGFR=estimated glomerular filtration rate. FG=fasting plasma glucose. LDL=low-density lipoprotein cholesterol. SBP=systolic blood pressure. SD=standard deviation. SHR=subdistribution hazard ratio.

#### Nonlinearity and time-dependence

Exposure–response curves fitted on the subdistribution hazard scale departed significantly from linearity for FG and BMI (nonlinearity p=0·0037 and p=0·0022; FDR-corrected p=0·0093 for both), but not for SBP, LDL, or eGFR (FDR-corrected p≥0·13; figure S19, tables S30–S31). Risk rose steeply across the lower range of both and then flattened, above about 110 mg/dL for FG and 26 kg/m² for BMI; the 20–25 kg/m² gradient alone corresponded to a 1·80-fold difference in subdistribution hazard (1·48–2·18). SBP and BMI also violated the proportional-subdistribution-hazards assumption (exposure-by-log-time interaction p<0·0001 and p=0·0001; table S32), their subdistribution hazards more than halving over follow-up without reversing direction; the SHRs above are therefore averages over the seven years. FG, LDL, and eGFR met the assumption.

#### Effect modification and robustness

Age was the strongest modifier (table S33): the SBP association was attenuated at 70 years or older (1·10 [0·89–1·36] vs 1·43 [1·31–1·55]; p=0·0047), and the eGFR association, null in both strata, differed in direction between them (p=0·02), a hypothesis-generating contrast given the null pooled estimate. Sex modified the SBP and BMI associations, both stronger in men (p=0·04 and 0·03), and residence modified none. Associations were reproduced by cause-specific models and, for SBP and BMI, by exposures averaged over two waves; FG was the least robust, losing significance under the averaged exposure. Results were unchanged with creatinine–cystatin C eGFR, and a gradient-boosted model reproduced the ordering of the three leading exposures and the nonlinear BMI and FG shapes (tables S34– S36, figure S20).

### Multi-ancestry MR component

#### European ancestry

In European-ancestry two-sample MR, genetically predicted SBP was associated with IS (OR 1·047 [95% CI 1·033–1·061]; FDR-corrected p<0·0001) but not, after correction, with either haemorrhagic subtype. LDL was associated with IS specifically (1·093 [1·034–1·155]; FDR p=0·0042) and not with ICH or SAH, and BMI showed a weak association with IS (1·030 [1·008–1·054]; FDR p=0·01); FG and eGFR were null across all subtypes (figure 6, table S38). In MVMR the LDL–IS effect persisted (1·091 [1·018–1·170]; conditional F=35·6) whereas the BMI–IS effect attenuated to the null (1·002 [0·974–1·031]); SBP remained associated with all three subtypes (table S40).

**Figure 6:**
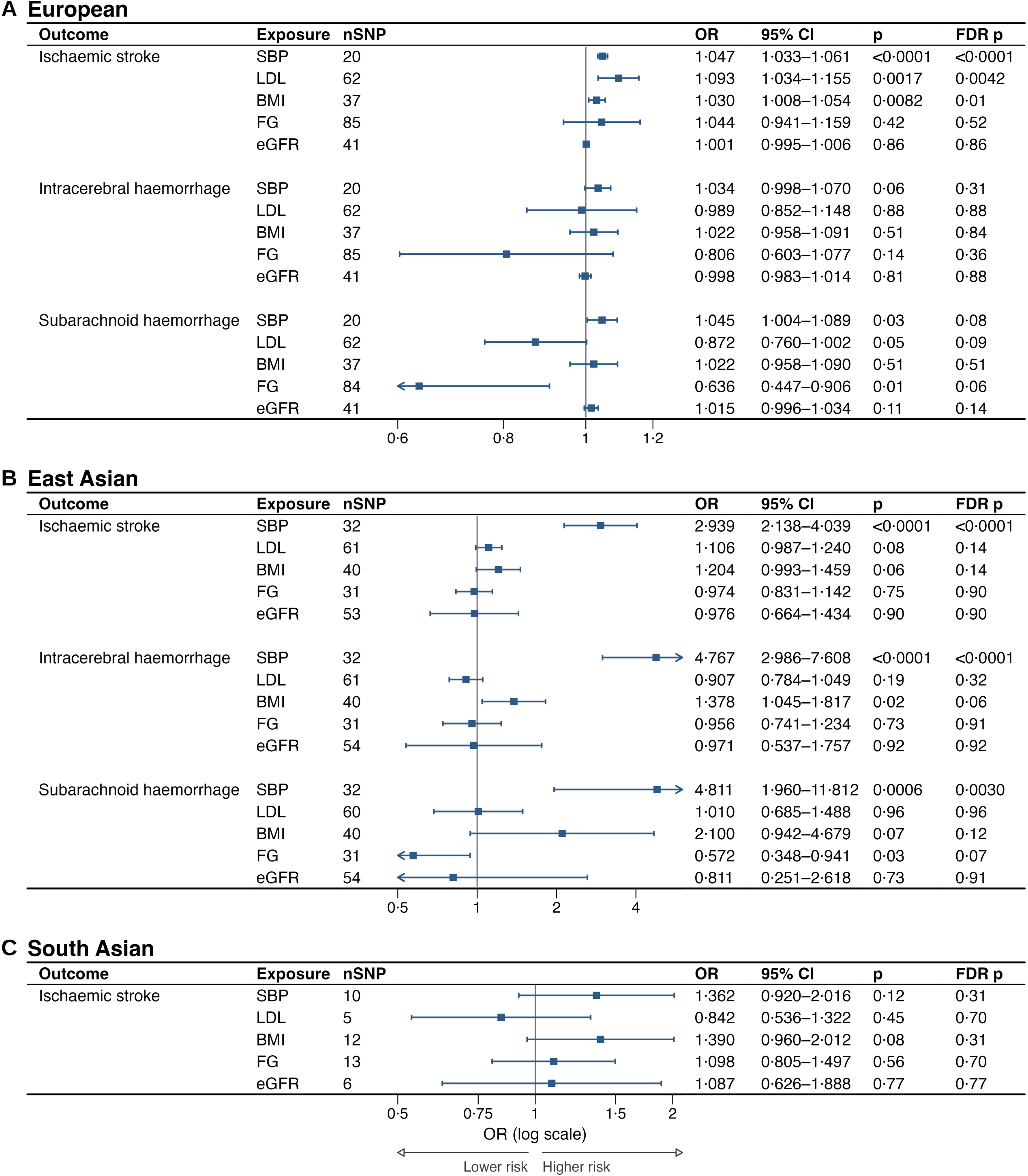
Genetically predicted metabolic exposures and stroke subtypes in three ancestries. Univariable inverse-variance-weighted estimates from two-sample Mendelian randomisation, shown for (A) European, (B) east Asian, and (C) south Asian ancestry. Squares are ORs per genetically predicted unit of exposure with 95% CIs; the vertical line marks the null and the x axis is on a log scale. Confidence intervals extending beyond the axis are truncated and marked with an arrowhead, their full limits being given in the adjacent column. The scale differs between panels because exposure units differ between ancestries, so estimates are comparable in direction and significance but not in magnitude. FDR correction is across the five exposures within each outcome. nSNP is the number of variants retained after harmonisation with each outcome; the instruments selected for each exposure before harmonisation are given in table S37. Full estimates are in table S38. South Asian analyses were exploratory and limited to ischaemic stroke, no GWAS being available for the haemorrhagic subtypes. **Abbreviations** BMI=body-mass index. CI=confidence interval. eGFR=estimated glomerular filtration rate. FDR=false discovery rate. FG=fasting plasma glucose. GWAS=genome-wide association study. IVW=inverse-variance weighted. LDL=low-density lipoprotein cholesterol. MR=Mendelian randomisation. nSNP=number of single-nucleotide polymorphisms retained as instruments. OR=odds ratio. SBP=systolic blood pressure. SNP=single-nucleotide polymorphism.

#### East Asian ancestry

In east Asian ancestry, genetically predicted SBP was associated with every subtype and with unclassified total stroke (all FDR-corrected p<0·05; figure 6, table S38), a more uniform subtype pattern than in European ancestry. For LDL and IS all three estimators were directionally positive but the primary IVW estimate did not reach significance (1·106 [0·987– 1·240]; FDR p=0·14). That interval leaves open effects as large as 1·24, and the analysis could have detected an effect of 1·18 or larger with 80% power, so its null does not distinguish a smaller effect from insufficient precision (table S14). BMI, FG, and eGFR were null across all subtypes, and MVMR showed the same pattern (table S40).

#### South Asian ancestry

South Asian analyses were exploratory and limited to IS, no GWAS data being available for the haemorrhagic subtypes. Instruments were far fewer and weaker than in the other two ancestries (5–13 variants per exposure after harmonisation; mean F 21–41; tables S37–S38), and no exposure was associated with IS after correction. Estimates for BMI (1·390 [0·960– 2·012]) and SBP (1·362 [0·920–2·016]) were directionally consistent with the other ancestries but imprecise.

#### Cross-ancestry pattern

Across ancestries the findings were consistent in direction but unequal in precision (figure 6). SBP was the only exposure associated with stroke in more than one ancestry; LDL detection was confined to IS and to European ancestry, in neither haemorrhagic subtype in either ancestry; and BMI, FG, and eGFR showed no consistent subtype-specific effect. Nulls in this stream were uninformative rather than negative: in all 31 of the 40 ancestry-by-exposure-by-outcome analyses that did not reach significance after correction, the magnitude observed was smaller than the smallest that analysis could have detected with 80% power, so none of them distinguishes a small effect from insufficient precision (table S14).

### Three streams, three orderings

SBP converged and BMI met a listed structural explanation; LDL met the revised M2 criterion: subtype aggregation is a supported qualitative explanation for the cohort non-detection, although its quantitative contribution cannot be estimated without subtype-resolved cohort outcomes. Fasting glucose and kidney dysfunction had unexplained divergence.

## Discussion

In this structurally heterogeneous bloc, priority evidence was not uniformly transportable across levels, member states, or ancestries. Population attribution was spatially structured but only partly aligned with the GBD estimation hierarchy, while ancestry-matched genetic evidence differed in availability and precision; the proximate explanation for the LDL contrast, though not for BMI, was subtype composition. The three C1 states had markedly lower ischaemic-stroke shares than the ischaemia-predominant subtype-composition cluster, but exact cluster membership agreed only modestly. The distal causes cannot be resolved here. With ten locations, geography is collinear with the GBD estimation hierarchy, which borrows strength within region and super-region; adjustment removes it for total stroke and subarachnoid haemorrhage and leaves it nominally for ischaemic stroke and intracerebral haemorrhage. Evidence ordering does not depend on attributing this spatial structure causally to geography; spatial heterogeneity still matters to whether one bloc-wide priority profile is adequate.

### Systolic blood pressure

The three streams converge on SBP: in every state (53–71% of attributable DALYs), the strongest independent cohort exposure (1·37, 1·26–1·48), and supported by genetic evidence in both ancestries with data at scale. In European MVMR its direct effects on the haemorrhagic subtypes exceeded that on ischaemic stroke (1·034, 1·041, and 1·055), although conditional F was borderline; this pattern is compatible with dilution by correlated exposures but does not establish that mechanism. LDL and FG were directionally inverse for both haemorrhagic subtypes in the same data. This is in keeping with the stronger blood-pressure dependence of haemorrhagic stroke reported previously.^3^ Associations attenuated over follow-up and at age 70 years or older; the design cannot establish a common mechanism.

### LDL cholesterol

Subtype aggregation is a supported qualitative explanation for the CHARLS null: the cohort aggregates subtypes, GBD assigns LDL attribution to IS alone, European MR detects LDL– IS while no haemorrhagic estimate survives correction, and the three C1 states had lower ischaemic-stroke shares than the ischaemia-predominant subtype-composition cluster (39–42% vs 58–75%; table S19).^19,20^ Earlier observational and genetic studies likewise reported inverse LDL–ICH associations, although the haemorrhagic estimates here were uninformative after correction. These sources do not quantify how much aggregation contributed because CHARLS subtypes were not observed.

### Body-mass index

BMI produced the most informative divergence: second strongest in the cohort (1·22, 1·14– 1·30) and genetically associated with IS in European ancestry (1·030, FDR p=0·01), yet lowest in population attribution (1·6–11·0%). A threshold counterfactual offers a structural explanation: GBD assigns no burden inside its 20–25 kg/m² minimum-risk window, whereas the cohort had a 1·80-fold within-window hazard gradient (1·48–2·18). Nonlinearity survived correction in the primary spline (FDR p=0·0093) and cause-specific sensitivity analysis (FDR p=0·03), but not after mutual adjustment (FDR p=0·12; appendix 5.7). These data support a possible omission of within-window variation, not a quantified bias in causal attribution.

The gradient inside the window is not peculiar to this cohort. Among 234 863 Korean men, ischaemic stroke risk rose across the whole range measured, by 11% per 1 kg/m², from a reference category of 22–23 kg/m² lying inside the window itself,^21^ and in the China Kadoorie Biobank, where 67·8% of participants had a BMI below 25 kg/m², the association with ischaemic stroke was positive and log-linear throughout the range examined.^22^ Where many people are inside the window, a threshold counterfactual omits any causal gradient within it; its effect on the burden rank cannot be quantified here. The genetic BMI–IS estimate attenuated on mutual adjustment (1·002), compatible with, but not proof of, pathways through other metabolic traits.^23^ At a different level, BMI attribution was two-to-three-fold lower in the C1 states within every subtype, a between-country contrast that composition does not explain.

### Fasting glucose and kidney function

Two factors diverged without a listed mechanism. FG was nonlinear and modest in the cohort, low in attribution, and null in every genetic analysis. Kidney dysfunction carried appreciable attribution (8·8–14·0%), yet neither the cohort nor MR in any ancestry detected an individual-level effect; the eGFR association differed in direction between age strata though neither estimate was significant. Cross-stream interpretation is limited because the population attribution construct is broader than eGFR alone. This construct mismatch limits comparability but does not predict divergence direction and was not admitted as an explanatory mechanism.

### What the three streams together support

BMI had a testable threshold explanation, and LDL a qualitative subtype-aggregation explanation; FG and kidney dysfunction remained unranked.

For a first regional agenda, SBP has the clearest cross-stream case for a shared prevention focus. LDL requires attention to ischaemic subtype mix, and BMI to the minimum-risk window underlying attribution. FG and kidney function remain unresolved for cross-stream priority. These classifications guide which evidence needs checking before priority-setting, rather than quantify intervention benefit.

This study applies an explicit rule set, with one transparently reported pre-submission amendment, to define which disagreements count and which explanations are admissible in a heterogeneous intergovernmental bloc. Triangulation of population, individual, and genetic evidence has been reported for a single country;^6^ the contribution here is an explicit decision procedure in which a factor may remain unranked.

### Transferability

The framework is specific neither to the SCO nor to stroke. It requires a bloc whose members differ in the respects the evidence depends on, so that the extrapolations fail and the streams can disagree. The SCO is an extreme rather than a representative bloc in these respects (tables S1–S2), which makes it a useful test rather than a typical case; independent application to another heterogeneous bloc, such as BRICS, should test the decision rules; its fragmented geography would require separate treatment of the spatial component.

### Limitations

Our evidence is geographically asymmetric. Population estimates cover all ten states, but genetic evidence is absent for five and individual-level data come from China alone. The classification of factors rests on all three streams, whereas between-country heterogeneity is an ecological finding corroborated only indirectly. Building cohort and genomic capacity in the unrepresented ancestry groups is an obvious role for regional coordination.

CHARLS did not distinguish subtypes, so the amended M2 explanation remains qualitative and its magnitude cannot be estimated; its weights resemble China’s.

Finally, the five exposures are those of the Global Burden of Disease Study and are defined as that study defines them. Other indices capture the same constructs and may behave differently: glycated haemoglobin integrates glycaemia over months, whereas a single fasting measurement is sensitive to fasting duration, stress, and day-to-day variation,^24^ and central adiposity measures capture fat distribution that BMI does not, although the largest collaborative analysis found they did not importantly improve cardiovascular risk prediction and were less reproducible than BMI.^25^ This constrains all five factors, including the one on which the streams converged, but is not an explanation for divergence.

## Data Availability

GBD 2023 estimates were obtained from the Institute for Health Metrics and Evaluation GBD Results tool (https://vizhub.healthdata.org/gbd-results/).
The individual-level analysis used the Harmonized CHARLS dataset, Version D (June 2021), together with the wave-1 and 3 blood subsample. The Harmonized CHARLS data are available through the CHARLS data portal after registration and approval (https://charls.pku.edu.cn/en/Data/Harmonized_CHARLS.htm). Individual-level CHARLS data cannot be redistributed by the authors.
The MR analyses used publicly available GWAS summary statistics. European exposure data were obtained from IEU OpenGWAS: SBP, ieu-b-4818 (https://gwas.mrcieu.ac.uk/datasets/ieu-b-4818/); BMI, ieu-b-4816 (https://gwas.mrcieu.ac.uk/datasets/ieu-b-4816/); LDL, ieu-b-4846 (https://gwas.mrcieu.ac.uk/datasets/ieu-b-4846/); eGFR, ieu-b-4832 (https://gwas.mrcieu.ac.uk/datasets/ieu-b-4832/); and fasting glucose, ebi-a-GCST90002232 (https://gwas.mrcieu.ac.uk/datasets/ebi-a-GCST90002232/; GWAS Catalog: https://www.ebi.ac.uk/gwas/studies/GCST90002232).
European outcome data were obtained from FinnGen total stroke, finn-b-C_STROKE (https://gwas.mrcieu.ac.uk/datasets/finn-b-C_STROKE/), and from UK Biobank-FinnGen meta-analyses for ischaemic stroke, ebi-a-GCST90018864 (https://gwas.mrcieu.ac.uk/datasets/ebi-a-GCST90018864/); intracerebral haemorrhage, ebi-a-GCST90018870 (https://gwas.mrcieu.ac.uk/datasets/ebi-a-GCST90018870/); and subarachnoid haemorrhage, ebi-a-GCST90018923 (https://gwas.mrcieu.ac.uk/datasets/ebi-a-GCST90018923/).
East Asian exposure data were obtained from the Taiwan Precision Medicine Initiative PheWeb portal for SBP, BMI, LDL, and fasting glucose (https://pheweb.ibms.sinica.edu.tw/) and from BioBank Japan PheWeb for eGFR (https://pheweb.jp/). East Asian stroke outcome data were obtained from the China Kadoorie Biobank PheWeb portal (https://pheweb.ckbiobank.org/phenotypes).
South Asian exposure data were obtained from Pan-UK Biobank for SBP, ukb-e-4080_CSA (https://gwas.mrcieu.ac.uk/datasets/ukb-e-4080_CSA/); BMI, ukb-e-23104_CSA (https://gwas.mrcieu.ac.uk/datasets/ukb-e-23104_CSA/); LDL, ukb-e-30780_CSA (https://gwas.mrcieu.ac.uk/datasets/ukb-e-30780_CSA/); and eGFR, ukb-e-recode6_CSA (https://gwas.mrcieu.ac.uk/datasets/ukb-e-recode6_CSA/). South Asian fasting-glucose data were obtained from MAGIC, ebi-a-GCST90002234 (https://gwas.mrcieu.ac.uk/datasets/ebi-a-GCST90002234/; GWAS Catalog: https://www.ebi.ac.uk/gwas/studies/GCST90002234). South Asian stroke outcome data were obtained from GIGASTROKE, GCST90104559 (https://www.ebi.ac.uk/gwas/studies/GCST90104559) and GCST90104560 (https://www.ebi.ac.uk/gwas/studies/GCST90104560).
The complete analysis code, including data processing, statistical analyses, and figure-generation scripts, will be deposited in a public repository and released upon publication. The repository link and accession information will be provided in the published article.

## Contributors

Xinyu Wang: conceptualisation, data curation, formal analysis, investigation, methodology, software, validation, visualisation, writing—original draft, and writing—review and editing. Mu Li: data curation, validation, and writing—review and editing. Junzhao Zhang: data curation, validation, and writing—review and editing. Hongqi Zhang: project administration, supervision, and writing—review and editing. Yongjie Ma: project administration, supervision, and writing—review and editing.

Xinyu Wang, Mu Li, and Junzhao Zhang directly accessed and verified the underlying data. All authors had full access to all the data in the study and accept responsibility for the decision to submit for publication. All authors reviewed and approved the final manuscript.

## Data sharing statement

GBD 2023 estimates were obtained from the Institute for Health Metrics and Evaluation GBD Results tool (https://vizhub.healthdata.org/gbd-results/).

The individual-level analysis used the Harmonized CHARLS dataset, Version D (June 2021), together with the wave-1 and 3 blood subsample. The Harmonized CHARLS data are available through the CHARLS data portal after registration and approval (https://charls.pku.edu.cn/en/Data/Harmonized_CHARLS.htm). Individual-level CHARLS data cannot be redistributed by the authors.

The MR analyses used publicly available GWAS summary statistics. European exposure data were obtained from IEU OpenGWAS: SBP, ieu-b-4818 (https://gwas.mrcieu.ac.uk/datasets/ieu-b-4818/); BMI, ieu-b-4816 (https://gwas.mrcieu.ac.uk/datasets/ieu-b-4816/); LDL, ieu-b-4846 (https://gwas.mrcieu.ac.uk/datasets/ieu-b-4846/); eGFR, ieu-b-4832 (https://gwas.mrcieu.ac.uk/datasets/ieu-b-4832/); and fasting glucose, ebi-a-GCST90002232 (https://gwas.mrcieu.ac.uk/datasets/ebi-a-GCST90002232/; GWAS Catalog: https://www.ebi.ac.uk/gwas/studies/GCST90002232).

European outcome data were obtained from FinnGen total stroke, finn-b-C_STROKE (https://gwas.mrcieu.ac.uk/datasets/finn-b-C_STROKE/), and from UK Biobank–FinnGen meta-analyses for ischaemic stroke, ebi-a-GCST90018864 (https://gwas.mrcieu.ac.uk/datasets/ebi-a-GCST90018864/); intracerebral haemorrhage, ebi-a-GCST90018870 (https://gwas.mrcieu.ac.uk/datasets/ebi-a-GCST90018870/); and subarachnoid haemorrhage, ebi-a-GCST90018923 (https://gwas.mrcieu.ac.uk/datasets/ebi-a-GCST90018923/).

East Asian exposure data were obtained from the Taiwan Precision Medicine Initiative PheWeb portal for SBP, BMI, LDL, and fasting glucose (https://pheweb.ibms.sinica.edu.tw/) and from BioBank Japan PheWeb for eGFR (https://pheweb.jp/). East Asian stroke outcome data were obtained from the China Kadoorie Biobank PheWeb portal (https://pheweb.ckbiobank.org/phenotypes).

South Asian exposure data were obtained from Pan-UK Biobank for SBP, ukb-e-4080_CSA (https://gwas.mrcieu.ac.uk/datasets/ukb-e-4080_CSA/); BMI, ukb-e-23104_CSA (https://gwas.mrcieu.ac.uk/datasets/ukb-e-23104_CSA/); LDL, ukb-e-30780_CSA (https://gwas.mrcieu.ac.uk/datasets/ukb-e-30780_CSA/); and eGFR, ukb-e-recode6_CSA (https://gwas.mrcieu.ac.uk/datasets/ukb-e-recode6_CSA/). South Asian fasting-glucose data were obtained from MAGIC, ebi-a-GCST90002234 (https://gwas.mrcieu.ac.uk/datasets/ebi-a-GCST90002234/; GWAS Catalog: https://www.ebi.ac.uk/gwas/studies/GCST90002234). South Asian stroke outcome data were obtained from GIGASTROKE, GCST90104559 (https://www.ebi.ac.uk/gwas/studies/GCST90104559) and GCST90104560 (https://www.ebi.ac.uk/gwas/studies/GCST90104560).

The complete analysis code, including data processing, statistical analyses, and figure-generation scripts, will be deposited in a public repository and released upon publication. The repository link and accession information will be provided in the published article.

## Abbreviations

ASDR: Age-standardised DALY rate
ASEAN: Association of Southeast Asian Nations
ASIR: Age-standardised incidence rate
ASMR: Age-standardised mortality rate
BMI: Body-mass index
BRICS: Proper name of the grouping formed by Brazil, Russia, India, China, and South Africa; not an initialism of its current membership
C1, C2: Metabolic-attribution clusters 1 and 2 (C1 being the cluster containing China)
CHARLS: China Health and Retirement Longitudinal Study
CI: Confidence interval
CIS: Commonwealth of Independent States
DALY: Disability-adjusted life-year
eGFR: Estimated glomerular filtration rate
FDR: False discovery rate
FG: Fasting plasma glucose
GATHER: Guidelines for Accurate and Transparent Health Estimates Reporting
GBD: Global Burden of Diseases, Injuries, and Risk Factors Study
GWAS: Genome-wide association study
ICH: Intracerebral haemorrhage
IS: Ischaemic stroke
IVW: Inverse-variance weighted
KD: Kidney dysfunction
LD: Linkage disequilibrium
LDL: Low-density lipoprotein cholesterol
MR: Mendelian randomisation
MVMR: Multivariable Mendelian randomisation
OR: Odds ratio
PAF: Population attributable fraction
SAH: Subarachnoid haemorrhage
SBP: Systolic blood pressure
SCO: Shanghai Cooperation Organisation
SD: Standard deviation
SDI: Socio-demographic Index
SHAP: SHapley Additive exPlanations
SHR: Subdistribution hazard ratio
SMD: Standardised mean difference
STROBE: Strengthening the Reporting of Observational Studies in Epidemiology
STROBE-MR: Strengthening the Reporting of Observational Studies in Epidemiology using Mendelian Randomization
TMREL: Theoretical minimum-risk exposure level
XGBoost: Extreme gradient boosting

## Declaration of interests

We declare no competing interests.

## Acknowledgments

This study received no funding. No medical writer or editor was involved in the preparation of this manuscript, and we have no further acknowledgments to make.

## Declaration of generative AI and AI-assisted technologies in the manuscript preparation process

During the preparation of this work, the authors used Claude (Opus 4.8 and Opus 5; Anthropic, San Francisco, CA, USA) for three purposes. First, to suggest language edits to author-written drafts of the manuscript and the appendix. Second, to assemble author-produced figure panels into composite figures, by writing the scripts that cropped surrounding white space, aligned and sized panels to the journal’s dimensional limits, and placed panel letters. Third, to check the authors’ analysis code and manuscript for internal consistency and correctness, and to standardise conventions across them: this included checking that reported values matched the source output files, that cross-references between text, tables, and figures corresponded, and that numerical formatting was uniform, together with checks of the analysis scripts for correctness and consistency of settings, from which the authors made and re-ran the resulting revisions. No generative imagery was produced: all figures were rendered by the authors’ own analysis code from the underlying data, and the tool was used only for the deterministic composition of those rendered panels. The tool was not used to generate study design, results, or interpretation. The authors conceived the study, wrote all analysis code, executed all analyses, reviewed and edited all output, verified all reported values against the source data files, take full responsibility for the content of the publication, and confirm that the tool is not listed or cited as an author. Records of the prompts used are retained by the corresponding author and are available to the editors on request.

## Notes

### Competing Interest Statement

The authors have declared no competing interest.

### Author Declarations

GBD 2023 estimates were obtained from the Institute for Health Metrics and Evaluation GBD Results tool (https://vizhub.healthdata.org/gbd-results/). The individual-level analysis used the Harmonized CHARLS dataset, Version D (June 2021), together with the wave-1 and 3 blood subsample. The Harmonized CHARLS data are available through the CHARLS data portal after registration and approval (https://charls.pku.edu.cn/en/Data/Harmonized_CHARLS.htm). Individual-level CHARLS data cannot be redistributed by the authors. The MR analyses used publicly available GWAS summary statistics. European exposure data were obtained from IEU OpenGWAS: SBP, ieu-b-4818 (https://gwas.mrcieu.ac.uk/datasets/ieu-b-4818/); BMI, ieu-b-4816 (https://gwas.mrcieu.ac.uk/datasets/ieu-b-4816/); LDL, ieu-b-4846 (https://gwas.mrcieu.ac.uk/datasets/ieu-b-4846/); eGFR, ieu-b-4832 (https://gwas.mrcieu.ac.uk/datasets/ieu-b-4832/); and fasting glucose, ebi-a-GCST90002232 (https://gwas.mrcieu.ac.uk/datasets/ebi-a-GCST90002232/; GWAS Catalog: https://www.ebi.ac.uk/gwas/studies/GCST90002232). European outcome data were obtained from FinnGen total stroke, finn-b-C_STROKE (https://gwas.mrcieu.ac.uk/datasets/finn-b-C_STROKE/), and from UK Biobank-FinnGen meta-analyses for ischaemic stroke, ebi-a-GCST90018864 (https://gwas.mrcieu.ac.uk/datasets/ebi-a-GCST90018864/); intracerebral haemorrhage, ebi-a-GCST90018870 (https://gwas.mrcieu.ac.uk/datasets/ebi-a-GCST90018870/); and subarachnoid haemorrhage, ebi-a-GCST90018923 (https://gwas.mrcieu.ac.uk/datasets/ebi-a-GCST90018923/). East Asian exposure data were obtained from the Taiwan Precision Medicine Initiative PheWeb portal for SBP, BMI, LDL, and fasting glucose (https://pheweb.ibms.sinica.edu.tw/) and from BioBank Japan PheWeb for eGFR (https://pheweb.jp/). East Asian stroke outcome data were obtained from the China Kadoorie Biobank PheWeb portal (https://pheweb.ckbiobank.org/phenotypes). South Asian exposure data were obtained from Pan-UK Biobank for SBP, ukb-e-4080_CSA (https://gwas.mrcieu.ac.uk/datasets/ukb-e-4080_CSA/); BMI, ukb-e-23104_CSA (https://gwas.mrcieu.ac.uk/datasets/ukb-e-23104_CSA/); LDL, ukb-e-30780_CSA (https://gwas.mrcieu.ac.uk/datasets/ukb-e-30780_CSA/); and eGFR, ukb-e-recode6_CSA (https://gwas.mrcieu.ac.uk/datasets/ukb-e-recode6_CSA/). South Asian fasting-glucose data were obtained from MAGIC, ebi-a-GCST90002234 (https://gwas.mrcieu.ac.uk/datasets/ebi-a-GCST90002234/; GWAS Catalog: https://www.ebi.ac.uk/gwas/studies/GCST90002234). South Asian stroke outcome data were obtained from GIGASTROKE, GCST90104559 (https://www.ebi.ac.uk/gwas/studies/GCST90104559) and GCST90104560 (https://www.ebi.ac.uk/gwas/studies/GCST90104560).

