## appendix for "Triangulating population, individual, and genetic evidence for priority setting in structurally heterogeneous blocs: metabolic risk factors for stroke in the Shanghai Cooperation Organisation"

**Contents**

**Abbreviations used in the appendix**	3

**1. The study bloc and its comparators**	6

1.1 Eligibility and selection of organisations for comparison	6

1.2 GBD super-region assignment	8

1.3 Genetic ancestry group assignment	9

1.4 Adjacency and connected components	11

**2. Spatial structure and the estimation hierarchy**	12

2.1 Rationale and the hierarchy distance	12

2.2 What this analysis can and cannot distinguish	13

2.3 The cluster structure does not follow the estimation hierarchy	13

2.4 Ischaemic stroke	14

2.5 The gradient within a single super-region	14

2.6 Why uncertainty intervals were not used	15

2.7 Subtype composition does not mediate the hierarchy association	15

2.8 Multiple comparisons	16

**3. Evidence licensing, ordering, and decision rules**	16

3.1 Within-stream ordering	16

3.2 Admissible mechanisms, and explanations not admitted	17

3.3 Sensitivity of the orderings	19

3.4 Why estimates are not compared in magnitude across ancestries	19

**4. GBD component**	20

4.1 Data and burden description	20

4.2 Burden-trajectory clustering	20

4.3 Age and sex structure	21

4.4 Metabolic risk-attribution clustering	21

4.5 Age-specific and sex-specific attribution	22

4.6 Stroke-subtype composition clustering and its concordance	23

4.7 Spatial structure	24

4.8 Cluster stability	24

4.9 SDI–burden relationship and efficiency frontier	25

**5. CHARLS component**	26

5.1 Data sources and eligibility	26

5.2 Baseline characteristics	26

5.3 Exposures and kidney function	27

5.4 Outcome, competing event, and follow-up time	27

5.5 Covariates	27

5.6 Competing-risk modelling	28

5.7 Nonlinearity and proportional subdistribution hazards	28

5.8 Subgroup analyses	31

5.9 Statistical inference and multiplicity	32

5.10 Sensitivity analyses	32

5.11 Auxiliary variable-importance analysis	33

**6. Multi-ancestry MR component**	34

6.1 Data sources and ancestry mapping	34

6.2 Instrument construction	34

6.3 Univariable MR and sensitivity analyses	35

6.4 Multivariable MR	35

6.5 European ancestry	35

6.6 East Asian ancestry	37

6.7 South Asian ancestry	38

6.8 Cross-ancestry pattern and multiplicity	39

**7. Software and packages**	39

**8. Ethics and sex reporting**	40

**References**	40

**Legends**	44

**Abbreviations used in the appendix**

AI = Artificial intelligence

AIDS = Acquired immunodeficiency syndrome

ASDR = Age-standardised DALY rate

ASEAN = Association of Southeast Asian Nations

ASIR = Age-standardised incidence rate

ASMR = Age-standardised mortality rate

AUC = Area under the receiver operating characteristic curve

BMI = Body-mass index

BRICS = Proper name of the grouping formed by Brazil, Russia, India, China, and South Africa; not an initialism of its current membership

C1, C2 = Metabolic-attribution clusters 1 and 2 (C1 being the cluster containing China)

CES-D = Center for Epidemiologic Studies Depression Scale

CHARLS = China Health and Retirement Longitudinal Study

CI = Confidence interval

CIS = Commonwealth of Independent States

CKB = China Kadoorie Biobank

CKD-EPI = Chronic Kidney Disease Epidemiology Collaboration

DALY = Disability-adjusted life-year

ΔPAF = Difference between a subgroup-specific and the all-age, both-sex population attributable fraction

eGFR = Estimated glomerular filtration rate

FDR = False discovery rate

FG = Fasting plasma glucose

GBD = Global Burden of Diseases, Injuries, and Risk Factors Study

GIGASTROKE = Name of the stroke genome-wide association consortium; not an initialism

GWAS = Genome-wide association study

HIV = Human immunodeficiency virus

ICD-10 = International Classification of Diseases, 10th revision

ICH = Intracerebral haemorrhage

IHME = Institute for Health Metrics and Evaluation

ilr = Isometric log-ratio

IS = Ischaemic stroke

IVW = Inverse-variance weighted

lasso = Least absolute shrinkage and selection operator

LD = Linkage disequilibrium

LDL = Low-density lipoprotein cholesterol

MAGIC = Meta-Analyses of Glucose and Insulin-related traits Consortium

MR = Mendelian randomisation

MVMR = Multivariable Mendelian randomisation

OR = Odds ratio

PAF = Population attributable fraction

REML = Restricted maximum likelihood

RLMS-HSE = Russia Longitudinal Monitoring Survey–Higher School of Economics

SAARC = South Asian Association for Regional Cooperation

SAH = Subarachnoid haemorrhage

SAIGE = Scalable and Accurate Implementation of GEneralized mixed model

SBP = Systolic blood pressure

SC1, SC2 = Subtype-composition clusters 1 and 2 (SC1 being the cluster containing China)

SCO = Shanghai Cooperation Organisation

SD = Standard deviation

SDI = Socio-demographic Index

SE = Standard error

SHAP = SHapley Additive exPlanations

SHR = Subdistribution hazard ratio

SMD = Standardised mean difference

STEPS = WHO STEPwise approach to noncommunicable disease risk factor surveillance

TMREL = Theoretical minimum-risk exposure level

TPMI = Taiwan Precision Medicine Initiative

WHO = World Health Organization

XGBoost = Extreme gradient boosting

**1. The study bloc and its comparators**

The Shanghai Cooperation Organisation was fixed as the study bloc at the outset. The criteria set out here were specified afterwards, to locate it among the blocs that might have been chosen and to define the conditions under which the design transfers. They are derived from the requirements of the design rather than from the features of any bloc: evidence-relevant structural heterogeneity is indexed on the dimensions along which the three evidence streams fail to transport—cause-of-death patterns for the population stream, pooled relative risks for attribution, and reference panels for the genetic stream—and geographical coherence is indexed because the spatial component is interpretable only where proximity tracks shared exposure environment. A reader can check that derivation against the design without reference to when it was made. That the comparison returns organisations exceeding the SCO on two of the three dimensions is the evidence that the criteria were not constructed around it.

**1.1 Eligibility and selection of organisations for comparison**

An intergovernmental organisation was eligible for comparison if it met all four of the following: at least five sovereign member states; a permanent secretariat or equivalent standing administrative body; at least one health-sector ministerial meeting or standing health body; and membership acquired by formal accession rather than by invitation or observer status. The third criterion is met by a ministerial track or a standing health institution that remains in operation, irrespective of the status of the organisation’s summit process; SAARC qualifies on the second limb through the SAARC Tuberculosis and HIV/AIDS Centre, although its summit process has been suspended since 2016.

These criteria are necessary but not sufficient for inclusion, and the thirteen organisations analysed are a purposive rather than an exhaustive set. They were chosen to span the range of both selection dimensions and to include every organisation whose membership substantially overlaps the SCO’s. The set comprises the blocs most often used as units in regional health analysis (ASEAN, the European Union, the Nordic Council, the African Union); those closest to the SCO in geography and membership (the CIS, the Eurasian Economic Union, SAARC); those exceeding the SCO on structural heterogeneity (BRICS, the Commonwealth, the Organisation of Islamic Cooperation, the Arab League); and the Gulf Cooperation Council as a maximally homogeneous anchor. No organisation was omitted because of where it fell on either dimension. Other organisations meeting the eligibility rule—in Latin America, sub-Saharan Africa, the Pacific, Europe, and the Americas as a whole—are not analysed individually; each is confined to one or two GBD super-regions, and the island groupings among them are highly fragmented, so none can satisfy the conjunction at issue. Their values can be computed from the code supplied with this study.

The set being purposive is a limitation, but a bounded one. Adding organisations cannot alter the SCO’s own values, so the claim that it is the only bloc combining four or more GBD super-regions with land contiguity is directly falsifiable: it requires identifying an eligible organisation that is both, and the code supplied with this study computes all three metrics for any membership list. We are not aware of one. Spanning four or more super-regions while remaining land-contiguous requires a membership traversing Eurasia from eastern Europe to south and east Asia, which is a demanding geographical condition rather than an artefact of the comparison set.

Membership was taken as at 2026. Timor-Leste is counted as an ASEAN member, having acceded on Oct 26, 2025. Turkmenistan is an associate rather than a full member of the CIS and is excluded; Ukraine’s and Georgia’s withdrawals from the CIS completed in 2018 and 2009 respectively. Saudi Arabia is excluded from BRICS, having been invited in 2023 without its accession being confirmed; including it would add no super-region, no ancestry group, and no edge, and would change no value in tables S1 or S2.

The standing health mechanisms and target-adoption status recorded in table S2 were taken from secretariat documents where available and from official communiqués otherwise, searched to July 24, 2026. For the SCO, health cooperation entered the organisation’s framework in 2011,^1^ and a dedicated centre for cooperation on metabolic disease was established in February, 2026.^2^ Target-adoption status is recorded as the result of that search rather than as an assertion that no instrument exists: for each organisation, a bloc-wide numerical target for non-communicable diseases or their principal metabolic risk factors was either located or not located by that date.

**1.2 GBD super-region assignment**

Each member state was assigned to one of the seven GBD super-regions using the IHME location hierarchy for GBD 2023.^3^ Assignments for all 155 countries appearing in the thirteen organisations are given in table S3.

We used the GBD hierarchy rather than a geographical or United Nations classification because the super-regions are themselves defined by similarity of cause-of-death patterns, and because the attribution estimates compared in this study are produced within that structure. The count of super-regions spanned is therefore internal to the data source rather than an external judgment about which countries resemble one another. It is nonetheless an ordinal index on a seven-category partition: a bloc spanning six super-regions is not heterogeneous by half as much again as one spanning four, and no arithmetic is performed on these counts.

Three assignments are counter-intuitive and materially affect the counts. Maldives and Sri Lanka are assigned to southeast Asia, east Asia, and Oceania rather than to south Asia, which is why SAARC spans two super-regions rather than one. Mauritius and Seychelles are assigned to the same super-region rather than to sub-Saharan Africa, which is why the African Union spans three rather than two. Brunei and Singapore are assigned to High-income, which is why ASEAN spans two super-regions rather than one; Timor-Leste’s accession does not change this. In each case the effect is to increase, not decrease, the apparent structural heterogeneity of a comparator relative to the SCO.

Locations not represented in the GBD hierarchy were excluded from all blocs; member counts in table S1 are therefore of GBD locations rather than of formal memberships.

**1.3 Genetic ancestry group assignment**

Table S1 counts the number of genetic ancestry groups represented in each organisation, as an index of how far genetic association estimates derived in one member state can be expected to transport to another. The count is deliberately coarse. It indexes the availability of matched reference data, not the structure of human genetic variation, which is continuous and is not partitioned by these labels.^4^

Each member state was assigned to exactly one group, corresponding to the ancestral background of its largest population. Groups were the five 1000 Genomes Project superpopulations (African, admixed American, east Asian, European, and south Asian),^5^ supplemented by the Middle Eastern group defined in the Genome Aggregation Database.^6^ Two further groups were added for populations corresponding to no current large-scale reference panel: Central Asian, comprising the Turkic and Indo-Iranian populations of Kazakhstan, Kyrgyzstan, Tajikistan, Turkmenistan, and Uzbekistan; and Oceanian, comprising Papuan and Melanesian populations. The groups and their reference panels are listed in table S4, and the complete assignment is in table S3.

Two assignments are boundary cases and were resolved by explicit rule. Timor-Leste has an Austronesian-speaking majority east of Wallace’s Line with substantial Papuan admixture, matched well by neither the east Asian nor an Oceanian panel; it was coded east Asian on the rule that the predominant population is mapped to the nearest available panel. Coding it Oceanian instead would give ASEAN two ancestry groups, one of them without a matched panel, and would not alter its position relative to the SCO on either selection dimension. The Central Asian group is discussed below.

Three further assignments are contestable, and each is recorded because a different but equally defensible coding would change a comparator’s count. Azerbaijan is coded Middle Eastern rather than Central Asian, and is the sole member of that group within the CIS, whose count would fall from three groups to two under the alternative. Armenia is coded European rather than Middle Eastern; under the alternative the Eurasian Economic Union’s count would rise from two to three. Albania is the sole European member of the Organisation of Islamic Cooperation, so that organisation’s European group rests on one state.

A count is fragile where the presence of a group turns on a contestable assignment, and by that test the SCO’s is not. Two of its five groups are represented by a single member state, China and Iran, but neither assignment is in question: China anchors the east Asian group, and Iranian populations are among those from which the Middle Eastern reference data are drawn.^7^ Four of the five groups—European, south Asian, east Asian, and Middle Eastern—are unambiguous under any reasonable coding. The fifth is the Central Asian group, whose treatment is examined in table S5.

Four limitations follow from this design, and none is incidental.

First, a single group per country cannot represent within-country ancestral diversity. India, China, Russia, Iran, and Pakistan each contain populations differing substantially in genetic ancestry, in some cases by more than the difference between the assigned groups of two other countries.^8^ The counts are therefore a lower bound for every organisation, and organisations containing large, internally diverse states are understated relative to those of smaller, more homogeneous ones.

Second, the reference panels are not of equal size or quality, and treating them as interchangeable units of a count obscures this. The European group is represented by orders of magnitude more genotyped individuals than the south Asian or Middle Eastern groups, so two organisations with the same count may face very different constraints on instrument availability.^9^

Third, the boundaries between groups are conventions of the reference panels rather than natural divisions, and the Central Asian group is the most consequential such convention, because it is the group that raises the SCO’s count. We therefore recomputed all counts under three alternatives in which Central Asian populations are treated as adequately represented by an existing panel rather than as unrepresented (table S5). Under every alternative the SCO’s count falls from five groups to four, and under every alternative the SCO still represents more ancestry groups than any organisation forming fewer than five connected components. The ordering of organisations on this measure is unchanged. What the alternatives assume, and what the baseline declines to assume, is that a European or east Asian panel provides adequate instrument transportability for Turkic and Indo-Iranian populations; that assumption is not established, which is why the baseline treats the group as unrepresented.

Fourth, and most consequentially, the Central Asian group has no matched panel, so the ancestral background of four of the ten SCO member states is not represented in any current large-scale reference panel. Instrument availability is more restrictive still: of the five groups represented in the bloc, only the European, east Asian, and south Asian have stroke genome-wide association data at the scale instrument selection requires, so five member states—Iran, Kazakhstan, Kyrgyzstan, Tajikistan, and Uzbekistan—contribute no genetic evidence. The two criteria are distinct and are not interchangeable: panel availability governs four states and one group, instrument availability governs five states and two groups. Which is in play is stated wherever either is used.

**1.4 Adjacency and connected components**

Two member states were treated as adjacent if they share a land border, taking country polygons at 1:50 000 000 scale from Natural Earth (public domain) through version 1.0.0 of the rnaturalearthdata R package.^10^ The package does not record which upstream Natural Earth release it bundles, so the package version is the reproducibility reference: different package versions carry different snapshots, and the treatment of self-declared territories differs between them. All component counts reported here were computed against that version.

Each organisation’s membership was represented as an undirected graph whose vertices are member states and whose edges are shared land borders internal to that organisation; the number of connected components was obtained by depth-first search. Borders with non-member states create no edge, so a bloc whose members are contiguous only by way of a third country forms more than one component.

The number of components is a binary summary in the case that matters here, and does not distinguish a membership held together by a single chain of borders from one that is densely interlinked. We therefore also report the algebraic connectivity of the largest component, the second-smallest eigenvalue of its Laplacian.^11^ It is zero if and only if the component is disconnected, and increases with the number of internal borders and with the number of alternative paths between member states, so it indexes how much of the component’s contiguity would survive the loss of any single border. It declines as the number of member states in a component rises, and is not comparable between components of very different size; it is therefore reported only for components of eight or more member states, and is used to describe the organisations rather than to rank them. For the SCO, whose ten member states form a single component joined by fifteen internal land borders, the value is 0·475, the highest of the five organisations for which it is reported.

Maritime boundaries create no edge. The choice is consequential for island and archipelagic memberships—ASEAN, the Commonwealth, the African Union, and the Organisation of Islamic Cooperation each fragment further under it than they would under a rule admitting short sea crossings—and it is made because the property being indexed is the practicability of contiguous land-based surveillance, harmonised border standards, and joint programmes, which a sea crossing does not confer. The rule is applied identically to every organisation, including the SCO, which contains no maritime-only link and is therefore unaffected by it.

Component counts are of GBD locations, consistent with section 1.2, so a formal member absent from the GBD hierarchy contributes neither a vertex nor an edge. Values for all thirteen organisations are in table S2 and are plotted in figure 1A.

**2. Spatial structure and the estimation hierarchy**

**2.1 Rationale and the hierarchy distance**

GBD estimates for locations with limited input data are informed by other locations in the same region and super-region. Where such borrowing is substantial, countries that share a position in the estimation hierarchy will resemble one another for reasons internal to the modelling, and any spatial concordance among the resulting estimates is ambiguous between a real distribution of exposure and the structure of the model. We made this alternative explicit and testable by defining a hierarchy distance between each pair of member states: 0 where both belong to the same GBD region, 1 where they belong to the same super-region but different regions, and 2 otherwise. The ten member states span five regions within four super-regions. India and Pakistan belong to South Asia, whose other members are Afghanistan, Bangladesh, Bhutan, the Maldives, Nepal, and Sri Lanka, none of which enters this analysis; China belongs to East Asia. No borrowing pathway connects them.

Association between distance matrices was assessed by the Mantel test with 99 999 permutations,^12^ and the hierarchy was controlled for by the partial Mantel test, which permutes one matrix while holding a third fixed.^13^ Both are permutation procedures on non-independent pairwise distances, and neither yields a statistic whose magnitude has an interpretation independent of the number of objects; with ten states and forty-five pairs they are used to order competing explanations, not to estimate an effect.^14^

**2.2 What this analysis can and cannot distinguish**

GBD super-regions are themselves constructed from epidemiological similarity, and epidemiological similarity is patterned by ancestry, diet, and environment, which are in turn patterned by geography. A stronger association with the hierarchy than with physical distance is therefore compatible with two readings: that borrowing generated the pattern, and that attribution profiles agree with an independently constructed epidemiological typology, which would be a stronger form of the same finding. The analyses below narrow this ambiguity from two sides but do not remove it, and we do not claim that an artefactual explanation has been excluded. These analyses address the Mantel statistic only. The join-count statistic did not reach significance at baseline for any outcome, so there is no positive result whose robustness to adjustment could be assessed; no standard partial form of a categorical spatial-autocorrelation statistic is available in any case, and the join-count results should be read as unadjusted throughout.

**2.3 The cluster structure does not follow the estimation hierarchy**

If borrowing were the sole source of similarity among these estimates, their natural structure would be the four GBD super-regions represented in the bloc. It is not recovered at any number of clusters from two to five (table S10), and the departures are informative in a way a single agreement statistic is not.

One departure holds at every number of clusters: China is never separated from India, Pakistan, or both, so the boundary between the southeast Asia, east Asia, and Oceania super-region and south Asia is never recovered. This is the departure the borrowing hypothesis should most strongly prevent, since those states lie in different super-regions and no borrowing pathway connects them. It is also the division that carries the attribution contrast reported in the main text.

The remaining departures are specific to the resolution. From three clusters upward the Central Europe, Eastern Europe, and Central Asia super-region is itself divided, Belarus and Russia grouping with Kazakhstan and Kyrgyzstan while Tajikistan and Uzbekistan separate; at two clusters that super-region is not divided, but only because Iran is absorbed into it. Iran, which constitutes a super-region on its own within the bloc, is absorbed into the Eurasian group at two clusters, joined to Tajikistan and Uzbekistan at three, and isolated—consistent with its super-region—only at four and five. At five clusters south Asia divides in turn, India separating while Pakistan remains with China.

Agreement with the super-region partition is therefore not monotone in the number of clusters (adjusted Rand index 0·651, 0·384, 0·471, and 0·454 at two, three, four, and five clusters). The two-cluster solution agrees best, but a coarse partition will tend to, since it leaves undivided most of the pairs the super-regions separate. What survives at every resolution is one specific failure of the hierarchy to account for the grouping; what varies is where the additional structure falls.

**2.4 Ischaemic stroke**

Ischaemic stroke was the exception and the clearest case. Cluster C1 comprised China, India, and Pakistan together with Tajikistan and Uzbekistan, dividing the Central Europe, Eastern Europe, and Central Asia super-region. The two states that moved are the Central Asian members closest to China and to south Asia, so the partition extends C1 in a geographically contiguous direction across a super-region boundary, which borrowing cannot produce. Consistently, the cluster assignment for this outcome showed no detectable correspondence with the hierarchy (adjusted Rand index 0·043 against region, p=0·49; 0·082 against super-region, p=0·29), while the geographic association survived adjustment for it.

**2.5 The gradient within a single super-region**

Six member states lie in the Central Europe, Eastern Europe, and Central Asia super-region and are treated identically by super-region-level borrowing. Among them the geographic gradient persisted (r=0·58, p=0·01 for total stroke; table S11), which super-region membership cannot explain. The gradient did not survive adjustment for the finer region level (r=0·13, p=0·31), but that comparison is not identified: within these six states the division between Eastern Europe and Central Asia coincides almost exactly with the geographic division, and fifteen country pairs cannot separate them. We report the adjusted value for completeness and place no weight on it.

**2.6 Why uncertainty intervals were not used**

We did not perturb the point estimates within their uncertainty intervals. The uncertainty in a population attributable fraction derives substantially from the pooled relative risk and the theoretical-minimum-risk exposure distribution, both of which are common to all countries and therefore cancel in a between-country comparison of profiles standardised within each risk factor; the remaining country-specific component cannot be separated from the published summaries. Interval width was also unusable as a proxy for input-data quality: across the ten member states it was, if anything, wider where cause-of-death data were better rated (Spearman ρ=0·56, p=0·09), the opposite of the direction such a proxy requires. Exclusion was therefore based directly on the published star ratings of cause-of-death data completeness and quality, which grade national vital registration on a five-point scale.^15^

That exclusion is reported in table S12, and its direction deserves to be stated. If the geographic concordance were produced by borrowing into member states whose input data are sparse, removing the two states with the weakest cause-of-death ratings should weaken it. It did the opposite: the total-stroke association rose from 0·498 in the full set to 0·671 in the remaining eight, and it rose for all three subtypes as well. Omitting any single member state also left the association in place, the lowest value being 0·409 on omitting Belarus. Neither result establishes that the concordance is substantive: eight member states support only 28 pairs, the comparison is not a formal test, and the two states removed are also two of the three members of cluster C1, so the partition necessarily rearranges. Both run counter, however, to what the data-quality explanation predicts, and they are reported for that reason rather than as evidence for the alternative.

**2.7 Subtype composition does not mediate the hierarchy association**

Because attribution for total stroke is a composition-weighted combination of subtype-specific attribution, and because subtype composition is itself spatially structured, we tested whether composition accounted for the hierarchy association. It did not: with composition distance held constant, the association between the hierarchy and attribution profiles was essentially unchanged (r=0·55, p=0·0008, against r=0·66 unadjusted). This statistic holds subtype composition constant and is not the one reported in table S13, which holds geographic distance constant (r=0·556, p=0·0024); the two are close in magnitude but answer different questions. We offer no alternative mechanism for the difference between total stroke and its two main subtypes, and note only that the partial statistic declines as the hierarchy association exceeds the geographic one.

**2.8 Multiple comparisons**

The analyses reported in this section comprise about 75 tests across four outcomes. They are sensitivity analyses of a prespecified primary result and are not included in the false-discovery-rate control applied to the two confirmatory families described in the Methods; the p values are nominal. The adjusted geographic associations for ischaemic stroke and intracerebral haemorrhage would not survive correction across the four outcomes, and we accordingly describe them as persisting rather than as significant.

**3. Evidence licensing, ordering, and decision rules**

The conclusions each stream was taken to license are stated in the Methods; what follows is how each stream was ordered internally, how a divergence between two streams was identified and classified, and what the rules decline to do. The framework follows the principle that evidence from sources with different and largely non-independent weaknesses is informative in proportion to how far those weaknesses differ.^16^

**3.1 Within-stream ordering**

**Population stream**

Factors were ordered by the median 2023 population attributable fraction of total-stroke DALYs across the ten member states (SBP 63·1%, LDL 17·0%, kidney dysfunction 11·4%, fasting glucose 9·6%, BMI 8·7%). Two properties are reported as evidence of the ordering’s stability rather than used to construct it: the modal within-state rank reproduces the same sequence, and every adjacent pair is ordered consistently in at least eight of the ten states (10, 9, 8 and 8 for the four adjacent pairs; table S15, final column). Ordering by attributable age-standardised DALY rate reproduces the within-state ranks exactly; this is arithmetically necessary, the two differing within a state by a single multiplicative constant, and is therefore not reported as a robustness check. No bloc-level aggregate attributable fraction was computed: aggregating the ten states into a single fraction presupposes the transferability this study sets out to examine, and the result would be determined almost entirely by the two most populous members.

**Individual stream**

Factors not flagged not detected were ordered by the subdistribution hazard ratio per 1 SD from the mutually adjusted model. The auxiliary gradient-boosted importance analysis was a prespecified corroboration of this ordering, not an input to it. The further-adjusted model was not used for ordering, adjustment for medication removing variation that is a consequence of the exposure.

**Genetic stream**

Factors were ordered lexicographically on three criteria applied in sequence: the number of ancestry-by-subtype comparisons—European and east Asian ancestry crossed with ischaemic stroke, intracerebral haemorrhage and subarachnoid haemorrhage, six in all—in which the univariable inverse-variance-weighted estimate was positive in direction and passed FDR correction; persistence in multivariable analysis; and the order of magnitude of the smallest FDR-corrected p value among the comparisons counted under the first criterion. All three criteria are properties of direction and significance; none uses the magnitude of an estimate, for the reasons given in section 3.4. South Asian analyses were exploratory, limited to ischaemic stroke, and did not contribute.

**Not detected**

A factor was flagged not detected in the individual stream when the 95% CI of the mutually adjusted subdistribution hazard ratio included 1, and in the genetic stream when no ancestry-by-subtype comparison was counted under the first criterion. Factors so flagged occupy the lowest position in that stream jointly and were not ordered among themselves. The population stream has no equivalent flag. It can register absence, but only as a global decision not to pair a risk factor with the outcome—low bone mineral density, in the same metabolic cluster, is paired with no stroke subtype—and such a decision applies uniformly to all locations. Once a pair is included the attributable fraction is positive in every location by construction, so the population stream cannot return a bloc-specific or country-specific null.

**3.2 Admissible mechanisms, and explanations not admitted**

A divergence was classified as structurally explicable only where at least one mechanism in table S16 predicted both its presence and its direction and where that mechanism’s precondition held. The requirement on direction is what allows the rules to fail: M1 predicts that the population stream will rank a factor lower than the individual stream does, and the kidney-dysfunction divergence runs the other way, so M1 is unavailable there even though a threshold counterfactual exists for that risk factor. After internal review but before submission, M2 was amended and the amendment is reported here to require a subtype-specific signal, aggregation of subtypes in the individual stream, and population-stream evidence of material heterogeneity in subtype composition, with genetic evidence supporting the subtype restriction; it does not require inference of the unobserved CHARLS subtype proportion. Table S17 gives the derivation. For LDL, M2 is admitted as a qualitative explanation because GBD assigns LDL attribution to IS alone, European MR detects LDL–IS while no haemorrhagic estimate survives correction, and the three C1 states had markedly lower ischaemic-stroke shares than the ischaemia-predominant subtype-composition cluster (39–42% vs 58–75% of stroke DALYs; table S19). The evidence does not quantify how much subtype aggregation contributed to the CHARLS null because cohort subtypes were not observed. The counterfactual exposure distributions to which M1 refers are those of the comparative risk assessment underlying the attribution estimates.^17^

Table S18 lists three explanations compatible with patterns observed here but not admitted. Each is a substantive claim about a particular exposure rather than a structural property of the streams, and in each case this design cannot separate it from a named alternative. Insufficient power is not admitted as an explanation unless it is quantified and reported rather than asserted; inferring adequate power from instrument strength is not a ground, instrument strength governing the exposure side rather than the precision of a ratio estimate. A fourth candidate, attenuation of an observational estimate by unstable exposure measurement, cannot account for any divergence in which the observational stream is the positive one, non-differential measurement error attenuating towards the null.

Unexplained divergence is an output of the framework rather than a failure of it. It records that the streams disagree and that the disagreement is not accounted for by any difference in what the streams are licensed to estimate. For priority setting it means the available evidence does not license a rank for that factor, whichever stream is consulted alone.

**3.3 Sensitivity of the orderings**

Two orderings depend on the rule used, and neither dependence changes a classification. In the genetic stream, LDL and BMI are tied on the first criterion (one counted comparison each) and separated by the second; applying the third criterion in place of the second leaves the order unchanged, the smallest corrected p value being 0·0042 for LDL against 0·01 for BMI. Under either rule LDL remains divergent from the cohort by the not-detected criterion, and BMI from the population stream by the positional criterion. Separately, the positional criterion for BMI requires that the three lowest-ranked factors in the population stream be ordered rather than tied; their median attributable fractions are close (11·4%, 9·6%, 8·7%), and under a rule that tied them the BMI divergence would be detected only by the precondition test for M1, which is independent of any ordering.

**3.4 Why estimates are not compared in magnitude across ancestries**

No contributing exposure study states the units in which its effect estimates are expressed, nor the standard deviation of the exposure in its own sample. Any common scale is therefore an inference of ours rather than a fact about the source, and we report none. Three further obstacles would remain even if the units were known. For SBP, LDL, BMI, and eGFR, we used the population GWAS estimates from a consortium that also conducted within-sibship analyses; our estimates are not within-family adjusted.^18^ Instrument sets and linkage disequilibrium structure differ, so the same causal variants are tagged with different fidelity. Outcome sources and case definitions differ. Ancestries are accordingly compared by direction of effect and significance only, and no comparison of absolute effect size appears anywhere in this study.

What replaces such a comparison is a statement internal to each analysis. For every ancestry-by-exposure-by-outcome analysis we report the smallest odds ratio that analysis could have detected with 80% power at a two-sided alpha of 0·05, obtained as exp(2·802 × SE) on the log-odds scale,^19^ together with the largest effect its interval did not exclude. Both quantities lie on that study’s own scale, so the comparison between them is internal to one row and needs no conversion; neither may be compared between rows. Where the observed magnitude falls below the detectable threshold, the analysis cannot separate a smaller true effect from insufficient precision. The threshold at which a factor was taken to be detected was the FDR-corrected p value, which is stricter than the uncorrected alpha used for the detectable effect, so the reported threshold is a lower bound on what detection required.

We record, for completeness, an analysis we did and do not report. An earlier version of this work recovered the standard deviation of each exposure from its own summary statistics, placed all estimates on a per-SD scale, and tested twelve cross-ancestry contrasts by Cochran’s Q; none differed after correction. That procedure required either an external population standard deviation, taken from a cohort of a different ancestry and age structure, or a recovered value carrying no error term into the intervals built on it. The first assumes precisely the transportability this study declines to assume elsewhere; the second understates uncertainty. We therefore withdrew the harmonisation rather than report a contrast the design cannot support, and retain from it only the finding that motivated the rule: the apparent several-fold difference between European and east Asian SBP estimates cannot be interpreted biologically while the units and outcome definitions remain unharmonised.

**4. GBD component**

**4.1 Data and burden description**

National-level GBD 2023 estimates were obtained for the ten SCO member states for 2001–2023.^17^ ASIR, ASMR, and ASDR were used for all analyses except the description of age structure, for which age-specific rates by sex were displayed as heatmaps (log10 scale) for incidence, mortality, and DALYs, ordered against the global rate. Country names were harmonised (Russian Federation→Russia; Iran [Islamic Republic of]→Iran).

**4.2 Burden-trajectory clustering**

For each outcome and each of the three rate measures, the 2001–2023 series for every country was summarised by five features: the linear slope (per-year change expressed as a percentage of the 2001 value), the total percentage change, volatility (the SD of year-on-year differences relative to their mean), trend acceleration (the relative change in slope between the first and second halves of the period), and the 23-year mean level. The features (five per measure across three measures) were standardised to zero mean and unit variance with equal weighting, and countries were clustered by Ward's method on Euclidean distances.^20^ The number of clusters was selected by the maximum mean silhouette width over k=2–4.^21^ Clusters were visualised with dendrograms and cluster-faceted trend plots.

For total stroke, the mean silhouette width was maximised at three clusters (0·19, 0·22, and 0·18 at two, three, and four clusters). For the subtypes, the silhouette width selected four clusters for IS (0·20) and SAH (0·16) and three for ICH (0·27), with memberships that differed from the total-stroke partition and between subtypes (figures S1–S3, table S20). Absolute silhouette widths were low throughout (0·16–0·27), as expected when clustering ten countries, but the selected partitions were the most reproducible in resampling.

**4.3 Age and sex structure**

Across all member states and both sexes, stroke incidence, mortality, and DALY rates increased monotonically and approximately log-linearly with age, from very low rates before age 50 years to the highest rates at 80 years or older (figure S5); the pattern was concordant across the three subtypes (figure S6). Male and female rates were broadly similar and showed no consistent direction of difference across countries, although female rates exceeded male rates at the oldest ages in some high-burden states (eg, Russia and Kazakhstan).

**4.4 Metabolic risk-attribution clustering**

For 2023, the PAF (the GBD "percent" metric for each risk–stroke–DALY combination) and the attributable DALY rate (the "rate" metric) were extracted for the five metabolic risk factors. Countries were clustered on the five PAFs after standardising each risk factor across countries (row z-score), using Ward's method on Euclidean distances, with the number of clusters selected by the maximum mean silhouette width over k=2–5 (main analysis k=2). A sensitivity clustering used a composite of PAF × log(1 + attributable DALY rate); agreement with the main clustering was quantified by the adjusted Rand index.^22^ PAF and attributable-rate matrices were displayed as paired heatmaps with within-country risk-factor ranks. In both the attribution and the subtype-composition clusterings the cluster containing China was designated C1 (SC1 for subtype composition) and the other cluster C2 (SC2), so that China-containing clusters can be compared descriptively; the labels do not imply identical membership.

The metabolic-attribution clustering selected two clusters by maximum mean silhouette width for total stroke (0·34) and all subtypes (IS 0·28; ICH 0·33; SAH 0·58; table S24). For total stroke the composite-metric sensitivity clustering (PAF × log attributable rate) returned an identical partition (adjusted Rand index 1·00); subtype agreement was weaker (adjusted Rand index 0·29–0·61).

The two clusters differed on all five factors, but not equally. In absolute terms the between-cluster gap was largest for LDL (C1 10·6–12·9% vs C2 12·1–24·2%), which is also the difference of greatest epidemiological consequence; on the standardised scale used for clustering, however, BMI separated the two groups most cleanly (C1 1·6–4·8% vs C2 8·1–11·0%, non-overlapping), whereas the LDL ranges overlapped through Tajikistan (12·1%). The 17–24% range quoted for C2 in the main text and abstract therefore excludes Tajikistan, whose LDL PAF of 12·1% falls inside the C1 range; taking all seven states together, C2 spans 12·1–24·2%. This single-variable overlap did not translate into cluster instability: resampling reproduced the three-country and seven-country partition exactly for every state, including Tajikistan (co-assignment frequency 0 or 1 throughout; figure S11).

**4.5 Age-specific and sex-specific attribution**

Sex-specific and age-specific PAFs were compared with the all-age, both-sex estimates as ΔPAF heatmaps in percentage points, with countries ordered by the main attribution clustering.

For total stroke, attribution differed more by age than by sex. In the 50–69-year subgroup, the PAFs of most metabolic factors exceeded the all-age estimate—most markedly for SBP (mean ΔPAF +4·3 percentage points), BMI (+2·7), and LDL (+2·0)—whereas FG was the exception, contributing slightly less than at all ages (−1·0). In the 70-years-or-older subgroup the pattern reversed for the adiposity-related factors: BMI fell well below its all-age attribution (mean ΔPAF −2·8) while FG rose (+2·6), and SBP and LDL returned towards their all-age values. By sex, attribution was very similar in men and women, with PAFs marginally lower in men and higher in women across most factors (mean |ΔPAF| ≤1·0 percentage points). These age and sex contrasts were broadly concordant across the C1 and C2 clusters, the exception being LDL at 70 years or older, where the two clusters moved in opposite directions (mean ΔPAF +1·1 percentage points in C1 and −0·9 in C2). Across subtypes, the fall in adiposity-related attribution at 70 years or older was largest for the haemorrhagic subtypes (mean ΔPAF −3·5 for intracerebral and −4·2 for subarachnoid haemorrhage, against −2·8 for total stroke), but the largest age-related contrast anywhere was for LDL in ischaemic stroke, which shifted from +9·1 percentage points at 50–69 years to −6·5 at 70 years or older (figures S12–S13). These subgroup analyses were descriptive and hypothesis-generating. For SBP they are directionally consistent with the cohort component, where the subdistribution hazard ratio fell from 1·43 below 70 years to 1·10 at 70 years or older; the two data sources otherwise highlight different exposures, since the age-related reversal of the eGFR association seen in the cohort is not mirrored by kidney dysfunction in the GBD estimates.

**4.6 Stroke-subtype composition clustering and its concordance**

For 2023, the proportions of IS, ICH, and SAH (of total stroke, by DALY rate and by incidence rate) were isometric log-ratio (ilr) transformed and clustered by Ward's method on Euclidean distances.^23^ The number of clusters was fixed to that of the risk-attribution clustering (k=2) for alignment, with the data-driven silhouette optimum also reported. Agreement with the risk-attribution clustering was assessed by a Mantel test (Spearman, 99 999 permutations) on the inter-country distance matrices (primary) and the adjusted Rand index with a permutation p value (99 999 permutations; auxiliary); the two dendrograms were compared in a tanglegram.

Clustering the ten states on the ilr-transformed subtype proportions of age-standardised DALY rates gave two clusters that were highly stable (section 4.8). SC1 (China, India, Pakistan, Tajikistan, Uzbekistan) was haemorrhage-heavy and SC2 (Belarus, Russia, Kazakhstan, Kyrgyzstan, Iran) ischaemia-predominant. The inter-country distances from this clustering were strongly correlated with those from the metabolic-attribution clustering on the Mantel test (r=0·68, p<0·0001), the primary measure of concordance; the adjusted Rand index, which requires exact co-assignment, was more modest (0·29, p=0·17), reflecting that Tajikistan and Uzbekistan grouped with the haemorrhage-heavy south and east Asian states on subtype composition while falling in attribution-C2. In the tanglegram this displacement appears as a change in label colour between the two sides rather than as a crossing connector, China, India, and Pakistan forming the consistent haemorrhagic core (figure S15).

The incidence-based (ASIR) subtype composition gave the same two-cluster partition as the DALY-based composition (figure S14, table S27); the two clusterings agreed exactly on all ten states, with the ASIR solution comparably stable (SC1 bootstrap Jaccard 0·90, SC2 0·88, versus 0·91 and 0·91 for ASDR). Its correlation with the attribution clustering was slightly lower (Mantel r=0·50, p=0·0024 vs r=0·68, p<0·0001), and the adjusted Rand index was necessarily identical (0·29; permutation p=0·17 in both), the two partitions being the same. We therefore treated the DALY-based composition as primary, since it weights subtypes by their disability and mortality burden, and the incidence-based composition as a confirmatory analysis.

**4.7 Spatial structure**

Geographical structure was assessed primarily with a Mantel test (Pearson correlation, 99 999 permutations) relating great-circle distances between national capitals to standardised feature distances,^12^ and secondarily with the join-count statistic for same-cluster adjacencies,^24^ tested by Monte Carlo permutation of cluster labels (99 999 permutations, one-sided for aggregation). The queen-contiguity adjacency of the ten states (shared land borders) comprised: China–Russia, China–Kazakhstan, China–Kyrgyzstan, China–Tajikistan, China–Pakistan, China–India, Russia–Kazakhstan, Russia–Belarus, Kazakhstan–Kyrgyzstan, Kazakhstan–Uzbekistan, Kyrgyzstan–Uzbekistan, Kyrgyzstan–Tajikistan, Tajikistan–Uzbekistan, Iran–Pakistan, and Pakistan–India.

For the burden-trajectory clustering, neither test was significant for total stroke (Mantel r=−0·18, p=0·77; join-count p=0·82), ICH (r=0·31, p=0·14; p=0·14), or SAH (r=−0·14, p=0·72; p=0·43); for IS the Mantel test was significant (r=0·37, p=0·03), though the join-count statistic was not (p=0·85; table S21).

For the metabolic-attribution clustering, geographical structure was significant for total stroke and every subtype on the Mantel test (r=0·39–0·61, p≤0·03), while the join-count statistic, of limited power given the few shared land borders, was non-significant throughout (p=0·17–0·29; table S26). For the subtype-composition clustering the corresponding statistics are reported in section 4.6, the concordance between that clustering and the attribution clustering being the quantity of interest there. These tests relate cluster membership to physical geography. The separate question of whether the clusters follow the GBD estimation hierarchy, which requires a hierarchy distance and a partial Mantel test, is addressed in section 2 and is not repeated here.

**4.8 Cluster stability**

Stability was assessed with fpc::clusterboot (B=1000)^25^ using non-parametric bootstrapping, jittering, and 80% subsetting, summarised by the mean Jaccard similarity per cluster (interpreted against thresholds of 0·60, 0·75, and 0·85); jittering added zero-mean gaussian noise whose principal directions match those of the feature matrix and whose standard deviation along each direction is the 0·05 quantile of the distances between neighbouring points projected on that direction. For the compositional clustering, internal rescaling was disabled to match the ilr coordinates. Country-level co-assignment matrices were derived separately, from 1000 resamples in which gaussian noise with a standard deviation of 5% of each feature's standard deviation was added to the matrix on which that clustering was performed—the standardised risk-factor matrix for the metabolic-attribution clustering and the untransformed isometric-log-ratio coordinates for the subtype-composition clustering.

For the total-stroke burden-trajectory clustering, bootstrap resampling, feature jittering, and subsetting confirmed the two multi-country clusters (bootstrap Jaccard 0·92 for Belarus, China, India, Iran, Tajikistan, and Uzbekistan, and 0·88 for Kazakhstan, Kyrgyzstan, and Russia); the single-country Pakistan cluster was less reproducible (0·63), as expected for a singleton (figure S4, table S20).

For the metabolic-attribution clustering, the two total-stroke clusters were highly reproducible (per-cluster bootstrap Jaccard 0·88 for C1 and 0·95 for C2, with jitter Jaccard 1·00 and subsetting Jaccard ≥0·98; figure S11, table S25).

For the subtype-composition clustering both clusters were highly stable (per-cluster bootstrap Jaccard 0·91 for both SC1 and SC2, with jitter and subset Jaccard ≥0·93; figure S16, table S27), and the incidence-based solution was comparably so (section 4.6).

**4.9 SDI–burden relationship and efficiency frontier**

The relationship between the age-standardised DALY rate and the SDI (2001–2023) was modelled with a generalised additive model (thin-plate spline, REML; mgcv),^26^ reporting effective degrees of freedom, the smooth-term significance, and deviance explained; departure from linearity was tested by a likelihood-ratio test between the smooth and a linear model, both refitted by maximum likelihood. An efficiency frontier was estimated by bootstrap data-envelopment analysis.^27^ Each country's relative inefficiency (relative distance to frontier, the gap between its ASDR and the frontier, as a proportion of the frontier) in 2023 and its 2001→2023 change were displayed.

Across all GBD locations, the association between the SDI and the age-standardised DALY rate was significantly nonlinear for total stroke and every subtype (penalised-spline nonlinearity p<0·0001 in the all-years generalised additive models; table S22), with deviance explained of 15·5–37·0%. For total stroke the relationship was inverted-U-shaped, with burden highest at intermediate SDI, and ischaemic stroke and intracerebral haemorrhage were concordant. Subarachnoid haemorrhage was also inverted-U-shaped but nearly symmetrically so, leaving no monotonic gradient: the linear correlation with SDI was null across all location-years (R=0·00) and small in the 2023 cross-section (R=−0·13; a linear-only model fitted for reference gives p=0·06, a quantity not tabulated for any outcome because it tests the linear component rather than the fitted smooth), so its burden is best described as peaking at intermediate SDI rather than as declining with development. In the bootstrap data-envelopment efficiency-frontier analysis (figures S7–S8; tables S22–S23), 2023 relative inefficiency ranged from Iran (closest to the frontier) to Russia (farthest); between 2001 and 2023, seven member states moved towards the frontier and three (India, Pakistan, and Tajikistan) moved away.

**5. CHARLS component**

**5.1 Data sources and eligibility**

Data were drawn from the Harmonized CHARLS dataset and the wave-1 venous blood subsample.^28^ Eligible participants were respondents alive and stroke-free at wave 1, aged ≥45 years, with non-missing wave-1 SBP, BMI, and age; blood-based exposures required participation in the wave-1 venous blood subsample and a fasting sample with non-missing glucose, creatinine, and LDL. Participants who were lost to follow-up after wave 2 and never re-contacted were excluded. Confounders with more than 20% missingness were dropped a priori. No comparable individual-level cohort was available for the other member states. The Longitudinal Ageing Study in India,^29^ the Russia Longitudinal Monitoring Survey–Higher School of Economics (RLMS-HSE),^30^ and the WHO STEPS surveys in the central Asian states were each considered but could not be used,^31^ because they lacked either prospective follow-up or the full set of baseline metabolic measurements required here; the cohort component is therefore confined to China.

**5.2 Baseline characteristics**

Baseline characteristics were compared across the three outcome groups (no event, stroke, competing death) using analysis of variance or chi-square/Fisher's exact tests, with SMDs; pairwise comparisons are tabulated in table S28.

**5.3 Exposures and kidney function**

SBP was the mean of the second and third wave-1 readings, to reduce the white-coat effect (if only one of these was available, that reading was used); BMI, FG, LDL, and creatinine were taken from wave 1 (units: mmHg; kg/m²; mg/dL; mg/dL). eGFR was computed with the race-free CKD-EPI 2021 creatinine equation;^32^ for a sensitivity analysis, eGFR was recomputed with the CKD-EPI 2021 creatinine–cystatin C equation in participants with a cystatin C measurement. Each exposure was standardised to a mean of 0 and SD of 1 for modelling; values outside physiologically plausible ranges were excluded (SBP 70–250 mmHg; FG 40–500 mg/dL; LDL 20–400 mg/dL; eGFR 15–200 mL/min/1·73m²; BMI 12–60 kg/m²).

**5.4 Outcome, competing event, and follow-up time**

Incident stroke was ascertained from self-reported physician diagnosis at waves 2–4. The competing event was non-stroke death during follow-up. Each participant was classified as having stroke (if a stroke was reported before any death), a competing death (if death was reported without a prior stroke), or censored (if neither occurred). Because events were observed only at biennial waves (interval censoring), event times were imputed as midpoints: for stroke, the midpoint between the wave at which stroke was first reported and the last preceding wave at which the participant was alive and interviewed; for competing death, the midpoint between the wave at which death was reported and the last preceding wave at which the participant was alive and interviewed. Censored participants contributed follow-up to their last interview at which they were alive. Follow-up time was measured from the wave-1 interview to the event or censoring time.

**5.5 Covariates**

Models were adjusted for age (continuous), sex, marital status, education, urban or rural residence, smoking, alcohol use, heart disease, and depression (CES-D >10);^33^ Model 3 additionally adjusted for antihypertensive, antidiabetic, and lipid-lowering medication use. Coding is given in table S9. Marital status (four categories) and education (three categories) were entered as indicator variables referenced to the most common category. The never-married category was sparse in some strata (six participants aged 70 years or older; one woman), but every model converged and the stratum-specific exposure estimates and confidence intervals were stable, so categories were not collapsed, in order to keep the covariate specification identical across all CHARLS analyses. All models were fitted on complete cases: Models 1 and 2 in 7896 participants (565 incident strokes) and Model 3, which additionally requires medication data, in 7710 (556). Refitting Model 2 in the Model 3 sample left the estimates essentially unchanged (SBP 1·37, BMI 1·21, FG 1·10, LDL 1·03, eGFR 1·02), so the attenuation seen in Model 3 reflects medication adjustment rather than the change in sample.

**5.6 Competing-risk modelling**

Cumulative incidence functions for stroke and competing death were estimated overall and, for descriptive comparison, stratified by clinical cut-offs (SBP ≥140 mmHg, BMI ≥28 kg/m² [Chinese criterion], FG ≥126 mg/dL, LDL ≥160 mg/dL, eGFR <60 mL/min/1·73m²) with Gray's test. SHRs were estimated with the Fine-Gray model (cmprsk::crr; failure code stroke, censoring code 0) for Models 1–3.^34^

**5.7 Nonlinearity and proportional subdistribution hazards**

Nonlinearity was assessed on the subdistribution hazard scale. Fine-Gray weighted risk sets were constructed with the finegray function of the survival package, which retains participants who experience the competing event in the risk set with inverse-probability-of-censoring weights; a weighted Cox model with robust variance and Breslow tie handling was then fitted to these data, a specification algebraically equivalent to the crr estimator used for Models 1–3. Equivalence was verified by refitting Model 2 with linear exposure terms in the weighted data; SHRs agreed with the crr estimates to within 0·02.

Each exposure was entered as a four-knot restricted cubic spline with knots at the 5th, 35th, 65th, and 95th percentiles,^35^ adjusted for the covariate set. Nonlinearity was tested by a joint Wald test of the two nonlinear spline coefficients (2 df) using the robust covariance matrix, and the overall association by a joint test of all three spline coefficients (3 df); FDR correction was applied to the nonlinearity tests across the five exposures. Because the reference value determines only the vertical position of the curve and not any test statistic or contrast, reference values were prespecified rather than data-derived, and were chosen to be clinically interpretable and, where the GBD TMREL is defined on the same continuous scale, to fall within it (BMI 22 kg/m², within the TMREL window of 20–25; FG 90 mg/dL, within 86·5–97·3; SBP 115 mmHg, within 110–115). For LDL the GBD TMREL (27–50 mg/dL) lies below the 10th percentile of the cohort distribution, and for kidney function the GBD exposure is defined categorically, on eGFR stages together with albuminuria, rather than on a continuous eGFR scale, whereas the cohort measured eGFR alone; reference values of 100 mg/dL and 100 mL/min per 1·73 m² were used instead and no TMREL alignment was possible. Curves and confidence intervals were obtained by contrasting the spline basis at each grid value against the basis at the reference value, with standard errors from the delta method; contrasts between pairs of exposure values, which are invariant to the reference, were computed in the same way. Two sensitivity analyses were done for the nonlinearity assessment: refitting each spline with the other four exposures included as linear terms (the Model 2 specification), and refitting on the cause-specific hazard scale with competing deaths censored, to establish whether the shapes were sensitive to the treatment of competing risk.

The proportional subdistribution hazards assumption was tested by adding exposure-by-log-time interaction terms to the Fine-Gray model; for exposures violating the assumption (SBP and BMI), time-specific SHRs were derived from the fitted main and interaction coefficients.

Four-knot restricted cubic splines fitted on the subdistribution hazard scale identified significant departures from linearity for FG (nonlinearity χ²=11·22, 2 df, p=0·0037; FDR-corrected p=0·0093) and BMI (χ²=12·28, p=0·0022; FDR p=0·0093), and no significant departure for SBP (p=0·36), LDL (p=0·25), or eGFR (p=0·08; FDR p=0·13). The overall three-degree-of-freedom test of association was significant for SBP, FG, and BMI (p<0·0001 for each) but not for LDL (p=0·15) or eGFR (p=0·13), consistent with the null estimates for these two exposures in Model 2; shape is therefore described below only where an overall association was present (figure S19, tables S30–S31).

Both departures survived a change of hazard scale: refitting all five splines on the cause-specific hazard scale with competing deaths censored reproduced them (FG nonlinearity p=0·01, FDR-corrected p=0·03; BMI p=0·0079, FDR p=0·03; p≥0·25 for the other three), so neither shape is an artefact of retaining competing deaths in the subdistribution risk set. The cause-specific refits did not mutually adjust the exposures. In the separate mutually adjusted refits, both departures were attenuated: with the other four exposures included as linear terms (Model 2 specification), BMI retained a nominal departure that did not survive correction (χ²=7·01, p=0·03; FDR p=0·12) and FG did not (χ²=4·70, p=0·10; FDR p=0·16), indicating that part of the curvature is shared with the other metabolic exposures. The shapes are therefore reported from the primary single-exposure specification.

Relative to a reference of 22 kg/m2, the BMI curve rose monotonically and with a decreasing slope: the SHR was 0·57 (95% CI 0·42–0·79) at 18 kg/m2, 0·76 (0·68–0·85) at 20, 1·37 (1·16–1·62) at 25, 1·60 (1·29–2·00) at 28, and 1·78 (1·41–2·26) at 32. The slope was about 12% per kg/m2 between 20 and 25 kg/m2 and about 5% per kg/m2 between 25 and 28. Notably, risk was lowest at the bottom of the BMI range rather than at both extremes, so no J-shaped excess at low BMI was evident. Participants who died were markedly leaner than those who did not (mean BMI 22·2 vs 23·5), so the handling of competing deaths could in principle have shaped this curve; the primary specification retains these participants in the risk set with inverse-probability-of-censoring weights, and censoring them instead, in the cause-specific refit above, left the shape unchanged. Reverse causation from subclinical illness depressing baseline weight, and the sparsity of observations below 19 kg/m2 (the 10th percentile), remain plausible contributors and are not excluded by this analysis.

Relative to a reference of 90 mg/dL, the FG curve showed the same pattern: SHR 0·76 (0·60–0·97) at 80 mg/dL, 1·17 (1·05–1·31) at 97, 1·40 (1·19–1·64) at 110, 1·57 (1·25–1·97) at 126, and 1·71 (1·34–2·18) at 160, corresponding to about 18% per 10 mg/dL between 90 and 110 and about 8% per 10 mg/dL between 110 and 126. Within the GBD TMREL window for FG (86·5–97·3 mg/dL) the fitted hazard differed 1·17-fold (1·05–1·31) between 90 and 97 mg/dL; within the corresponding BMI window (20–25 kg/m²) it differed 1·80-fold (1·48–2·18). SBP was compatible with a linear effect, with SHRs of 1·74 (1·40–2·15) at 140 mmHg and 2·95 (2·28–3·84) at 180 mmHg relative to 115 mmHg. The eGFR curve was inverted-U in shape, with no excess risk at low values and a decline above 100 mL/min per 1·73 m² (SHR 0·56, 0·31–1·01, at 120); this did not reach significance and lies above the 90th percentile of the cohort distribution, so it is reported descriptively only.

These within-window gradients bear directly on how attributable fractions are constructed. GBD assigns zero attributable burden inside the TMREL window—20–25 kg/m² for BMI and 86·5–97·3 mg/dL for FG—yet across those same intervals the fitted subdistribution hazard differed 1·80-fold and 1·17-fold in this cohort. The TMREL counterfactual does not register variation inside its window, so this observed gradient supplies a structural explanation for the BMI ranking discrepancy. The cohort subdistribution hazard is not the pooled relative risk used by GBD: these results cannot establish how much causal attributable burden, if any, the counterfactual omits. The primary nonlinearity survived FDR correction, but the mutually adjusted nonlinearity did not (section 5.7).

Whether such a gradient carries any consequence for the classification of a factor is a separate question, settled by the decision rules and not by the gradient itself. Fasting glucose shows a gradient inside its window as body-mass index does, yet the population and individual streams place it at adjacent positions, fourth and third; a one-position difference falls below the two-position threshold at which a divergence between those two streams is recorded, so mechanism M1 has no divergence to explain and is not applicable. The divergence that does exist for fasting glucose is with the genetic stream, which M1 does not address. The derivation for each factor is set out in section 3.2 and table S17.

Only SBP (p<0·0001) and BMI (p=0·0001) violated the proportional subdistribution hazards assumption, both with negative log-time coefficients indicating attenuation over follow-up (table S32); FG, LDL, and eGFR were compatible with proportional hazards. Time-specific SHRs declined monotonically for both exposures (SBP: 3·04, 2·21, 1·83, 1·61, 1·45, 1·33, and 1·24 at years 1–7; BMI: 2·46, 1·88, 1·60, 1·43, 1·31, 1·22, and 1·15) without crossing the null, indicating a diminishing but persistently harmful effect. Because events were observed only at biennial waves and event times imputed at wave midpoints, the observed event times take few distinct values and the year-1 estimates lie at the boundary of the data; the year-1 values should be read as an extrapolation of the fitted log-time trend rather than as an estimate anchored in observed events, and the overall magnitude of attenuation is the interpretable quantity.

**5.8 Subgroup analyses**

Model 2 was refitted within subgroups defined by age (<70 vs ≥70 years), sex, and residence. Exposures were standardised on the whole-sample SD so that stratum-specific SHRs share a common scale and are comparable with the interaction tests; the within-stratum raw SDs are given in the lower panel of table S33. Interactions were tested with exposure-by-subgroup product terms in the full sample, and binary covariates showing complete separation within a subgroup were dropped. These analyses were prespecified but exploratory and were not corrected for multiplicity.

Age modified the SBP association most strongly: the SHR fell from 1·43 (1·31–1·55) below 70 years to 1·10 (0·89–1·36) at 70 years or older (p_interaction_=0·0047), mirroring the attenuation of the SBP subdistribution hazard over follow-up (table S32). The SBP SD was larger in the older stratum (23·1 vs 20·4 mmHg), so on the common scale the attenuation is if anything understated and is not an artefact of standardisation. The eGFR association was null in both age strata but differed in direction between them—0·99 (0·89–1·09) below 70 years and 1·12 (0·89–1·41) at 70 years or older (p_interaction_=0·02)—so that no association was seen in the pooled Model 2; because neither stratum-specific estimate was significant, this age-related difference is hypothesis-generating. Sex modified the SBP (p=0·04) and BMI (p=0·03) associations, both stronger in men, though all stratum-specific estimates remained directionally concordant; residence modified none (all p_interaction_≥0·18).

**5.9 Statistical inference and multiplicity**

The primary Fine-Gray models were prespecified, and the five exposures were mutually adjusted; inference focused on SHRs with 95% CIs. We therefore did not apply multiplicity correction to the five exposures of the primary models. False-discovery-rate correction was reserved for the family of exploratory nonlinearity tests, and subgroup interaction tests were interpreted as exploratory and hypothesis-generating. Taken across the study, FDR correction was therefore applied to two defined families only—the five spline nonlinearity tests, and the five exposures within each MR outcome (see below)—while clustering, spatial, subgroup, and secondary attribution analyses were interpreted as descriptive or exploratory and were not corrected.

**5.10 Sensitivity analyses**

Three sensitivity analyses were done: (1) a cause-specific Cox model with competing deaths censored (Models 1–3); (2) Fine-Gray models (Models 1–3) using the mean of wave 1 and 3 exposures to reduce regression dilution, restricted to eligible participants who were also stroke-free and reinterviewed at wave 3 with complete wave-3 measurements, with follow-up starting at wave 3 and covariates taken at wave 1 apart from age, which was age at wave 3; and (3) within the subsample with both creatinine and cystatin C, re-estimation of the kidney-function association using the CKD-EPI 2021 creatinine–cystatin C eGFR, comparing the two eGFR distributions (paired t test, Wilcoxon signed-rank test, SMD) and the two sets of SHRs (Z test).

First, cause-specific Cox models with competing deaths censored reproduced the primary associations at every adjustment level, with SBP, BMI, and FG remaining significant and LDL and eGFR null (table S34). Second, in the 4284 participants of the two-wave sample, averaging exposures over waves 1 and 3 preserved the SBP association and, in Model 2, the BMI association, which became marginal after medication adjustment (Model 3 SHR 1·13 [1·00–1·27]; 4134 with complete covariate data), and reduced the FG association to non-significant (Model 2 SHR 1·06 [0·95–1·18]; Model 3 0·99 [0·88–1·12]), identifying FG as the exposure most sensitive to measurement, whereas the LDL and eGFR nulls were unchanged (table S35). Third, within the 6109 participants with both creatinine and cystatin C, the two eGFR equations differed appreciably in level (mean 95·0 vs 88·2 mL/min per 1·73 m²; mean difference 6·81, 95% CI 6·53–7·09; SMD 0·42; Pearson r=0·79), yet the creatinine–cystatin C eGFR gave the same null overall association (Model 3 SHR 1·03 [0·92–1·15]; 5876 with complete covariate data); the age-related difference also persisted with the combined equation (≥70 years 1·20 vs <70 years 0·97), indicating it does not arise from creatinine-based overestimation of eGFR in sarcopenic older adults (table S36).

**5.11 Auxiliary variable-importance analysis**

To check that the ranking and functional form of the associations were not artefacts of the Fine-Gray specification, two contrasting model-agnostic learners were fitted. A gradient-boosted tree ensemble (XGBoost)^36^ was trained to discriminate incident stroke (competing deaths excluded) using the five exposures and the covariates, with a 70/30 train–test split, 5-fold cross-validated tuning, class-weighting for imbalance, and marital status and education entered as indicator variables as in the primary models; importance was summarised by mean absolute SHAP values.^37^ A penalised logistic (lasso) model was fitted to the same participants and variables, with importance summarised by absolute standardised coefficients.^38^ These analyses were interpreted for aetiological ranking, not prediction, and served only as a corroborative check that did not inform the primary inference.

Both learners placed SBP first among the five exposures and reproduced the leading order of the primary analysis (SBP > BMI > FG), but the two least important exposures were not separated consistently: SHAP ranked eGFR above LDL, whereas the lasso shrank both to zero. Discrimination was modest and near-identical for the two models in the test set (AUC 0·66 for both; DeLong p=0·56), showing no advantage of the flexible ensemble. SHAP dependence plots reproduced the threshold-type, saturating shapes for BMI and FG independently identified by the spline analysis. SHAP summary (beeswarm) plots add no aetiological information and are not shown (figure S20).

**6. Multi-ancestry MR component**

**6.1 Data sources and ancestry mapping**

Two-sample MR was done in three ancestries mapped to SCO members of predominantly that ancestry: European (Russia, Belarus), east Asian (China), and south Asian (India, Pakistan). Five member states, falling into two ancestry groups, could not be instrumented: no suitable exposure or outcome data were available for central Asian populations (Kazakhstan, Kyrgyzstan, Tajikistan, Uzbekistan) or for Iran. Exposure and outcome GWAS, with sample sizes and accession identifiers, are listed in tables S7–S8. European exposures were drawn from a within-family GWAS consortium (LDL, SBP, BMI, eGFR),^18^ from which we used the population rather than the within-sibship estimates because the within-sibship subsamples yielded too few genome-wide significant variants to instrument the exposures; these estimates are therefore not adjusted for within-family confounding, as is also true of the MAGIC FG estimates.^39^ Outcomes were from FinnGen (total stroke)^40^ and a UK Biobank–FinnGen meta-analysis (IS, ICH, SAH).^41^ East Asian exposures were from TPMI (Taiwan; FG, LDL, SBP, BMI)^42^ and Biobank Japan (eGFR),^43^ with outcomes from the China Kadoorie Biobank;^44^ because the exposure and outcome samples are from separate populations, there is no sample overlap. The CKB phenotype used as the east Asian total-stroke outcome corresponds to ICD-10 I64 (stroke, not specified as haemorrhage or infarction) and therefore represents unclassified stroke rather than all stroke, which explains its small case count (876) relative to the CKB subtype outcomes. FinnGen C_STROKE and the GIGASTROKE any-stroke phenotype are, by contrast, broad composite definitions. Total-stroke estimates are consequently not comparable across ancestries and were interpreted within ancestry only; all cross-ancestry inference rests on the subtype outcomes. South Asian exposures were from Pan-UK Biobank (central/south Asian)^45^ and MAGIC (FG),^39^ with total stroke and IS outcomes from GIGASTROKE;^46^ GIGASTROKE includes some Pan-UK Biobank participants, giving possible modest overlap, which we note as a limitation.

**6.2 Instrument construction**

Instruments were independent variants associated with each exposure at p<5×10−8 (relaxed to p<1×10−5 for south Asian ancestry owing to the small sample), LD-clumped with an ancestry-matched 1000 Genomes reference panel (r²<0·01 and a 10 000 kb window for univariable MR). For MVMR the instruments selected for each exposure were clumped at r²<0·001 within that exposure; in east Asian ancestry the sets were then pooled across the five exposures and re-clumped at r²<0·005 to remove residual correlation between variants chosen for different exposures.^5^ The per-variant F statistic was computed as (β/SE)², and only variants with F≥10 were retained. TPMI summary statistics in SAIGE format were mapped from chromosome–position to rsID using SNPnexus;^47^ China Kadoorie Biobank and GIGASTROKE variants without rsIDs were matched to exposures by chromosome and position. No LD proxy substitution was used in any ancestry: variants absent from the outcome dataset were dropped rather than replaced by a correlated surrogate. Exposure and outcome data were harmonised with inference of the positive strand from allele frequencies (harmonisation action 2),^48^ and at least three variants were required for estimation.

**6.3 Univariable MR and sensitivity analyses**

The primary estimator was IVW MR.^49^ Sensitivity analyses comprised MR-Egger and the weighted median (robust to differing assumptions about invalid instruments).^50,51^ Between-instrument heterogeneity was assessed by Cochran's Q, directional horizontal pleiotropy by the MR-Egger intercept, and the influence of individual variants by leave-one-out analysis.

**6.4 Multivariable MR**

MVMR modelled all five exposures jointly to estimate each exposure's direct effect. The primary estimator was MVMR-IVW (MendelianRandomization), with MVMR-Egger as a sensitivity analysis (its intercept testing directional pleiotropy). Instrument strength was quantified by the conditional F statistic (instruments regarded as weak below 5 and borderline between 5 and 10)^52,53^ and residual pleiotropy by the conditional Q statistic (MVMR), assuming zero genetic covariance between exposures, a conservative simplification we note as a limitation. Estimates for exposures with borderline conditional strength are reported but interpreted alongside their univariable counterparts rather than in isolation. Multivariable IVW p values were false-discovery-rate corrected across the five exposures within each outcome, as in the univariable analysis; MVMR-Egger, used only as a sensitivity estimator, was not corrected. Exposure-matrix column order was verified against the source identifiers before estimation.

**6.5 European ancestry**

Instruments were strong for all five exposures (mean F 40–129; table S37). In univariable IVW analysis, SBP was robustly associated with IS (OR 1·047 [1·033–1·061]; FDR p<0·0001) and with the composite total-stroke phenotype (1·042 [1·027–1·058]; FDR p<0·0001), and was nominally associated with SAH before correction, the ICH estimate being borderline (p=0·06). LDL was associated with IS (1·093 [1·034–1·155]; FDR p=0·0042) and total stroke (1·175 [1·098–1·257]; FDR p<0·0001) but was null for ICH (0·989) and directionally protective but non-significant for SAH (0·872 [0·760–1·002]; FDR p=0·09). BMI was associated with IS only (1·030 [1·008–1·054]; FDR p=0·01). FG and eGFR showed no association with any outcome; the nominal protective FG–SAH estimate (0·636) did not survive correction. MR-Egger and weighted-median estimates were directionally concordant for the significant associations (table S39), and Egger intercepts showed no directional pleiotropy for the SBP–IS, LDL–IS, or BMI–IS relationships (all p>0·28; table S39). Heterogeneity was present for the SBP instruments with all three subtypes (Q p<0·0001 for IS and SAH and p=0·0010 for ICH) and for LDL with IS and ICH (both p=0·02), as expected for highly polygenic exposures, and was accommodated by the random-effects IVW estimator.

In MVMR (141 instruments; 138 for total stroke), the LDL–IS association persisted after mutual adjustment (1·091 [1·018–1·170]; FDR p=0·03; conditional F=35·6), whereas the BMI–IS association attenuated to the null (1·002 [0·974–1·031]), indicating mediation or confounding by the other metabolic traits. SBP was associated with all three subtypes, reaching significance for ICH (FDR p=0·0047) and SAH (FDR p=0·0003) with the larger pooled instrument set, though its conditional F (5·82) indicates borderline conditional instrument strength. FG and eGFR remained null. For total stroke the multivariable model showed substantial heterogeneity (Q p<0·0001) without evidence of directional pleiotropy (Egger intercept p=0·08), and MVMR-Egger estimates for LDL (1·224 [1·120–1·338]) and SBP (1·034) were consistent with the IVW results (table S40).

The multivariable attenuation of the BMI–ischaemic stroke estimate to the null is not in conflict with the cohort, in which the same mutual adjustment leaves body-mass index the second strongest exposure (subdistribution hazard ratio 1·22, 1·14–1·30). The two analyses do not estimate the same quantity: the instruments capture genetically determined lifetime differences in body-mass index and were modelled linearly, so an effect operating through downstream metabolic traits is absorbed by those traits once they are conditioned on, whereas the cohort captures measured body-mass index at a single point and finds a markedly nonlinear association (section 5.7). The classification that follows from this is derived in section 3.2 and table S17; no further reading is offered here.

**6.6 East Asian ancestry**

Exposure instruments were derived from Taiwan Precision Medicine Initiative (TPMI) data for SBP, LDL, BMI, and FG and from Biobank Japan for eGFR, and were strong for all five exposures (mean F 51–112; table S37). The total-stroke outcome was the CKB phenotype for stroke not specified as haemorrhage or infarction (ICD-10 I64) and is therefore not comparable with the composite total-stroke phenotypes used in the other ancestries; it is reported within ancestry only.

In univariable IVW analysis, SBP was associated with IS, ICH, SAH, and total stroke (all FDR-corrected p<0·05; table S38), a more uniform subtype pattern than in European ancestry. The LDL–IS estimate was positive under all three estimators (IVW OR 1·106 [0·987–1·240], p=0·08; MR-Egger 1·254 [1·040–1·511], p=0·02; weighted median 1·137 [0·974–1·328], p=0·10; table S39), but the primary IVW estimate did not reach significance after correction (FDR-corrected p=0·14). Whether that null is informative was assessed quantitatively rather than by comparing instrument strength, which governs the exposure side and not the precision of a ratio estimate. The smallest odds ratio this analysis could have detected with 80% power was 1·18, above the point estimate of 1·106, and the interval does not exclude an effect as large as 1·240. The analysis was therefore not able to distinguish a smaller causal effect of LDL on ischaemic stroke in east Asian populations from insufficient precision, and the result is reported as uninformative rather than as evidence of a weaker effect. BMI showed nominal positive estimates for IS and ICH that did not survive correction, and FG, eGFR, and the remaining subtype associations were null.

MR-Egger and weighted-median estimates were directionally concordant with IVW for every SBP association. MR-Egger was, however, considerably less precise in this ancestry, and for SBP and IS returned a substantially larger point estimate than IVW (8·02 [2·36–27·23] vs 2·94 [2·14–4·04]) alongside a borderline intercept (p=0·11). The weighted-median estimate, which is robust to a minority of invalid instruments, closely matched the IVW estimate (2·87 [1·97–4·19]), so the SBP associations are unlikely to be artefacts of directional pleiotropy; the remaining intercepts gave no indication of it (table S39). Instrument heterogeneity was substantial for the SBP and eGFR instruments with the ischaemic and intracerebral-haemorrhage outcomes (all Q p<0·01, most markedly for eGFR with ICH, p<0·0001) and for FG with intracerebral haemorrhage (p=0·0032); it was accommodated by the random-effects estimator.

In MVMR, SBP remained associated with all subtypes (IS and ICH FDR-corrected p<0·0001, SAH p=0·03) whereas LDL, BMI, FG, and eGFR showed no independent effect; conditional instrument strength was borderline for SBP (F=7·5) and BMI (F=8·6) and adequate for the others. Multivariable pleiotropy tests were null for all outcomes except modest heterogeneity for ICH (Q p=0·0049; table S40).

One estimate is noted for completeness because it recurs across ancestries. Fasting glucose was inversely associated with subarachnoid haemorrhage in east Asian ancestry (0·572, 0·348–0·941; uncorrected p=0·028, FDR-corrected p=0·07) and in the same direction in European ancestry (0·636, 0·447–0·906; uncorrected p=0·012, FDR-corrected p=0·061). Neither survived correction within its outcome, neither enters the genetic-stream ordering, and the classification of fasting glucose is unchanged; the pair is recorded here rather than in the main text so that a directionally consistent result is not silently omitted.

**6.7 South Asian ancestry**

South Asian instruments were drawn from the Pan-UK Biobank Central and south Asian analysis for SBP, LDL, BMI, and eGFR, and from GCST90002234 for FG. Outcome data were the GIGASTROKE south Asian analyses. No ICH or SAH GWAS data were available in this ancestry, so analyses were restricted to IS and the composite total-stroke phenotype. That composite is moreover almost entirely ischaemic (3462 of 3640 cases), so it carries little information beyond the IS analysis and is not interpreted separately. Together with the broad composite used in European ancestry and the unclassified-only phenotype used in east Asian ancestry, this illustrates why total stroke cannot be compared across ancestries: the three phenotypes differ not in precision but in what they count.

Instruments were substantially weaker than in the other ancestries, with 5–13 variants surviving harmonisation per exposure (table S38) and mean F statistics of 21–41 (table S37), against 19–85 variants and F 40–129 in European and 31–61 variants and F 51–112 in east Asian. No exposure was associated with IS after FDR correction (lowest FDR-corrected p=0·31; table S38). Point estimates for BMI (1·390 [0·960–2·012]) and SBP (1·362 [0·920–2·016]) were directionally concordant with the European and east Asian results, whereas the LDL estimate was directionally opposite (0·842 [0·536–1·322]) but rested on five instruments and was accompanied by an implausible MR-Egger estimate (0·230 [0·053–0·993]), consistent with instrument noise rather than a protective effect. There was no evidence of heterogeneity or directional pleiotropy for any exposure (all Q p>0·20; all Egger intercept p>0·16).

Unlike the east Asian analyses, in which the same outcome data yielded a strong SBP signal, the south Asian data did not detect an association even for SBP, the exposure most consistently associated with stroke in the other two ancestries. The confidence intervals therefore reflect limited statistical information rather than evidence against an effect, and the south Asian estimates are reported for completeness without contributing to the genetic-stream ordering. Multivariable MR was attempted but the conditional F statistic was below 5 for four of the five exposures and borderline for the fifth, indicating insufficient conditional instrument strength; those estimates are provided as source data and are not interpreted.

**6.8 Cross-ancestry pattern and multiplicity**

Across the three ancestries the findings were consistent in direction but unequal in precision. SBP was the only exposure associated with stroke in more than one ancestry: with every subtype in east Asian ancestry and, in multivariable analysis, with every subtype in European ancestry. LDL was associated with IS in European ancestry and was directionally concordant but weaker and estimator-dependent in east Asian ancestry, and was associated with neither haemorrhagic subtype in either ancestry. BMI showed a univariable association with IS in European ancestry that did not survive mutual adjustment, and nominal estimates for IS and ICH in east Asian ancestry that did not survive correction; FG and eGFR were null throughout. The south Asian estimates were too imprecise to contribute to any of these comparisons. All cross-ancestry statements concern the direction of effect and statistical significance, not its magnitude, for the reasons set out in section 3.4.

Within each outcome, inverse-variance-weighted and multivariable inverse-variance-weighted p values were corrected across the five exposures by the Benjamini-Hochberg false-discovery-rate method. South Asian analyses were regarded as exploratory.

**7. Software and packages**

All analyses were done in R version 4.5.0. Data management used tidyverse and data.table. The GBD component used cluster, dendextend, fpc, and mclust (hierarchical clustering, stability, and agreement), compositions (isometric log-ratio transformation), vegan, spdep, and geosphere (Mantel, join-count, and spatial weights), ComplexHeatmap and circlize (heatmaps), mgcv (generalised additive models), and sf with rnaturalearth (maps). The CHARLS component used survival (Fine-Gray weighted risk sets via finegray, weighted Cox fits, and cause-specific models), cmprsk (crr), and Hmisc (restricted cubic spline bases), tableone and rstatix (baseline comparisons), and xgboost with SHAPforxgboost, shapviz, glmnet, caret, and pROC (the auxiliary gradient-boosted and penalised models). The multi-ancestry MR component used TwoSampleMR, MVMR, and MendelianRandomization (instrument extraction, harmonisation, univariable and multivariable estimation, and diagnostics). Figures used ggplot2, patchwork, cowplot, forestploter, and ggrepel. The authors' analysis code was additionally checked for internal consistency and correctness with the assistance of an AI tool (Claude; Anthropic), as described in the declaration at the end of the main text; all resulting revisions were made and re-run by the authors.

**8. Ethics and sex reporting**

Ethics approvals and participant consent for CHARLS are stated in the Methods of the main text. The GBD and MR analyses used publicly available aggregate data and required no additional approval. Sex was self-reported in CHARLS and analysed as a binary variable (female or male); in GBD and the genetic datasets, sex was defined as in the source data.

3.	Institute for Health Metrics and Evaluation. Global Burden of Disease Study 2023 (GBD 2023) cause, REI, and location hierarchies [dataset]. Seattle, WA: IHME, 2025.

4.	National Academies of Sciences, Engineering, and Medicine. Using population descriptors in genetics and genomics research. Washington, DC: National Academies Press, 2023.

10.	South A, Michael S, Massicotte P. rnaturalearthdata: world vector map data from Natural Earth used in 'rnaturalearth'. 2024.

11.	Fiedler M. Algebraic connectivity of graphs. *Czechoslovak Math J* 1973; **23**(2): 298-305.

12.	Mantel N. The detection of disease clustering and a generalized regression approach. *Cancer Res* 1967; **27**: 209–20.

13.	Smouse PE, Long JC, Sokal RR. Multiple regression and correlation extensions of the Mantel test of matrix correspondence. *Syst Zool* 1986; **35**(4): 627-32.

14.	Legendre P, Legendre L. Numerical ecology. 3rd edn. Amsterdam: Elsevier, 2012.

24.	Cliff AD, Ord JK. Spatial autocorrelation. London: Pion, 1973.

35.	Harrell FE. Regression modeling strategies: with applications to linear models, logistic and ordinal regression, and survival analysis. 2nd edn. Cham: Springer, 2015.

**Legends**

**Figure S1: Temporal burden trajectories of ischaemic stroke in the ten SCO member states, 2001–2023**

As figure 2, for ischaemic stroke. (A) Annual rates per 100 000 by trajectory cluster: rows are ASIR, ASMR, and ASDR and columns are the four clusters. Coloured lines are individual member states and the dashed black line is the global rate. The y-axis scale is shared across clusters within a row and differs between rows. (B) Dendrogram of the same clustering, obtained by Ward's linkage on the five standardised temporal features defined in figure 2; dashed boxes delimit the clusters, each labelled beneath, and height is the Ward merge distance. Cluster 1 comprises India and Pakistan; cluster 2 Belarus, Kazakhstan, and Kyrgyzstan; cluster 3 Russia and Tajikistan; and cluster 4 China, Iran, and Uzbekistan. The tip order of the dendrogram follows the clustering and not the left-to-right order of the columns in (A). The number of clusters was chosen by maximum mean silhouette width separately for each outcome and therefore differs between the subtypes and from the total-stroke clustering of figure 2; silhouette widths and per-cluster bootstrap stability are given in table S20. Cluster numbers are labels: they do not order the clusters by burden and are not comparable between subtypes.

**Abbreviations**

ASDR=age-standardised DALY rate. ASIR=age-standardised incidence rate. ASMR=age-standardised mortality rate. DALY=disability-adjusted life-year. SCO=Shanghai Cooperation Organisation.

**Figure S2: Temporal burden trajectories of intracerebral haemorrhage in the ten SCO member states, 2001–2023**

As figure S1, for intracerebral haemorrhage, with three clusters. Cluster 1 comprises Belarus, Kazakhstan, Kyrgyzstan, Russia, Tajikistan, and Uzbekistan; cluster 2 China and Pakistan; and cluster 3 India and Iran.

**Abbreviations**

ASDR=age-standardised DALY rate. ASIR=age-standardised incidence rate. ASMR=age-standardised mortality rate. DALY=disability-adjusted life-year. SCO=Shanghai Cooperation Organisation.

**Figure S3: Temporal burden trajectories of subarachnoid haemorrhage in the ten SCO member states, 2001–2023**

As figure S1, for subarachnoid haemorrhage, with four clusters. Cluster 1 comprises India and Uzbekistan; cluster 2 Belarus, Kazakhstan, and Kyrgyzstan; cluster 3 Russia alone; and cluster 4 China, Iran, Pakistan, and Tajikistan.

**Abbreviations**

ASDR=age-standardised DALY rate. ASIR=age-standardised incidence rate. ASMR=age-standardised mortality rate. DALY=disability-adjusted life-year. SCO=Shanghai Cooperation Organisation.

**Figure S4: Stability of the total-stroke burden-trajectory clustering**

(A) Country co-assignment frequencies at k=3: the proportion of 1000 resamples—each obtained by adding gaussian noise with a standard deviation of 5% of each feature's standard deviation to the standardised feature matrix and reclustering—in which a given pair of member states fell in the same cluster (1·00=always in the same cluster, 0·00=never). States are grouped by their cluster assignment from figure 2—cluster 1 (Belarus, China, India, Iran, Tajikistan, Uzbekistan), cluster 2 (Kazakhstan, Kyrgyzstan, Russia), and cluster 3 (Pakistan)—and ordered alphabetically within each cluster; the black lines mark the cluster boundaries.

(B) Mean Jaccard similarity of each cluster to its closest counterpart in resampled data, at k=2, 3, and 4, under three resampling schemes implemented in fpc::clusterboot—non-parametric bootstrapping, jittering, and 80% subsetting—with 1000 replicates each; the jittering here is scaled to the local spacing of the data along each principal direction and is therefore not the fixed perturbation used in (A) (appendix). Dashed lines mark the conventional thresholds of 0·60, 0·75, and 0·85, at or above which a cluster is regarded as indicating a pattern, as stable, and as highly stable, respectively; higher values indicate a more reproducible cluster. Cluster numbers are internal to each value of k and correspond to those of figure 2 only in the k=3 panel, which is the solution selected by maximum mean silhouette width and reported throughout.

Both perturbation schemes recovered the three clusters in nearly every resample, so every co-assignment frequency in (A) is 0 or 1; the lower values under bootstrapping and subsetting in (B) reflect the information lost when countries are resampled or dropped, most markedly for cluster 3, which contains Pakistan alone.

**Figure S5: Age and sex structure of total-stroke burden across the ten SCO member states and globally, 2023**

Age-specific incidence, mortality, and DALY rates per 100 000 (rows) by age group—under 20 years, then 5-year groups to 90–94 years, and 95 years and older (x-axis)—for the global population and each member state (y-axis), shown separately for males and females (columns). Cell shading is the rate on a log10 scale, common to all panels so that measures and sexes are directly comparable; the global row, in bold, is shown for reference.

**Abbreviations**

DALY=disability-adjusted life-year. SCO=Shanghai Cooperation Organisation.

**Figure S6: Age and sex structure of stroke-subtype burden, 2023**

As figure S5, with columns giving each combination of stroke subtype and sex. The age groups are those of figure S5; here the x-axis is ticked at the boundary ages between adjacent groups rather than labelled group by group.

**Abbreviations**

DALY=disability-adjusted life-year. ICH=intracerebral haemorrhage. IS=ischaemic stroke. SAH=subarachnoid haemorrhage. SCO=Shanghai Cooperation Organisation.

**Figure S7: Frontier analysis of total stroke burden against SDI, 2001–2023**

(A) Trajectories of the ten SCO member states in the SDI–ASDR plane from 2001 to 2023. Each line is one member state coloured by year, open circles marking 2001 and filled circles 2023; grey points are all other GBD locations in all years.

(B) The 2023 cross-section, in which each point is one GBD location coloured by whether its relative distance from the frontier fell (closer to frontier) or rose (farther from frontier) between 2001 and 2023; member states are ringed and labelled, the two closest to the frontier and the two farthest from it being ringed more heavily. Country codes are as in figure 3B. In both panels the solid line is the burden frontier, the lowest ASDR attainable at each level of SDI as estimated by bootstrapped data envelopment analysis, and the dashed line is a generalised additive model fitted to the observed rates; both are estimated from all GBD locations and not from the ten member states alone. The y axis is reversed so that lower burden lies towards the top and proximity to the frontier can be read directly. The nonlinearity p value in each panel is from a likelihood-ratio test of the fitted smooth against a linear fit. R is the Pearson correlation between SDI and ASDR—taken across all location-years in (A) and across locations in 2023 in (B)—and describes the linear component only; no p value is given for it in either panel, for the reasons set out in table S22, where both quantities are given for all four outcomes.

**Abbreviations**

Member states are identified on the map by three-letter ISO country code—BLR=Belarus, CHN=China, IND=India, IRN=Iran, KAZ=Kazakhstan, KGZ=Kyrgyzstan, PAK=Pakistan, RUS=Russia, TJK=Tajikistan, UZB=Uzbekistan—and the same codes are used in supplementary figures. ASDR=age-standardised DALY rate. DALY=disability-adjusted life-year. GBD=Global Burden of Diseases, Injuries, and Risk Factors Study. SCO=Shanghai Cooperation Organisation. SDI=Socio-demographic Index.

**Figure S8: Frontier analysis of stroke-subtype burden against SDI, 2001–2023**

As figure S7, for (A) ischaemic stroke, (B) intracerebral haemorrhage, and (C) subarachnoid haemorrhage. The upper row shows the trajectories of the ten SCO member states from 2001 to 2023 and the lower row the 2023 cross-section; the burden frontier and the fitted generalised additive model are estimated separately for each subtype from all GBD locations, not from the ten member states alone. Country codes are as in figure 3B. The ASDR scale differs between subtypes and between the two rows, so panels are comparable in shape but not in magnitude. For subarachnoid haemorrhage the fitted smooth is strongly nonlinear yet nearly symmetrical about its peak, so R is near zero in both rows (0·00 across all location-years; −0·13 in 2023); this indicates a burden peak at intermediate SDI rather than the absence of an SDI effect.

**Abbreviations**

Member states are identified on the map by three-letter ISO country code—BLR=Belarus, CHN=China, IND=India, IRN=Iran, KAZ=Kazakhstan, KGZ=Kyrgyzstan, PAK=Pakistan, RUS=Russia, TJK=Tajikistan, UZB=Uzbekistan—and the same codes are used in supplementary figures. ASDR=age-standardised DALY rate. DALY=disability-adjusted life-year. GBD=Global Burden of Diseases, Injuries, and Risk Factors Study. ICH=intracerebral haemorrhage. IS=ischaemic stroke. SAH=subarachnoid haemorrhage. SCO=Shanghai Cooperation Organisation. SDI=Socio-demographic Index.

**Figure S9: Metabolic attribution profiles by stroke subtype across the ten SCO member states, 2023**

As figure 3A, for (A) ischaemic stroke, (B) intracerebral haemorrhage, and (C) subarachnoid haemorrhage. Within each panel the upper heatmap gives the population attributable fraction and the lower heatmap the attributable ASDR per 100 000 for each metabolic risk factor (rows) in each member state (columns); the upper number in each cell is the rank of that risk factor within that state and measure. Columns are ordered by the dendrogram above each panel and the coloured bar marks membership of the two-cluster solution: cluster 1 comprises China, India, Pakistan, Tajikistan, and Uzbekistan in (A) and China, India, and Pakistan in (B) and (C), with the remaining states forming cluster 2 in each panel. The cluster and PAF colour keys shown in (A) apply to all three panels. The number of risk factors differs between subtypes because GBD 2023 estimates a different set of metabolic risk–outcome pairs for each: five for ischaemic stroke, four for intracerebral haemorrhage, and two for subarachnoid haemorrhage. Each panel's attributable-ASDR colour scale is normalised to that subtype's own maximum, so blue shading is not comparable between panels.

**Abbreviations**

ASDR=age-standardised DALY rate. BMI=body-mass index. C1, C2=metabolic-attribution clusters 1 and 2, C1 being the cluster containing China. DALY=disability-adjusted life-year. FG=fasting plasma glucose. GBD=Global Burden of Diseases, Injuries, and Risk Factors Study. ICH=intracerebral haemorrhage. IS=ischaemic stroke. KD=kidney dysfunction. LDL=low-density lipoprotein cholesterol. PAF=population attributable fraction. SAH=subarachnoid haemorrhage. SBP=systolic blood pressure. SCO=Shanghai Cooperation Organisation.

**Figure S10: Geographical distribution of the metabolic-attribution clusters by stroke subtype, 2023**

As figure 3B, for (A) ischaemic stroke, (B) intracerebral haemorrhage, and (C) subarachnoid haemorrhage. Member states are coloured by their cluster in the two-cluster solution for that subtype, with cluster 1 defined as the cluster containing China; non-member states are shown in grey. Each panel is annotated with the Mantel correlation between geographical distance, taken between capital cities, and distance in metabolic-attribution profile, and beneath it the same correlation with the estimation hierarchy held fixed, each with its p value on 99 999 permutations; the hierarchy distance, and what holding it fixed can and cannot show, are defined in figure 3B. The association was attenuated in all three subtypes: it persisted for ischaemic stroke and for intracerebral haemorrhage and fell to near zero for subarachnoid haemorrhage. Full test statistics are in table S26, including the join-count statistic, which has no standard partial form and is reported unadjusted throughout. The clusters for intracerebral and subarachnoid haemorrhage are identical to each other and to the total-stroke clusters shown in figure 3B, so panels (B) and (C) differ only in the annotated test statistics; ischaemic stroke differs in that Tajikistan and Uzbekistan fall in cluster 1. Cluster membership for every subtype is listed in table S25.

**Abbreviations**

Member states are identified on the map by three-letter ISO country code—BLR=Belarus, CHN=China, IND=India, IRN=Iran, KAZ=Kazakhstan, KGZ=Kyrgyzstan, PAK=Pakistan, RUS=Russia, TJK=Tajikistan, UZB=Uzbekistan—and the same codes are used in supplementary figures. C1, C2=metabolic-attribution clusters 1 and 2, C1 being the cluster containing China. GBD=Global Burden of Diseases, Injuries, and Risk Factors Study.

**Figure S11: Stability of the metabolic-attribution clustering for total stroke**

(A) Country co-assignment frequencies at k=2: the proportion of 1000 resamples—each obtained by adding gaussian noise with a standard deviation of 5% of each feature's standard deviation to the standardised risk-factor matrix and reclustering—in which a given pair of member states fell in the same cluster (1·00=always in the same cluster, 0·00=never). States are grouped by their cluster assignment from figure 3—cluster 1 (China, India, Pakistan) and cluster 2 (Belarus, Iran, Kazakhstan, Kyrgyzstan, Russia, Tajikistan, Uzbekistan)—and ordered alphabetically within each cluster; the black lines mark the cluster boundaries. Co-assignment at k=2 was perfect for every state (0 or 1 throughout); the three-country and seven-country clusters were reproduced exactly in every resample.

(B) Mean Jaccard similarity of each cluster to its closest counterpart in resampled data, at k=2, 3, and 4, under three resampling schemes implemented in fpc::clusterboot—non-parametric bootstrapping, jittering, and 80% subsetting—with 1000 replicates each; the jittering here is scaled to the local spacing of the data along each principal direction and is therefore not the fixed perturbation used in (A) (appendix). Dashed lines mark the conventional thresholds of 0·60, 0·75, and 0·85, at or above which a cluster is regarded as indicating a pattern, as stable, and as highly stable, respectively; higher values indicate a more reproducible cluster. Cluster numbers are internal to each value of k and correspond to those of figure 3 only in the k=2 panel, which is the solution selected by maximum mean silhouette width and reported throughout (table S25).

**Figure S12: Age-specific metabolic risk attribution for stroke and its subtypes, 2023**

Difference in the population attributable fraction of stroke DALYs between each age subgroup and the corresponding all-age estimate (ΔPAF), in percentage points, for every member state and risk factor. Row blocks are total stroke, ischaemic stroke, intracerebral haemorrhage, and subarachnoid haemorrhage; the last two include fewer risk factors because GBD estimates attribution only for those exposures. Each cell gives ΔPAF, with positive values indicating that the risk factor accounts for a larger share of the burden in that subgroup than at all ages. The reference is the age-standardised all-age estimate, so both age subgroups can differ from it in the same direction; the two columns are not a decomposition of the all-age value. The coloured bar at the top of each row block gives the attribution cluster for that outcome, which differs between outcomes: Tajikistan and Uzbekistan fall in C1 for ischaemic stroke but in C2 for the other three (table S25). The colour scale is truncated at the 95th centile of |ΔPAF| and cells beyond it are shown at the extreme colour; exact values are printed in every cell. These analyses were secondary and were not corrected for multiplicity.

**Abbreviations**

ASDR=age-standardised DALY rate. BMI=body-mass index. C1, C2=metabolic-attribution clusters 1 and 2. DALY=disability-adjusted life-year. FG=fasting plasma glucose. GBD=Global Burden of Diseases, Injuries, and Risk Factors Study. ICH=intracerebral haemorrhage. IS=ischaemic stroke. KD=kidney dysfunction. LDL=low-density lipoprotein cholesterol. PAF=population attributable fraction. SAH=subarachnoid haemorrhage. SBP=systolic blood pressure.

**Figure S13: Sex-specific metabolic risk attribution for stroke and its subtypes, 2023**

As figure S12, comparing the sex-specific PAF with the both-sex estimate. Both are age-standardised, so the comparison is not affected by differences in age structure between men and women; the colour scale is set independently of figure S12 and spans a narrower range.

**Abbreviations**

BMI=body-mass index. C1, C2=metabolic-attribution clusters 1 and 2. FG=fasting plasma glucose. KD=kidney dysfunction. LDL=low-density lipoprotein cholesterol. PAF=population attributable fraction. SBP=systolic blood pressure.

**Figure S14: Stroke-subtype composition by incidence and its alignment with the metabolic-attribution clusters, 2023**

As figure 4, using the proportions of IS, ICH, and SAH in ASIR rather than ASDR. Left, dendrogram of the ten member states clustered on the isometric-log-ratio-transformed incidence proportions (Ward's linkage, with k fixed at 2 to match the attribution clustering); the axis is the Ward merge distance. Centre, stacked bars of the three subtype proportions, labelled where the proportion is at least 3%. Right, colour strip giving each state's metabolic-attribution cluster (C1, C2; figure 3). Country-label colour denotes the incidence-based subtype-composition cluster (SC1, SC2); SC1 and C1 are the clusters containing China. The ASDR- and ASIR-based subtype-composition clusterings assign the same ten states to SC1 and SC2, shown as a tanglegram in figure S15 (table S27).

**Abbreviations**

ASDR=age-standardised DALY rate. ASIR=age-standardised incidence rate. C1, C2=metabolic-attribution clusters 1 and 2, C1 being the cluster containing China. DALY=disability-adjusted life-year. ICH=intracerebral haemorrhage. IS=ischaemic stroke. SAH=subarachnoid haemorrhage. SC1, SC2=stroke subtype-composition clusters 1 and 2, SC1 and C1 both being the clusters containing China.

**Figure S15: Correspondence between subtype-composition clusters and the metabolic-attribution clusters**

Tanglegrams comparing, for (A) ASDR and (B) ASIR, the dendrogram of figure 4 (or figure S14, respectively; left, "Subtype") with the metabolic-attribution dendrogram of figure 3A (right, "Metabolic PAF"), both restricted to the ten member states and both from Ward's-linkage clustering—on ilr-transformed subtype proportions for the left-hand dendrogram and on the five PAFs standardised within risk factor for the right-hand one. Country order within each dendrogram follows its own clustering, so the two sides need not align; connecting lines join each country to itself, and crossing lines mark countries whose relative position differs between the two clusterings. Left branch and label colours give the subtype-composition cluster (SC1, SC2) at k=2; right branch and label colours give the metabolic-attribution cluster (C1, C2; figure 3), also at k=2, shared by both panels. The ASDR- and ASIR-based subtype-composition clusterings assign the same ten states to SC1 and SC2 (table S27), so the two panels differ only in the ordering of the tips. No connecting line crosses the SC1/SC2 or the C1/C2 boundary in either panel: the crossings shown reflect the order of adjacent tips within a cluster, not membership. Membership discordance—Tajikistan and Uzbekistan falling in SC1 but in attribution-C2—is shown by the change in label colour between the two sides rather than by a crossing line.

**Abbreviations**

ASDR=age-standardised DALY rate. ASIR=age-standardised incidence rate. C1, C2=metabolic-attribution clusters 1 and 2, C1 being the cluster containing China. DALY=disability-adjusted life-year. ILR=isometric log-ratio. PAF=population attributable fraction. SC1, SC2=stroke subtype-composition clusters 1 and 2, SC1 and C1 both being the clusters containing China.

**Figure S16: Stability of the ASDR-based subtype-composition clustering**

(A) Country co-assignment frequencies at k=2: the proportion of 1000 resamples—each obtained by adding gaussian noise with a standard deviation of 5% of each feature's standard deviation to the ilr-transformed subtype-proportion matrix and reclustering—in which a given pair of member states fell in the same cluster (1·00=always in the same cluster, 0·00=never). States are grouped by their subtype-composition cluster assignment from figure 4—SC1 (China, India, Pakistan, Tajikistan, Uzbekistan) and SC2 (Belarus, Iran, Kazakhstan, Kyrgyzstan, Russia)—and ordered alphabetically within each cluster; the black lines mark the cluster boundaries. Co-assignment was near-complete for most states; Kazakhstan was the most ambiguous (0·13–0·17 with SC1, 0·87–0·89 with SC2).

(B) Mean Jaccard similarity of each cluster to its closest counterpart in resampled data, at k=2, 3, and 4, under three resampling schemes implemented in fpc::clusterboot—non-parametric bootstrapping, jittering, and 80% subsetting—with 1000 replicates each; the jittering here is scaled to the local spacing of the data along each principal direction and is therefore not the fixed perturbation used in (A) (appendix). Dashed lines mark the conventional thresholds of 0·60, 0·75, and 0·85, at or above which a cluster is regarded as indicating a pattern, as stable, and as highly stable, respectively; higher values indicate a more reproducible cluster. Cluster numbers are internal to each value of k and correspond to those of figure 4 only in the k=2 panel, which is the solution fixed to match the metabolic-attribution clustering and reported throughout (table S27).

**Abbreviations**

SC1, SC2=stroke subtype-composition clusters 1 and 2, SC1 being the cluster containing China.

**Figure S17: Participant flow through the CHARLS analysis**

Exclusions were applied sequentially to the 25 586 wave-1 participants: prevalent stroke or absent follow-up, then missing anthropometric data or age under 45 years, then absent, non-fasting, or incomplete blood-based measures, then physiologically implausible exposure values, leaving 8012 eligible participants. Counts at each step are given in the figure and the plausibility ranges in the appendix. The lower part of the figure shows the samples entering each analysis. Participants with incomplete covariate data were excluded when the models were fitted, so the number entering each model is smaller than the sample above it; these differences are unions of the per-variable missing counts, which overlap. In the primary analysis 116 participants were excluded from Models 1–2 (smoking 1, alcohol use 5, heart disease 29, depression 82) and 302 from Model 3, which additionally adjusts for antihypertensive, glucose-lowering, and lipid-lowering drug use (missing in 40, 76, and 165 participants, respectively). The cause-specific Cox sensitivity analysis uses the same samples as the primary Fine-Gray models. The two-wave sensitivity analysis takes wave 3 as the start of follow-up, so its exclusions concern wave-3 participation and measurement. Numbers in the model boxes are incident strokes; competing deaths are shown for the eligible cohort and the two-wave mean sample.

**Abbreviations**

BMI=body-mass index. CHARLS=China Health and Retirement Longitudinal Study. eGFR=estimated glomerular filtration rate. LDL=low-density lipoprotein cholesterol. SBP=systolic blood pressure.

**Figure S18: Cumulative incidence of stroke by clinical cut-off**

Cumulative incidence of first stroke with non-stroke death as the competing event, stratified by prespecified clinical cut-offs and ordered as in figure 5: (A) systolic blood pressure 140 mmHg, (B) body-mass index 28 kg/m² (the Chinese criterion for obesity), (C) fasting plasma glucose 126 mg/dL, (D) LDL cholesterol 160 mg/dL, and (E) eGFR 60 mL/min per 1·73 m². For eGFR the higher-risk stratum is the one below the cut-off and for the other four the one at or above it; the number of participants in each stratum is given in the key. p values are from Gray's test for equality of the cumulative incidence functions between strata and are not corrected for multiplicity. Because CHARLS ascertains outcomes at biennial waves and event times were imputed at wave midpoints, the curves rise in steps at the wave boundaries rather than continuously, the rise between years 5 and 6 corresponding to the final wave. The y axis is truncated at 0·15. These strata are descriptive and were not used in the modelling, which treated all five exposures as continuous (figure 5, tables S29–S32).

**Abbreviations**

CHARLS=China Health and Retirement Longitudinal Study. eGFR=estimated glomerular filtration rate. LDL=low-density lipoprotein cholesterol.

**Figure S19: Exposure–response curves for the five metabolic exposures and incident stroke**

Log subdistribution hazard ratio (line) with 95% CI (band) for (A) systolic blood pressure, (B) body-mass index, (C) fasting plasma glucose, (D) LDL cholesterol, and (E) eGFR, from four-knot restricted cubic splines fitted on the subdistribution hazard scale with non-stroke death as the competing event, each exposure modelled separately with the Model 1 covariate set (table S9). Curves are plotted against the reference value for each exposure (dotted vertical line; 115 mmHg, 22 kg/m², 90 mg/dL, 100 mg/dL, and 100 mL/min per 1·73 m² respectively), at which the curve passes through zero by construction; the dashed horizontal line marks the null. Tick marks on the lower axis show knot positions, and curves span the 1st to 99th percentiles of each exposure. Nonlinearity p values are from a two-degrees-of-freedom Wald test of the two nonlinear spline terms, corrected for false discovery rate across the five exposures; the test statistics, the three-degrees-of-freedom test of overall association, and fitted values at selected exposure concentrations are in tables S30–S31. The y-axis scale differs between panels.

**Abbreviations**

CI=confidence interval. eGFR=estimated glomerular filtration rate. FDR=false discovery rate. LDL=low-density lipoprotein cholesterol. SHR=subdistribution hazard ratio.

**Figure S20: Machine-learning corroboration of the exposure ranking in CHARLS**

(A) Mean absolute SHAP value for each predictor in the gradient-boosted tree model (XGBoost), fitted to the 7238 participants without a competing death event who had complete covariate data, of whom 565 had an incident stroke; models were trained on a random 70% of these participants. Variables are ranked by importance. Marital status and education were entered as indicator variables; levels are shown against their reference categories, which were married or cohabiting and less than lower secondary education, respectively. A bar of zero length denotes a variable never used for a split.

(B) Absolute standardised coefficient for each predictor in the penalised logistic (lasso) model, fitted to the same participants and variables as in (A). A bar of zero length denotes a variable shrunk to zero by the lasso.

(C) Receiver operating characteristic curves in the 30% test set for XGBoost and the lasso, with the area under each curve and the DeLong test for the difference between them. Five-fold cross-validated areas under the curve over the whole sample were 0·66 (SD 0·02) for XGBoost and 0·68 (SD 0·03) for the lasso.

(D) SHAP dependence plot for systolic blood pressure in the XGBoost training set. Grey points are individual participants, the blue line is a loess fit, and the horizontal axis is truncated at the 99·5th centile of the exposure while the fit uses all observations.

(E) As (D), for body-mass index.

(F) As (D), for fasting plasma glucose.

(G) As (D), for LDL cholesterol.

(H) As (D), for estimated glomerular filtration rate. These analyses were interpreted for aetiological ranking, not prediction.

**Abbreviations**

AUC=area under the curve. BMI=body-mass index. eGFR=estimated glomerular filtration rate. FG=fasting plasma glucose. LDL=low-density lipoprotein cholesterol. SBP=systolic blood pressure. SD=standard deviation. SHAP=SHapley Additive exPlanations.

**Table S1: Structural heterogeneity of thirteen intergovernmental organisations**

Organisations are ordered by the number of GBD super-regions spanned. Data are counts unless otherwise stated, and member states are counted as GBD locations rather than as formal memberships, so a member absent from the GBD location hierarchy is not counted. Membership is as at 2026 throughout: Timor-Leste is counted as an ASEAN member, Turkmenistan is excluded from the Commonwealth of Independent States as an associate member, and Saudi Arabia is excluded from BRICS as an invited state whose accession is unconfirmed, an exclusion that changes no value in this table other than the count of member states. Eligibility criteria and the basis for selecting these thirteen organisations are in appendix 1.1. The geographical coherence and health coordination capacity of the same thirteen organisations are in table S2.

GBD super-regions follow the IHME location hierarchy for GBD 2023. The count is an ordinal index on a seven-category partition and no arithmetic is performed on it. Genetic ancestry groups were assigned by mapping the predominant population of each member state to the 1000 Genomes Project superpopulations, supplemented by the Genome Aggregation Database Middle Eastern group, with Central Asian and Oceanian added for populations matched by no current reference panel; the mapping assigns one group per country and does not represent within-country ancestral diversity (appendix 1.3, table S3). Groups without a matched reference panel are included in the ancestry group count and shown separately in the adjacent column, so that count is not a count of groups for which genetic data exist. Reference panel availability and instrument availability are distinct again: of the five groups represented in the SCO, stroke genome-wide association data at the scale instrument selection requires exist for the European, east Asian, and south Asian groups only, so five SCO member states contribute no genetic evidence to this study. SDI is the Socio-demographic Index for 2023 (IHME); the range is the arithmetic difference between the highest and lowest member state values, is reported for context, and was not used in selection. Because Iran and Belarus acceded to the SCO in 2023 and 2024, burden trends reported elsewhere in this study from 2001 onwards describe the current membership retrospectively rather than the bloc as constituted at the time. The sensitivity of the ancestry group counts to the coding of the Central Asian group is in table S5.

**Abbreviations**

ASEAN=Association of Southeast Asian Nations. BRICS is a proper name and not an initialism of its current membership: the grouping was formed by Brazil, Russia, India, China, and South Africa, and comprises ten member states as at 2026. GBD=Global Burden of Diseases, Injuries, and Risk Factors Study. IHME=Institute for Health Metrics and Evaluation. SAARC=South Asian Association for Regional Cooperation. SCO=Shanghai Cooperation Organisation. SDI=Socio-demographic Index.

**Table S2: Geographical coherence and health coordination capacity of thirteen intergovernmental organisations**

Organisations are ordered by the number of GBD super-regions spanned, as in table S1, where the structural and developmental characteristics of the same thirteen organisations are given. Data are counts unless otherwise stated. Membership is as at 2026 throughout: Timor-Leste is counted as an ASEAN member, Turkmenistan is excluded from the Commonwealth of Independent States as an associate member, and Saudi Arabia is excluded from BRICS as an invited state whose accession is unconfirmed, an exclusion that changes no value in this table. Eligibility criteria and the basis for selecting these thirteen organisations are in appendix 1.1.

Connected components are the maximal sets of member states mutually reachable through land borders internal to the organisation; borders with non-member states create no edge, and maritime links, including bridges and causeways, are excluded (appendix 1.4). A count of one therefore means that every member state can be reached from every other by land without leaving the organisation. The size of the largest component is given as a count of member states, with the percentage of the organisation's member states that it contains in parentheses. Algebraic connectivity is the second-smallest eigenvalue of the Laplacian of the largest component. It is zero for a disconnected graph and rises as a component acquires more internal borders and more alternative paths between its members, so it separates a component strung together in a single chain, which one removal would break, from one that is densely interlinked. It falls as the number of member states in a component rises and is not comparable between components of very different size; it is therefore reported only where the largest component contains eight or more member states, and an ellipsis (···) marks a component below that threshold rather than a missing value. It is given because the count of components is binary in the case that matters and would not distinguish blocs that are contiguous to different degrees.

Standing health coordination mechanisms are the principal bodies through which member states coordinate health policy, with the year first convened or established where documented; the list is not exhaustive. Bloc-wide targets were recorded as adopted where a formally endorsed strategy sets shared objectives for non-communicable diseases or their risk factors. Both health coordination columns are descriptive and were not used to characterise organisations on the structural dimensions in table S1: a standing health body is a condition of eligibility and so does not vary here, and the adoption of bloc-wide targets bears on the timeliness of this analysis rather than on its validity.

**Abbreviations**

AIDS=acquired immunodeficiency syndrome. ASEAN=Association of Southeast Asian Nations. BRICS is a proper name and not an initialism of its current membership: the grouping was formed by Brazil, Russia, India, China, and South Africa, and comprises ten member states as at 2026. EU=European Union. GBD=Global Burden of Diseases, Injuries, and Risk Factors Study. HIV=human immunodeficiency virus. NCD=non-communicable disease. SAARC=South Asian Association for Regional Cooperation.

**Table S3: Assignment of 155 countries to genetic ancestry groups**

Genetic ancestry groups were assigned by mapping the predominant population of each country to the 1000 Genomes Project superpopulations, supplemented by the Genome Aggregation Database Middle Eastern group, with Central Asian and Oceanian added for populations that no current reference panel matches. One group is assigned per country. The mapping is coarse: it does not represent within-country ancestral diversity, and the groups are labels applied to a continuum of genetic variation rather than discrete categories (appendix 1.3). The groups themselves, their reference panels, and the number of countries assigned to each are in table S4.

All 155 countries appearing in at least one of the thirteen organisations, with the group assigned to each and the organisations to which it belongs. Membership is denoted by a filled circle (●). Country names and super-regions follow the IHME location hierarchy for GBD 2023. Super-region and ancestry group are independent classifications made for different purposes, so they need not agree: Azerbaijan and Türkiye are assigned to the Middle Eastern group while falling in the Central Europe, Eastern Europe, and Central Asia and the North Africa and Middle East super-regions respectively, and Maldives is assigned to the south Asian group while falling in the Southeast Asia, East Asia, and Oceania super-region. Turkmenistan appears in the Central Asian group of the Organisation of Islamic Cooperation but not of the Commonwealth of Independent States, from which it is excluded as an associate member (appendix 1.1); it is the one Central Asian country that is not an SCO member. Also supplied as a spreadsheet.

**Abbreviations**

AFR=African. AMR=admixed American. ASEAN=Association of Southeast Asian Nations. AU=African Union. BRICS is a proper name and not an initialism of its current membership. CAS=Central Asian. CIS=Commonwealth of Independent States. CW=Commonwealth of Nations. EAEU=Eurasian Economic Union. EAS=east Asian. EU=European Union. EUR=European. GBD=Global Burden of Diseases, Injuries, and Risk Factors Study. GCC=Gulf Cooperation Council. gnomAD=Genome Aggregation Database. IHME=Institute for Health Metrics and Evaluation. LAS=League of Arab States. MID=Middle Eastern. Nordic=Nordic Council. OCE=Oceanian. OIC=Organisation of Islamic Cooperation. SAARC=South Asian Association for Regional Cooperation. SAS=south Asian. SCO=Shanghai Cooperation Organisation.

**Table S4: Genetic ancestry groups, reference panels, and coverage**

Genetic ancestry groups were assigned by mapping the predominant population of each country to the 1000 Genomes Project superpopulations, supplemented by the Genome Aggregation Database Middle Eastern group, with Central Asian and Oceanian added for populations that no current reference panel matches. One group is assigned per country. The mapping is coarse: it does not represent within-country ancestral diversity, and the groups are labels applied to a continuum of genetic variation rather than discrete categories (appendix 1.3). The assignment of each of the 155 countries is in table S3.

Countries is the number of the 155 countries in table S3 assigned to each group, not a count of all countries worldwide. Reference panel is the source from which allele frequencies and linkage disequilibrium structure for that group would be taken; None available means that no panel of comparable scale currently exists, which is the reason the Central Asian and Oceanian groups were added rather than a property of the populations themselves. An em dash (—) denotes a group with no SCO member state. Availability of a reference panel and availability of instruments are distinct. Of the five groups represented in the SCO, stroke genome-wide association data at the scale instrument selection requires exist for EUR, EAS, and SAS only; MID and CAS have none, so five SCO member states—Iran, Kazakhstan, Kyrgyzstan, Tajikistan, and Uzbekistan—contribute no genetic evidence to this study (appendix 6.1).

**Abbreviations**

AFR=African. AMR=admixed American. CAS=Central Asian. EAS=east Asian. EUR=European. gnomAD=Genome Aggregation Database. MID=Middle Eastern. OCE=Oceanian. SAS=south Asian. SCO=Shanghai Cooperation Organisation.

**Table S5: Genetic ancestry group counts under alternative codings of the Central Asian group**

Organisations are ordered by the number of GBD super-regions spanned, as in table S1. Membership is as at 2026 throughout: Timor-Leste is counted as an ASEAN member, Turkmenistan is excluded from the Commonwealth of Independent States as an associate member, and Saudi Arabia is excluded from BRICS as an invited state whose accession is unconfirmed, an exclusion that changes no value in this table. Eligibility criteria and the basis for selecting these thirteen organisations are in appendix 1.1.

The Central Asian populations of Kazakhstan, Kyrgyzstan, Tajikistan, and Uzbekistan are matched by no current reference panel, so treating them as a distinct ancestry group is a coding decision and not an empirical finding. Each column gives the number of ancestry groups an organisation spans under one treatment of that decision: the baseline column counts Central Asian as its own unrepresented group, as in table S1, and the remaining three columns instead fold it into the European, east Asian, or south Asian group. The table is given to show that the comparison in table S1 does not rest on the coding. The SCO spans five groups at baseline and four under every alternative, and in all four codings only three organisations span more, so its position relative to the other twelve is unchanged.

**Abbreviations**

ASEAN=Association of Southeast Asian Nations. BRICS is a proper name and not an initialism of its current membership: the grouping was formed by Brazil, Russia, India, China, and South Africa, and comprises ten member states as at 2026. GBD=Global Burden of Diseases, Injuries, and Risk Factors Study. SAARC=South Asian Association for Regional Cooperation. SCO=Shanghai Cooperation Organisation.

**Table S6: GBD 2023 measures and ICD-10 mapping**

Total stroke comprises the three subtypes listed. Country names were harmonised as Russian Federation→Russia and Iran (Islamic Republic of)→Iran.

**Abbreviations**

DALY=disability-adjusted life-year. GBD=Global Burden of Diseases, Injuries, and Risk Factors Study. ICD-10=International Classification of Diseases, 10th revision. LDL=low-density lipoprotein cholesterol. PAF=population attributable fraction. SCO=Shanghai Cooperation Organisation.

**Table S7: MR exposure GWAS, by ancestry**

N is the GWAS sample size for the trait. Dataset IDs are OpenGWAS or GWAS Catalog accessions; — denotes summary statistics obtained directly from the study without a public accession. European traits other than FG are the population (not within-sibship) estimates released by the within-family consortium; the within-sibship subsamples yielded too few genome-wide significant variants to instrument the exposures. These estimates are therefore not adjusted for within-family confounding. Pan-UKBB (CSA) is the central and south Asian panel.

**Abbreviations**

BBJ=BioBank Japan. BMI=body-mass index. CSA=central and south Asian. eGFR=estimated glomerular filtration rate. FG=fasting plasma glucose. GWAS=genome-wide association study. LDL=low-density lipoprotein cholesterol. MAGIC=Meta-Analyses of Glucose and Insulin-related traits Consortium. MR=mendelian randomisation. Pan-UKBB=Pan-UK Biobank. SBP=systolic blood pressure. TPMI=Taiwan Precision Medicine Initiative.

**Table S8: MR outcome GWAS, by ancestry**

The three total-stroke phenotypes count different things—a broad composite in European ancestry, unclassified stroke only (ICD-10 I64) in east Asian ancestry, and a composite in which 3462 of 3640 cases are ischaemic in south Asian ancestry—and were used within ancestry only. No south Asian GWAS was available for ICH or SAH. GIGASTROKE includes some Pan-UK Biobank participants, so modest overlap with the south Asian exposure samples is possible.

**Abbreviations**

CKB=China Kadoorie Biobank. GIGASTROKE is the name of the stroke genome-wide association consortium and is not an initialism. GWAS=genome-wide association study. ICD-10=International Classification of Diseases, 10th revision. ICH=intracerebral haemorrhage. MR=mendelian randomisation. SAH=subarachnoid haemorrhage. UKB=UK Biobank.

**Table S9: CHARLS covariates, coding, and model specification**

Coding follows the Harmonized CHARLS dataset. Depression is a CES-D-10 score above 10. Confounders with more than 20% missingness were excluded a priori.

**Abbreviations**

CES-D-10=10-item Center for Epidemiologic Studies Depression scale. CHARLS=China Health and Retirement Longitudinal Study.

**Table S10: Partitions of the ten member states at two to five clusters, total stroke**

Partitions are obtained by cutting the same dendrogram at each level and are listed in the order returned; clusters are separated by a vertical rule. Two clusters is the solution selected in the primary analysis, and cluster C1, so designated elsewhere in this report, is the cluster containing China. The clustering is of attribution profiles, each the vector of population attributable fractions of the five metabolic risk factors for one member state, standardised across the ten states (appendix 2.1).

ARI is the adjusted Rand index against the partition of the ten member states into the four GBD super-regions they span; the two-cluster value is the same statistic as the total-stroke entry in table S11, and no p values are repeated here because the comparison across cutting levels is descriptive. Agreement is not monotone in the number of clusters, which is expected: a coarse partition leaves undivided most of the pairs that the super-regions separate, so the two-cluster value is not evidence that the hierarchy is recovered (appendix 2.3).

**Abbreviations**

ARI=adjusted Rand index. GBD=Global Burden of Diseases, Injuries, and Risk Factors Study.

**Table S11: Alignment of the risk-structure clustering with the GBD estimation hierarchy, and the geographic gradient within a single super-region**

Three distances between pairs of member states are used in this section. An attribution profile is the vector of population attributable fractions of the five metabolic risk factors for one member state, each standardised across the ten states, and the distance between two profiles is Euclidean; it measures how differently two states' stroke burden is apportioned among the five factors. Geographic distance is the great-circle distance between capitals. Hierarchy distance is 0 where two member states belong to the same GBD region, 1 where they belong to the same super-region but different regions, and 2 otherwise; it stands for the opportunity GBD has to borrow strength between them when input data are sparse, so an association with it is what an artefact of the estimation procedure would look like (appendix 2.1). Permutation tests use 99 999 permutations and Mantel statistics are one sided. A column heading of the form X ∼ Y denotes the association between distance X and distance Y, and any distance named after adjusted for is the one held constant. These are sensitivity analyses of a prespecified result: the p values are nominal and are not corrected for the roughly 75 tests in this section (appendix 2.8). The corresponding comparison made through distances alone is in table S13.

The first four columns compare the risk-structure clustering with the hierarchy directly, rather than through distances. ARI is the adjusted Rand index between the cluster assignment for that outcome and the partition of the ten member states into GBD regions, or into super-regions; it is 1 for exact agreement and 0 for the agreement expected by chance, and p values are from 99 999 permutations of the cluster labels. Values are identical for total stroke, intracerebral haemorrhage, and subarachnoid haemorrhage because the cluster assignment is identical for those three outcomes.

The last four columns are restricted to the six member states that lie within one super-region, Central Europe, Eastern Europe, and Central Asia: Belarus, Kazakhstan, Kyrgyzstan, Russia, Tajikistan, and Uzbekistan. Super-region-level borrowing treats these six identically, so a geographic gradient among them cannot be produced by borrowing at that level. The final two columns hold region distance constant, which tests the finer level; that comparison is not identified, because within these six states the boundary between the Eastern Europe and Central Asia regions almost coincides with the geographic division and fifteen pairs cannot separate them. It is shown for completeness and no weight is placed on it (appendix 2.5).

**Abbreviations**

ARI=adjusted Rand index. GBD=Global Burden of Diseases, Injuries, and Risk Factors Study.

**Table S12: Robustness of the spatial concordance to omission of member states**

Each row repeats the primary Mantel test of attribution-profile distance against geographic distance on the member states that remain, with the standardisation, the clustering, and both distance matrices recomputed within the reduced set rather than taken as a submatrix of the full analysis. An attribution profile is the vector of population attributable fractions of the five metabolic risk factors for one member state, standardised within each factor across the states retained; geographic distance is the great-circle distance between capitals (appendix 2.1). p values are one sided, from 99 999 permutations, and are nominal (appendix 2.8).

*The first row is the primary analysis, quoted from table S26 so that each statistic in this report has a single source. It is retained because the leave-one-out rows cannot be read without a baseline.

The final row omits the only two member states whose cause-of-death data are rated two stars or fewer in the GBD data-quality assessment: India at two stars and Pakistan at one, all other member states at three or more. These two are also two of the three members of cluster C1, so the partition necessarily rearranges once they are removed and the cluster-stability statistics for this row are not interpretable; the row bears on the continuous geographic gradient only, and with eight member states rests on 28 country pairs.

Because the clustering is re-derived within each reduced set, the selected number of clusters changed from two to three in three of the ten leave-one-out analyses for total stroke, on omitting India, Kyrgyzstan, or Tajikistan, and those partitions differ from the primary one. This does not affect the statistics reported here: the Mantel test is computed from the distance matrices and does not use the cluster assignment.

**Abbreviations**

GBD=Global Burden of Diseases, Injuries, and Risk Factors Study.

**Table S13: Spatial concordance of metabolic attribution profiles adjusted for the GBD estimation hierarchy**

Three distances between pairs of member states are used in this section. An attribution profile is the vector of population attributable fractions of the five metabolic risk factors for one member state, each standardised across the ten states, and the distance between two profiles is Euclidean; it measures how differently two states' stroke burden is apportioned among the five factors. Geographic distance is the great-circle distance between capitals. Hierarchy distance is 0 where two member states belong to the same GBD region, 1 where they belong to the same super-region but different regions, and 2 otherwise; it stands for the opportunity GBD has to borrow strength between them when input data are sparse, so an association with it is what an artefact of the estimation procedure would look like (appendix 2.1). Statistics are Mantel and partial Mantel correlations from 99 999 permutations, one sided; a partial statistic is the association between two distances with the third held constant. A column heading of the form X ∼ Y denotes the association between distance X and distance Y, and any distance named after adjusted for is the one held constant. These are sensitivity analyses of a prespecified result: the p values are nominal and are not corrected for the roughly 75 tests in this section (appendix 2.8).

The two partial columns ask which of the two candidate explanations survives when the other is removed. The unadjusted association between geography and attribution, against which the first partial column should be read, is r=0·498 (p=0·0063) for total stroke, 0·555 (p=0·0005) for ischaemic stroke, 0·606 (p=0·0009) for intracerebral haemorrhage, and 0·389 (p=0·03) for subarachnoid haemorrhage. These are quoted from table S26, so that each statistic in this report has a single source: the correlations are identical to three decimal places in the run that produced the partial statistics shown here, and two of the four p values agree exactly, but permutation p values remain Monte Carlo estimates and may differ in their final digit between runs. Geographic and hierarchy distance are themselves correlated, and that association does not depend on the outcome: r=0·490, p=0·01 throughout. The adjusted geographic associations for ischaemic stroke and intracerebral haemorrhage are nominally significant and would not survive correction across the four outcomes; they are described as persisting rather than as significant (appendix 2.4, 2.8).

Two further comparisons with the hierarchy, made through cluster agreement rather than through distances, are in tables S10–S11; the robustness of the unadjusted geographic association to omission of member states is in table S12.

**Abbreviations**

GBD=Global Burden of Diseases, Injuries, and Risk Factors Study.

**Table S14: Instrument sources and the precision of the ancestry-specific mendelian randomisation analyses**

Each row is one combination of ancestry, exposure and outcome. Source and N are the exposure genome-wide association study and its sample size; the number of instruments retained differs between outcomes within an exposure because not every variant is present in every outcome dataset.

Largest effect not excluded is the limit of the 95% interval further from the null, and Side states which side that is. A 95% interval is the set of null values the same test would not reject, so its far limit is the largest effect the data leave open; because it is read after a null result, and conditioning on non-significance shrinks an estimate towards the null, a limit quoted this way understates rather than overstates the range still open.

Smallest detectable effect is the smallest odds ratio the analysis could have detected with 80% power at a two-sided alpha of 0·05, obtained as exp(2·802 × SE) on the log-odds scale, where 2·802 is the sum of the critical value and the 80th centile of the standard normal distribution. The calculation treats the standard error as fixed with respect to the true effect, and 0·80 is a convention rather than a property of the data.

Ratio and |Z|/2·802 make the same comparison, of the effect observed against the smallest the analysis could detect, on two different scales. Ratio divides the observed effect by the smallest detectable effect, the observed effect first expressed in its own direction so that both are at least 1; a value below 1 means the effect seen was smaller than the analysis could reliably have detected. Because it is a ratio of two odds ratios rather than of their logarithms, it is not free of the exposure scale and is a within-row indicator only. |Z|/2·802, the absolute z statistic divided by the same constant, is the corresponding quantity on the log scale, is therefore dimensionless, and equals 1 at the 80% detectability boundary; it is the only column that may be read down. It is a monotone function of the p value and is reported as an index rather than as evidence.

Verdict records the factor as detected where the FDR-corrected p value was below 0·05, and otherwise as null; a null is marked uninformative where Ratio is below 1, that is, where the analysis could not have detected an effect of the size observed. Detection was judged on the corrected p value, which is stricter than the uncorrected alpha used for the detectable effect, so that column is a lower bound on what detection required. Corrected p values below 0·0001 are shown as <0·0001; their relative ordering is preserved in the |Z|/2·802 column.

The estimate, its interval, the unexcluded bound and the detectable effect are all expressed on the scale in which the contributing study reports its exposure. No contributing study states its units or the standard deviation of the exposure in its own sample, so that scale is unknown and differs between studies: none of these columns, nor Ratio, may be compared between rows. Estimates are not placed on a common scale and no comparison of magnitude between ancestries is made anywhere in this study (appendix 3.4). Total stroke is reported here because precision is internal to a single analysis, but is excluded from the genetic-stream ordering and from the subtype contrasts because the available total-stroke phenotypes count different things across ancestries.

**Abbreviations**

BMI=body-mass index. CI=confidence interval. EAS=east Asian. eGFR=estimated glomerular filtration rate. EUR=European. FDR=false discovery rate. FG=fasting plasma glucose. GWAS=genome-wide association study. ICH=intracerebral haemorrhage. IS=ischaemic stroke. KD=kidney dysfunction. LDL=low-density lipoprotein cholesterol. MAGIC=Meta-Analyses of Glucose and Insulin-related traits Consortium. OR=odds ratio. SAH=subarachnoid haemorrhage. SBP=systolic blood pressure. SE=standard error. TPMI=Taiwan Precision Medicine Initiative.

**Table S15: Within-state rank frequencies and the inputs to the population-stream ordering**

For each risk factor the table gives the median and range of the attributable fraction across the ten member states, and the number of states in which the factor took each within-state rank. The ordering criterion is the median; the rank frequencies are given so that the ordering can be checked against the dispersion behind it, and so that a factor whose median position rests on a narrow margin can be identified. The final column counts the states in which a factor is placed above the factor that follows it in the pooled sequence, which runs SBP, LDL, kidney dysfunction, fasting glucose, BMI; it is given because the rank frequencies are marginal and do not by themselves determine how often a given pair is ordered one way. The five exceptions are India for LDL against kidney dysfunction, China and Tajikistan for kidney dysfunction against fasting glucose, and Belarus and Iran for fasting glucose against BMI; BMI has no entry, marked by an em dash (—), because no factor follows it. The same comparisons can be read directly from the within-state ranks printed in figure 3A. Values are derived from table 1.

**Abbreviations**

BMI=body-mass index. PAF=population attributable fraction. SBP=systolic blood pressure.

**Table S16: Mechanisms admitted as explanations for structurally explicable divergence**

The original rules were fixed before the streams were compared and are stated in terms of what each stream licenses rather than in terms of any particular exposure. After internal review but before submission, M2 was amended as described in the main text and appendix 3.2; the LDL classification is therefore interpreted under the amended qualitative criterion. The remaining rules retain their original scope and are intended for testing in another bloc or disease. Their application to each of the five factors is derived in table S17, and the explanations considered and not admitted are in table S18.

Mechanisms are labelled M1 to M4 and are referred to by those labels throughout this report. Predicted pattern is the pattern that must be present in the comparison, not one the mechanism merely allows. Precondition states what must additionally hold in the data, except for M4, where it states a restriction on scope instead: that mechanism may qualify a position in the genetic stream but may not be used to set aside an observational result.

**Table S17: Mechanical derivation of the triangulation classification for each risk factor**

The three stream columns each give the factor's position within that stream, followed by the quantity on which the position rests: for the genetic stream, the number of the six exposure–outcome comparisons in which the factor was detected, and whether the association persisted under multivariable adjustment. The six are the two instrumented ancestries, European and east Asian, crossed with the three stroke subtypes; south Asian analyses are exploratory and do not enter the count, and the composite total-stroke phenotypes are excluded because the east Asian one is not comparable with the others (appendix 6.6). For the population stream, the median attributable fraction across the ten member states and the number of states placing the factor at that rank; for the individual stream, the subdistribution hazard ratio with its 95% CI. Positions are internal to each stream and carry no quantitative equivalence between them: a factor placed second in two streams is not thereby estimated at the same magnitude twice. Ordering criteria for each stream are in appendix 3.1.

Divergence records whether the streams disagree in the specific senses the rules recognise, and by which of the two criteria. A divergence is recorded either where one stream flags a factor as not detected while another places it (the not-detected criterion), or where two streams both place the factor but at positions differing by two or more (the positional criterion, the entry giving the size of the gap). The three entries in this column read as follows. No flag; positions identical means neither criterion is met: no stream flagged the factor, and the three streams placed it at the same position, so there is nothing for the mechanisms to explain. Not-detected criterion means a factor was flagged as not detected in at least one stream while another placed it; because the population stream apportions a fraction to every factor and has no not-detected flag, this criterion can be triggered only by the genetic or the individual stream. Positional, 3 places means both streams placed the factor and their positions differ by three, meeting the threshold of two. A gap of one is below the threshold and is not recorded as a divergence, so a factor may sit at adjacent positions in two streams and still be classified without any mechanism being invoked for that pair.

Mechanism test records, for each factor, which mechanisms in table S16 were tested, which were admitted, and on what ground the remainder were declined. A mechanism is declined either because its precondition fails in the data or because the pattern it predicts is not the pattern observed; both grounds appear. Subtype contrasts within an ancestry require no rescaling of the exposure and are therefore comparable as reported.

Control checks. The rules decline as well as admit, and at both gates. M1's precondition fails for SBP, whose window lies within the cohort's range but carries no gradient inside it, so the mechanism that admits BMI is unavailable to the factor that converges. M2's direction requirement fails for BMI, whose genetic effect is confined to a single minority subtype but which the cohort detects, so the mechanism that admits LDL is unavailable to BMI. Fasting glucose shows a gradient inside its window as BMI does, yet M1 is not applicable to it because its population and individual positions are adjacent and no divergence between those two streams is recorded. Two of five factors resist classification altogether, and the two explanations most readily available for them, confounding and prevalence-driven attribution, are excluded by rule rather than by argument (table S18). Two further checks were run on the rules themselves: the second criterion of the genetic ordering separates at least two factors, so it is not inert; and of the fifteen subtype contrasts available in European ancestry, one survives correction, the LDL contrast on which mechanism M2 relies.

**Abbreviations**

BMI=body-mass index. C=convergence. CI=confidence interval. DALY=disability-adjusted life-year. FDR=false discovery rate. FG=fasting plasma glucose. GBD=Global Burden of Diseases, Injuries, and Risk Factors Study. ICH=intracerebral haemorrhage. IS=ischaemic stroke. KD=kidney dysfunction. LDL=low-density lipoprotein cholesterol. MVMR=multivariable mendelian randomisation. SAH=subarachnoid haemorrhage. SBP=systolic blood pressure. SED=structurally explicable divergence. TMREL=theoretical minimum risk exposure level. UD=unexplained divergence. eGFR=estimated glomerular filtration rate.

**Table S18: Explanations not admitted as structurally explicable divergence**

A divergence was recorded where two streams placed a factor at positions differing by two or more, or where one stream flagged the factor as not detected while another placed it (main text, Methods). A recorded divergence is classified as structurally explicable only where a mechanism listed in table S16 predicts both its presence and its direction and that mechanism's precondition holds; where none does, the factor is classified as unexplained and no post-hoc explanation is offered. This table lists the explanations that were considered and not admitted.

Explanations are labelled X1 to X3. Each is compatible with one or more of the patterns observed in this study. None is excluded for being implausible; each is excluded because it is a substantive claim about a particular exposure rather than a structural property of what the streams license, and admitting any of them would allow a divergence to be accounted for after the fact by whichever explanation happened to fit.

**Table S19: National age-standardised stroke burden, 2001 and 2023**

Rates are per 100 000 population with 95% uncertainty intervals; "Both" is both sexes combined. Global estimates are shown for reference and were not included in the clustering analyses.

**Abbreviations**

ASDR=age-standardised DALY rate. ASIR=age-standardised incidence rate. ASMR=age-standardised mortality rate.

**Table S20: Burden-trajectory cluster membership and stability**

Silhouette widths in the row headings are the mean values at k=2/3/4; the selected k is the one maximising them. Mean bootstrap Jaccard values in the same headings are the unweighted means across clusters at k=2/3/4; in every outcome the selected k also maximises them. Jaccard similarities are means over 1000 resamples under non-parametric bootstrapping, feature jittering (appendix), and 80% subsetting; values at or above 0·60, 0·75, and 0·85 indicate a cluster regarded as indicating a pattern, as stable, and as highly stable, respectively. Single-country clusters are inherently less reproducible.

**Table S21: Tests of geographical structure of the clusterings**

Mantel r is the Pearson correlation between great-circle distances separating national capitals and standardised feature distances (99 999 permutations). Join-count is the number of bordering state pairs assigned to the same cluster, tested one-sided against 99 999 permutations of the cluster labels; with few shared land borders among the ten states this statistic has low power.

**Table S22: Generalised additive model of the SDI–ASDR association**

Models were fitted across all GBD locations, not the ten member states alone. edf is the effective degrees of freedom of the SDI smooth, deviance explained is a percentage, and the nonlinearity p value is from a likelihood-ratio test of the smooth against a linear fit. R is the Pearson correlation between SDI and ASDR, taken across all location-years in the all-years columns and across locations in the 2023 columns, and describes the linear component of the association only. No p value is given for R: in the all-years columns the observations are repeated measures on the same locations, so a correlation test would be anticonservative, and in neither column does a test of linear correlation evaluate the fitted smooth, which the nonlinearity p value tests directly. *For SAH the smooth is strongly nonlinear but nearly symmetrical about its peak, so R is near zero across all location-years and small in 2023, where a linear-only model fitted for reference gives p=0·06. That p value is not tabulated for any outcome because it tests the linear component rather than the fitted smooth. R should not be read as evidence against an SDI effect.

**Abbreviations**

ASDR=age-standardised DALY rate. edf=effective degrees of freedom. GBD=Global Burden of Diseases, Injuries, and Risk Factors Study. SAH=subarachnoid haemorrhage. SDI=Socio-demographic Index.

**Table S23: Relative inefficiency (relative distance to frontier) in 2023, by SCO member state**

Relative inefficiency is the gap between a state's ASDR and the bootstrap data-envelopment frontier at its SDI, expressed as a proportion of the frontier value; 0 denotes a state on the frontier and larger values a greater excess burden. Trend gives the direction of movement relative to the frontier between 2001 and 2023.

**Abbreviations**

ASDR=age-standardised DALY rate. SCO=Shanghai Cooperation Organisation. SDI=Socio-demographic Index.

**Table S24: Metabolic-attribution cluster selection: silhouette widths, selected k, and composite-metric agreement**

Silhouette widths are mean values over k=2–5 and the selected k is the maximum. The composite metric is PAF × log(1 + attributable DALY rate). The ARI compares the PAF-based and composite-based partitions, each at its own selected k; 1 indicates identical partitions.

**Abbreviations**

ARI=adjusted Rand index. DALY=disability-adjusted life-year. PAF=population attributable fraction.

**Table S25: Metabolic-attribution cluster membership and stability (k=2), by outcome**

C1 is the cluster containing China. Jaccard similarities as defined in table S20.

**Abbreviations**

C1, C2=metabolic-attribution clusters 1 and 2.

**Table S26: Tests of geographical structure of the metabolic-attribution clusterings**

Statistics as defined in table S21.

**Table S27: Subtype-composition cluster membership and stability (k=2), by rate basis (DALY-based and incidence-based)**

Clustering was on isometric-log-ratio-transformed subtype proportions, with k fixed at 2 to match the attribution clustering; SC1 is the cluster containing China. Jaccard similarities as defined in table S20.

**Abbreviations**

ASDR=age-standardised DALY rate. ASIR=age-standardised incidence rate. DALY=disability-adjusted life-year. SC1, SC2=subtype-composition clusters 1 and 2.

**Table S28: Full pairwise comparisons of baseline characteristics, with p values and SMDs**

Comparisons are no event vs stroke, no event vs death, and stroke vs death. p values are from pairwise t tests or chi-square/Fisher exact tests and are not corrected for multiplicity. SMD asterisks as in table 2.

**Abbreviations**

BMI=body-mass index. eGFR=estimated glomerular filtration rate. FG=fasting plasma glucose. LDL=low-density lipoprotein cholesterol. SBP=systolic blood pressure. SMD=standardised mean difference.

**Table S29: Associations of metabolic risk factors with incident stroke: Fine-Gray SHRs (Models 1–3)**

SHRs are per 1 SD increment, with non-stroke death as the competing event; model specifications are in table S9. Sample sizes are given with each model. All models were fitted on complete cases, Model 3 being smaller because it additionally requires medication data; 8012 participants met the eligibility criteria. Marital status and education were entered as indicator variables. The five exposures were prespecified and mutually adjusted, so no multiplicity correction was applied.

**Abbreviations**

BMI=body-mass index. CI=confidence interval. eGFR=estimated glomerular filtration rate. FG=fasting plasma glucose. LDL=low-density lipoprotein cholesterol. SBP=systolic blood pressure. SD=standard deviation. SHR=subdistribution hazard ratio.

**Table S30: Restricted cubic spline nonlinearity tests for the five metabolic exposures**

Four-knot restricted cubic splines with knots at the 5th, 35th, 65th, and 95th percentiles of the analysis sample, fitted on the subdistribution hazard scale in 7896 participants (565 incident strokes) with the Model 1 covariate set. Overall p is a 3 df joint test of all spline coefficients and nonlinearity p a 2 df joint test of the nonlinear terms, with FDR correction across the five exposures. Reference values lie within the GBD TMREL window where one is defined on the same continuous scale (SBP, BMI, FG); no such alignment was possible for LDL or eGFR.

**Abbreviations**

BMI=body-mass index. eGFR=estimated glomerular filtration rate. FDR=false discovery rate. FG=fasting plasma glucose. GBD=Global Burden of Diseases, Injuries, and Risk Factors Study. LDL=low-density lipoprotein cholesterol. SBP=systolic blood pressure. TMREL=theoretical minimum risk exposure level.

**Table S31: Fitted SHRs at selected exposure values**

Each SHR contrasts the stated value against the reference for that exposure, from the spline models of table S30 (7896 participants; 565 incident strokes). p values are not corrected for multiple values within an exposure and should be read against the overall test in table S30, which was not significant for LDL or eGFR. BMI 18 and 32 kg/m², eGFR 45 and 120 mL/min per 1·73 m², and FG 160 mg/dL lie outside the 10th–90th percentile range of the cohort.

**Abbreviations**

BMI=body-mass index. CI=confidence interval. eGFR=estimated glomerular filtration rate. FG=fasting plasma glucose. LDL=low-density lipoprotein cholesterol. SBP=systolic blood pressure. SHR=subdistribution hazard ratio.

**Table S32: Test of the proportional subdistribution hazards assumption**

Exposure-by-log-time interaction added to the Fine-Gray model of Model 2 specification (7896 participants; 565 incident strokes). The SHR column is the fitted year-1 value and the log-time coefficient its rate of change, negative values indicating attenuation over follow-up. Event times were imputed at wave midpoints, so the year-1 estimates extrapolate the fitted trend beyond the observed data.

**Abbreviations**

BMI=body-mass index. eGFR=estimated glomerular filtration rate. FG=fasting plasma glucose. LDL=low-density lipoprotein cholesterol. SBP=systolic blood pressure. SD=standard deviation. SHR=subdistribution hazard ratio.

**Table S33: Subgroup-specific SHRs and tests of interaction**

Level 1 and Level 2 are the two strata named in each panel heading. SHRs are per 1 SD from Model 2 refitted within each stratum, standardised on the whole-sample SD; pinteraction is from exposure-by-subgroup product terms fitted in the full sample. Analyses were restricted to the 7896 participants (565 events) with complete covariate and stratifying-variable data.

**Abbreviations**

BMI=body-mass index. CI=confidence interval. eGFR=estimated glomerular filtration rate. FG=fasting plasma glucose. LDL=low-density lipoprotein cholesterol. SBP=systolic blood pressure. SD=standard deviation. SHR=subdistribution hazard ratio.

**Table S34: Cause-specific Cox hazard ratios for the five exposures**

Hazard ratios per 1 SD from cause-specific Cox models with competing deaths censored; Models 1–3 as in table S29, and fitted on the same complete-case samples, given with each model.

**Abbreviations**

BMI=body-mass index. eGFR=estimated glomerular filtration rate. FG=fasting plasma glucose. LDL=low-density lipoprotein cholesterol. SBP=systolic blood pressure. SD=standard deviation.

**Table S35: SHRs using exposures averaged over waves 1 and 3**

SHRs per 1 SD with each exposure averaged over waves 1 and 3 to reduce regression dilution; Models 1–3 as in table S29. The sample is the subset of the 8012 eligible participants who were also stroke-free and reinterviewed at wave 3 with complete wave-3 measurements (n=4284), with follow-up starting at wave 3 and a single follow-up wave; the number entering each model is given in the column heading. Covariates are wave-1 values apart from age, which is age at wave 3, because education and rural residence are recorded only at wave 1; medication use, depression, and marital status are therefore measured about four years before follow-up begins. Exposures were standardised on this sample’s SD, which is 0·83–0·93 times the corresponding main-analysis SD because two-wave means are less variable, so the SHRs are not on the same scale as those in table S29.

**Abbreviations**

BMI=body-mass index. eGFR=estimated glomerular filtration rate. FG=fasting plasma glucose. LDL=low-density lipoprotein cholesterol. SBP=systolic blood pressure. SD=standard deviation. SHR=subdistribution hazard ratio.

**Table S36: Kidney-function association re-estimated with creatinine–cystatin C eGFR (n=6109)**

Distributional comparisons are paired and use all participants with both creatinine and cystatin C measured; model estimates use those among them with complete covariate data. Sample sizes are given with each row. Exposures were standardised on this subsample’s SD, so the SHRs are not on the same scale as those in tables S29 and S33. The age-stratified estimates are mutually adjusted for all five exposures, as in table S33.

**Abbreviations**

CI=confidence interval. eGFR=estimated glomerular filtration rate. SD=standard deviation. SHR=subdistribution hazard ratio.

**Table S37: Instruments**

Instrument counts are before harmonisation with each outcome; harmonised counts are given in table S38. F was computed per variant as (β/SE)², with F≥10 required for retention. Variant selection used p<5×10^−8^, relaxed to p<1×10^−5^ in south Asian ancestry.

**Abbreviations**

BMI=body-mass index. CSA=central and south Asian. eGFR=estimated glomerular filtration rate. FG=fasting plasma glucose. LDL=low-density lipoprotein cholesterol. nSNP=number of single-nucleotide polymorphisms retained as instruments. SBP=systolic blood pressure. SE=standard error. TPMI=Taiwan Precision Medicine Initiative.

**Table S38: Univariable IVW estimates**

Random-effects IVW; OR per genetically predicted unit of exposure, on scales that differ between ancestries. FDR correction is across the five exposures within each outcome. Total stroke is an ancestry-internal auxiliary outcome whose phenotype definition differs by ancestry (table S8) and is interpreted within ancestry only; south Asian ICH and SAH analyses were not possible. In south Asian ancestry the GIGASTROKE any-stroke and ischaemic-stroke datasets differ at only a small number of variants, none of which was retained as an instrument for SBP, LDL, BMI, or eGFR; estimates for those four exposures are consequently identical across the two outcomes, and only FG differs.

**Abbreviations**

BMI=body-mass index. CI=confidence interval. eGFR=estimated glomerular filtration rate. FDR=false discovery rate. FG=fasting plasma glucose. GIGASTROKE is the name of the stroke genome-wide association consortium and is not an initialism. ICD-10=International Classification of Diseases, 10th revision. ICH=intracerebral haemorrhage. IVW=inverse-variance weighted. LDL=low-density lipoprotein cholesterol. nSNP=number of single-nucleotide polymorphisms retained as instruments. OR=odds ratio. SAH=subarachnoid haemorrhage. SBP=systolic blood pressure.

**Table S39: Sensitivity analyses for the principal associations**

Entries were selected by three criteria: the univariable IVW estimate reached FDR-corrected significance; the outcome is a stroke subtype, total stroke being excluded; and, as a single addition, the east Asian LDL–IS comparison discussed in the text although it did not reach significance. No south Asian association qualified and none is shown. Q p is Cochran's Q for between-instrument heterogeneity and the Egger intercept p value tests directional pleiotropy. Full estimates for every exposure–outcome pair in all three ancestries are provided as source data.

**Abbreviations**

BMI=body-mass index. FDR=false discovery rate. ICH=intracerebral haemorrhage. IS=ischaemic stroke. IVW=inverse-variance weighted. LDL=low-density lipoprotein cholesterol. MR=mendelian randomisation. SAH=subarachnoid haemorrhage. SBP=systolic blood pressure.

**Table S40: Multivariable MR**

All five exposures modelled jointly; 141 instruments in European ancestry (138 for total stroke) and 123 in east Asian ancestry. Q and Egger intercept p values in the panel headings are the multivariable statistics for that outcome. † conditional F between 5 and 10, indicating borderline conditional instrument strength; these estimates are interpreted alongside their univariable counterparts. South Asian multivariable analysis was attempted but conditional F was below 5 for four of five exposures, and those estimates are provided as source data without interpretation. MVMR-Egger estimates, used as a sensitivity analysis and not FDR-corrected, are in the source data.

**Abbreviations**

BMI=body-mass index. CI=confidence interval. eGFR=estimated glomerular filtration rate. FDR=false discovery rate. FG=fasting plasma glucose. LDL=low-density lipoprotein cholesterol. MR=mendelian randomisation. MVMR=multivariable mendelian randomisation. OR=odds ratio. SBP=systolic blood pressure.
