## Supplementary figures and images for "Triangulating population, individual, and genetic evidence for priority setting in structurally heterogeneous blocs: metabolic risk factors for stroke in the Shanghai Cooperation Organisation"

### figure S1

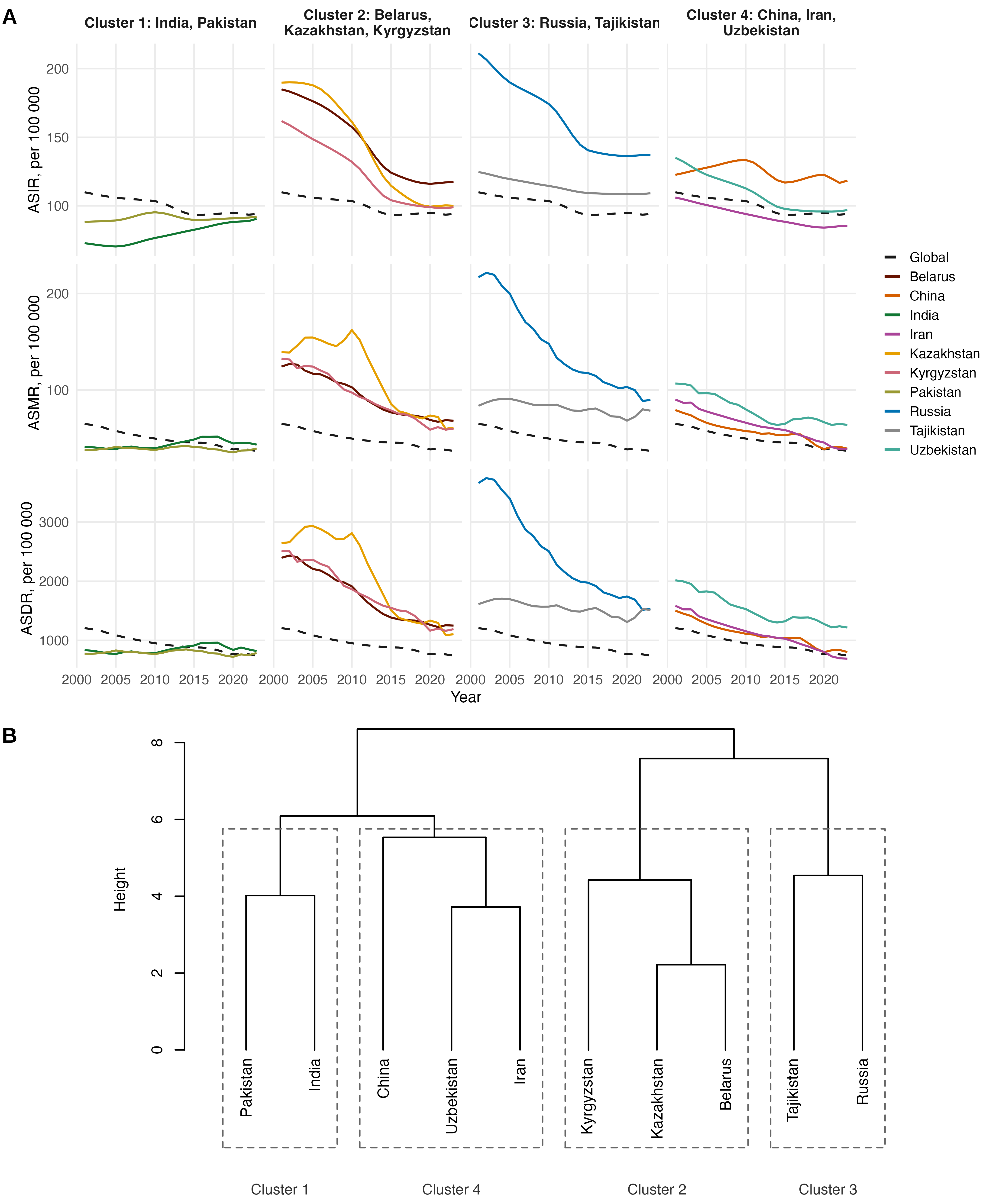

### figure S2

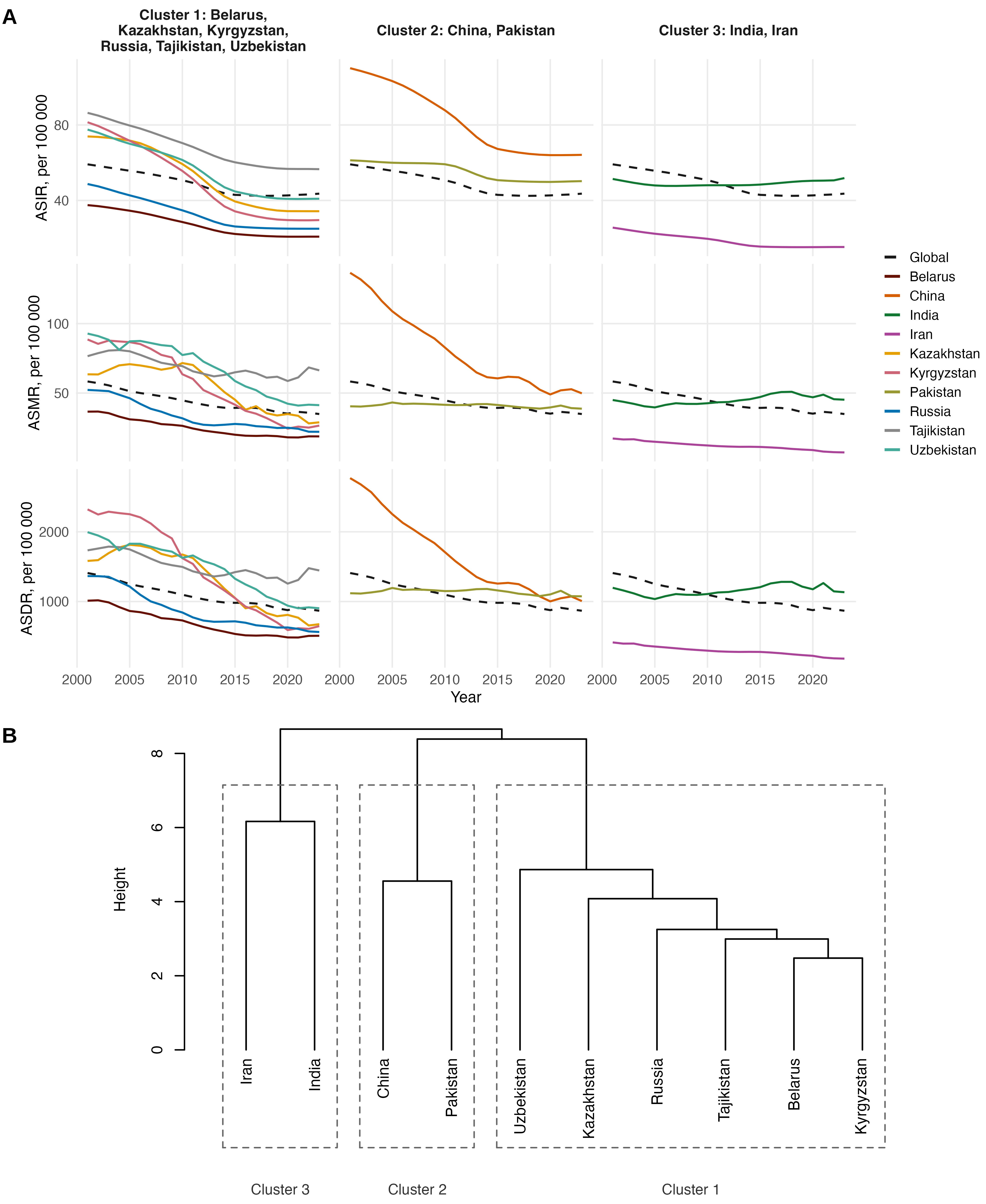

### figure S3

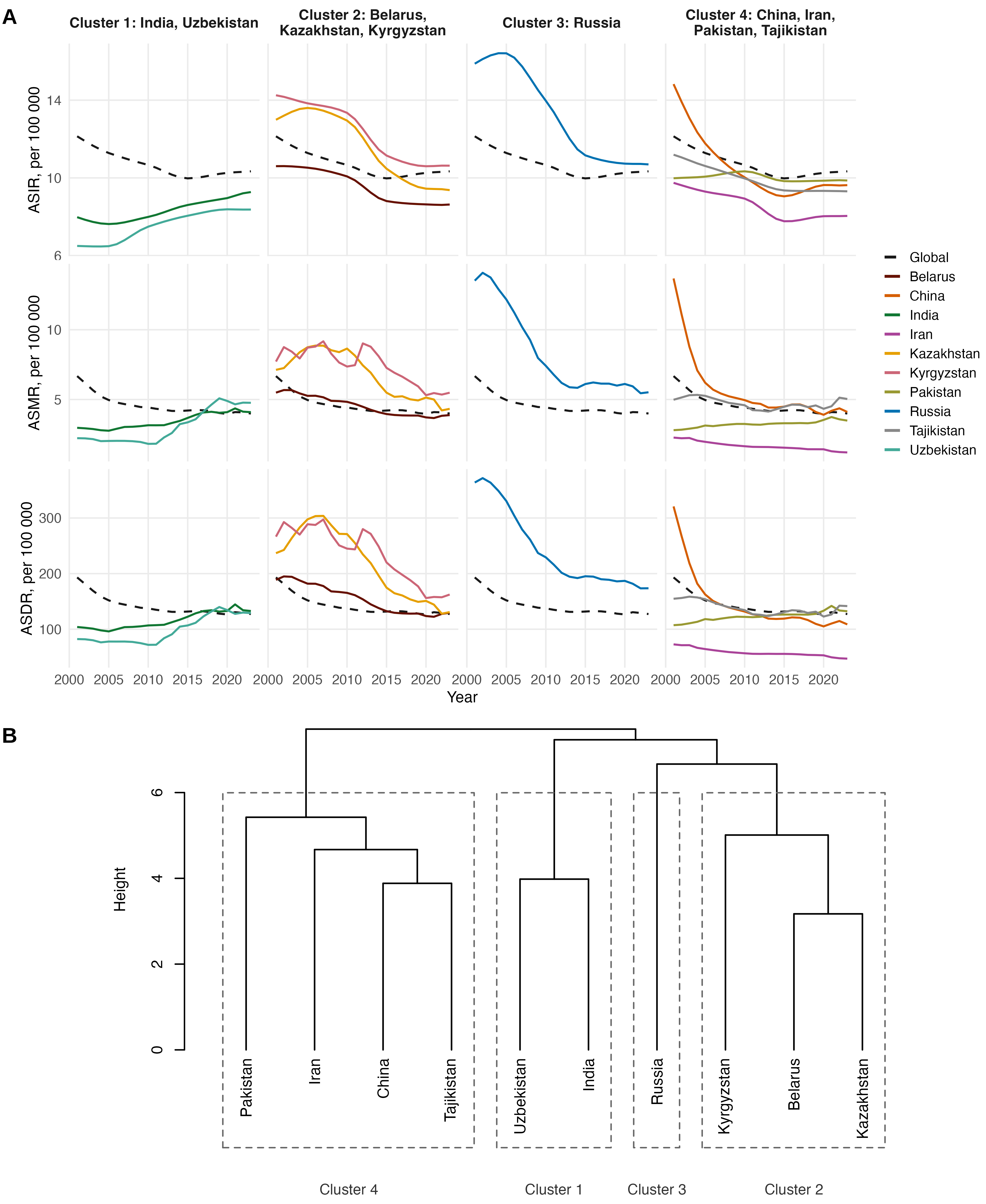

### figure S4

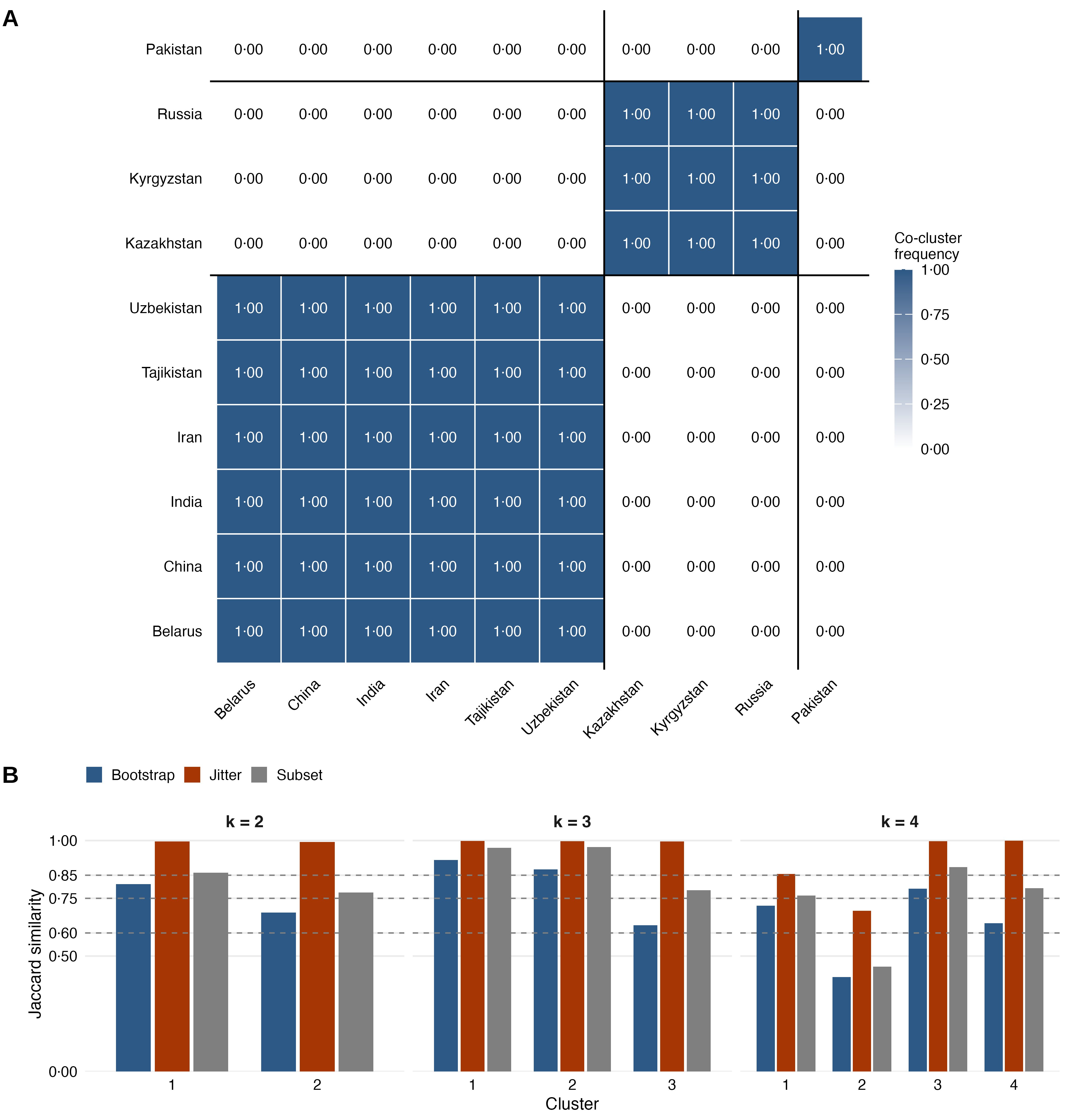

### figure S5

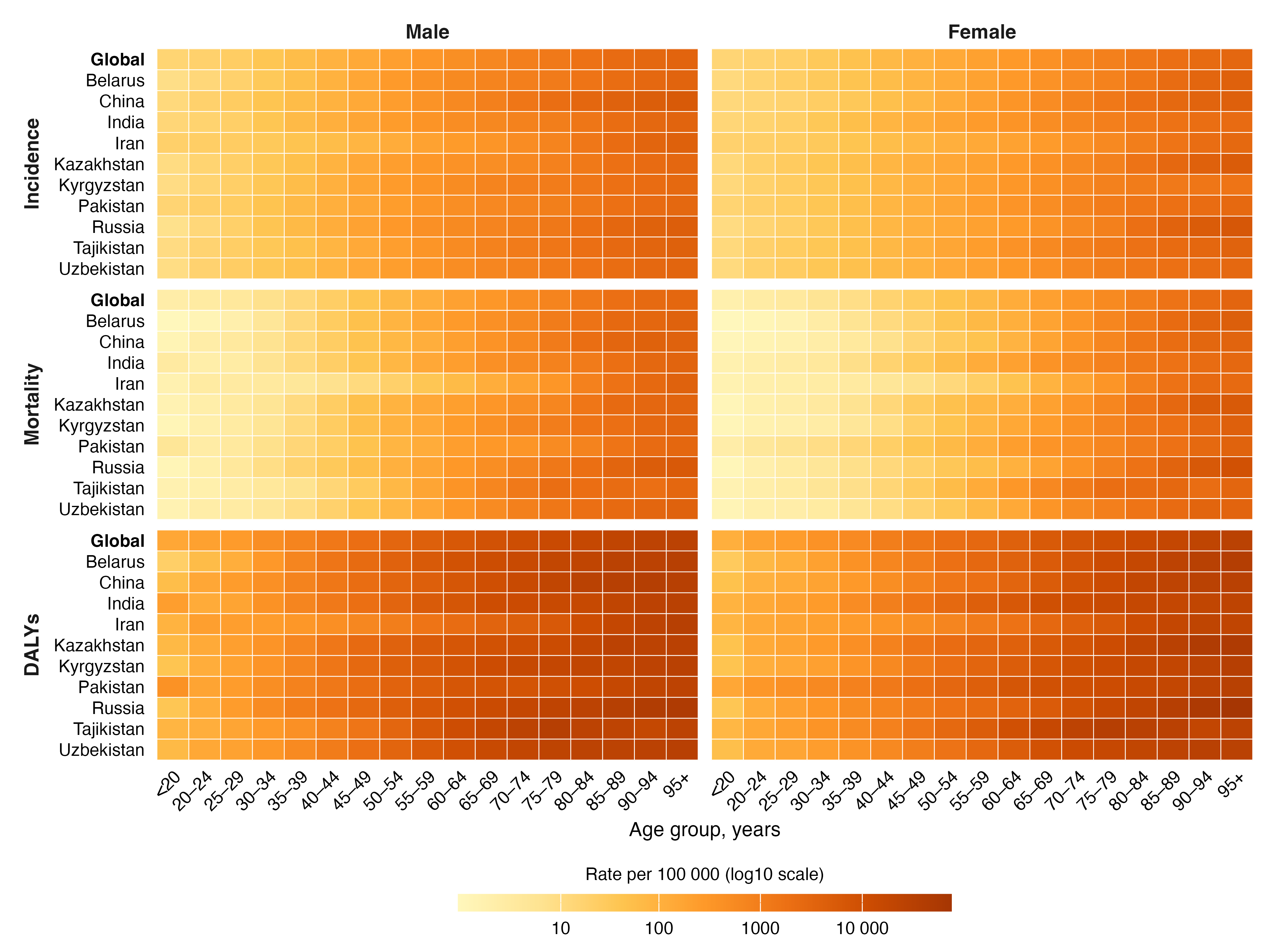

### figure S6

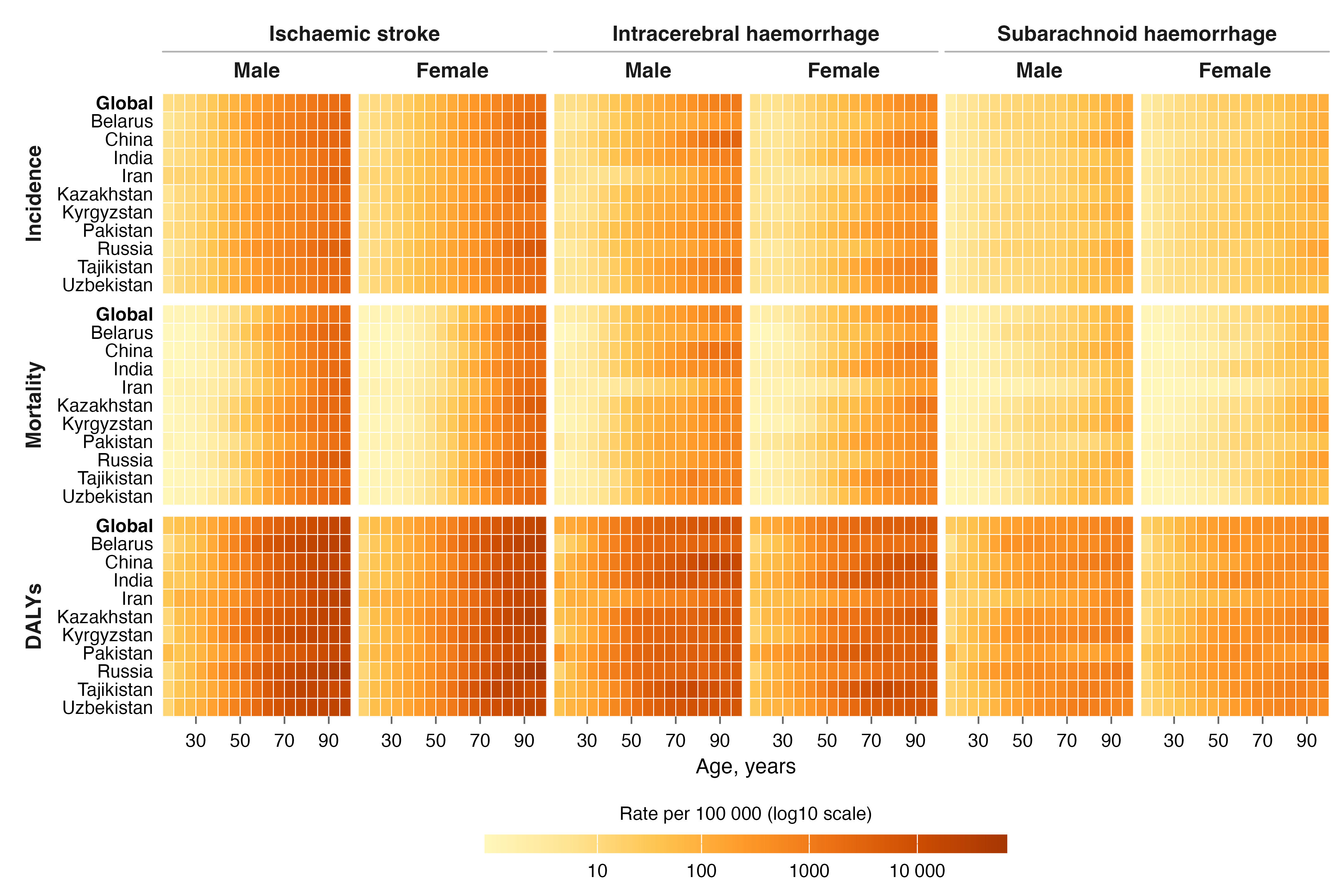

### figure S7

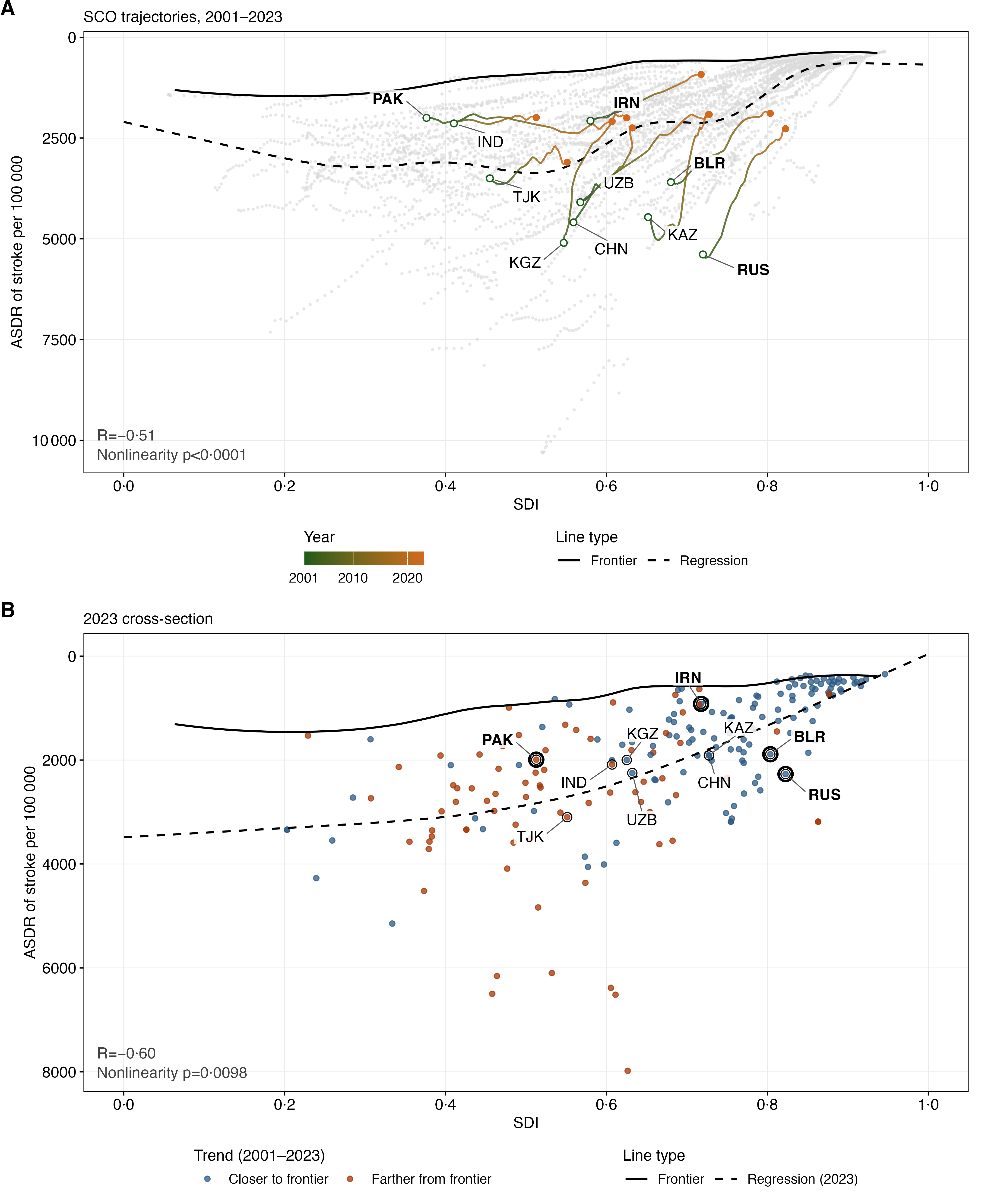

### figure S8

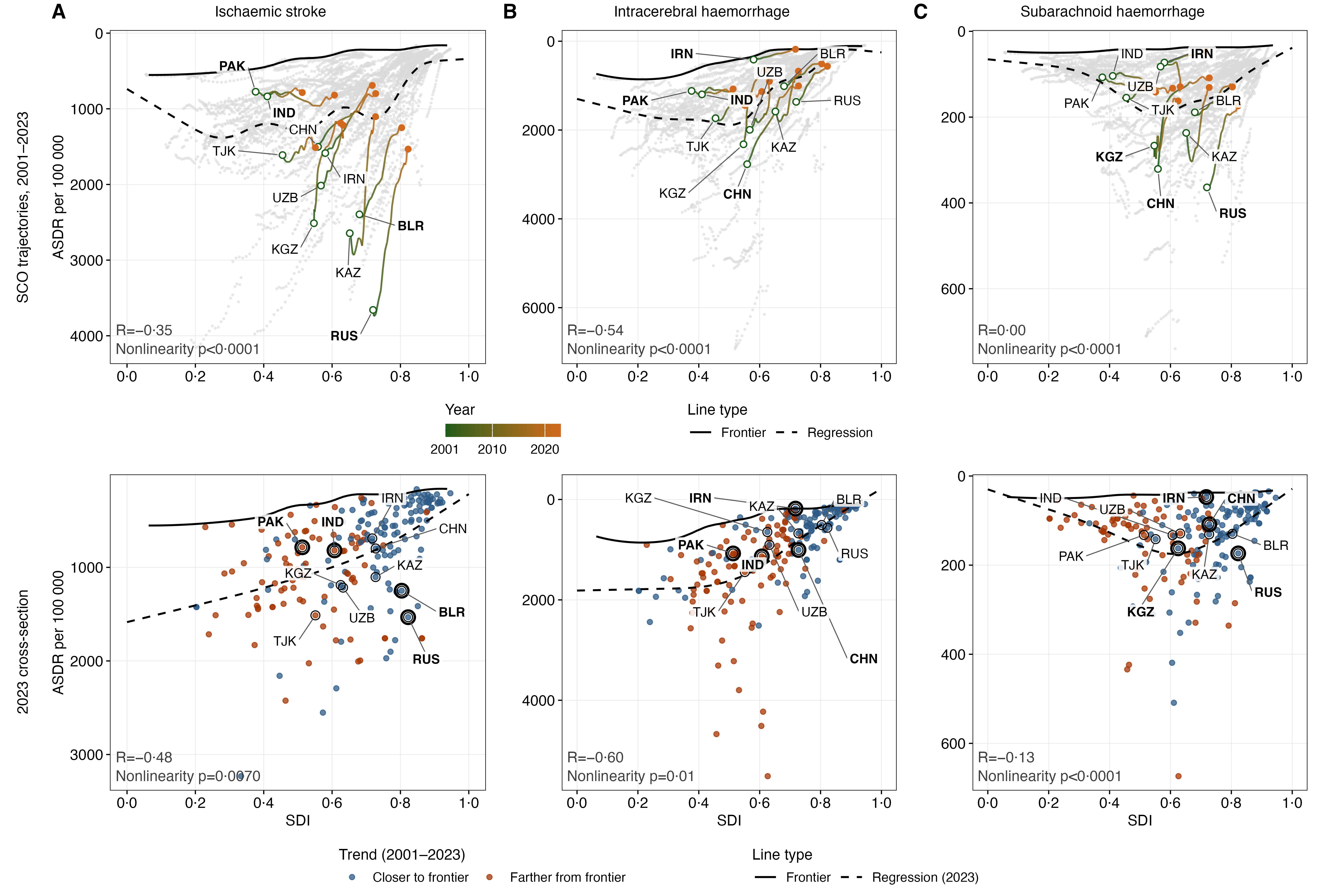

### figure S9

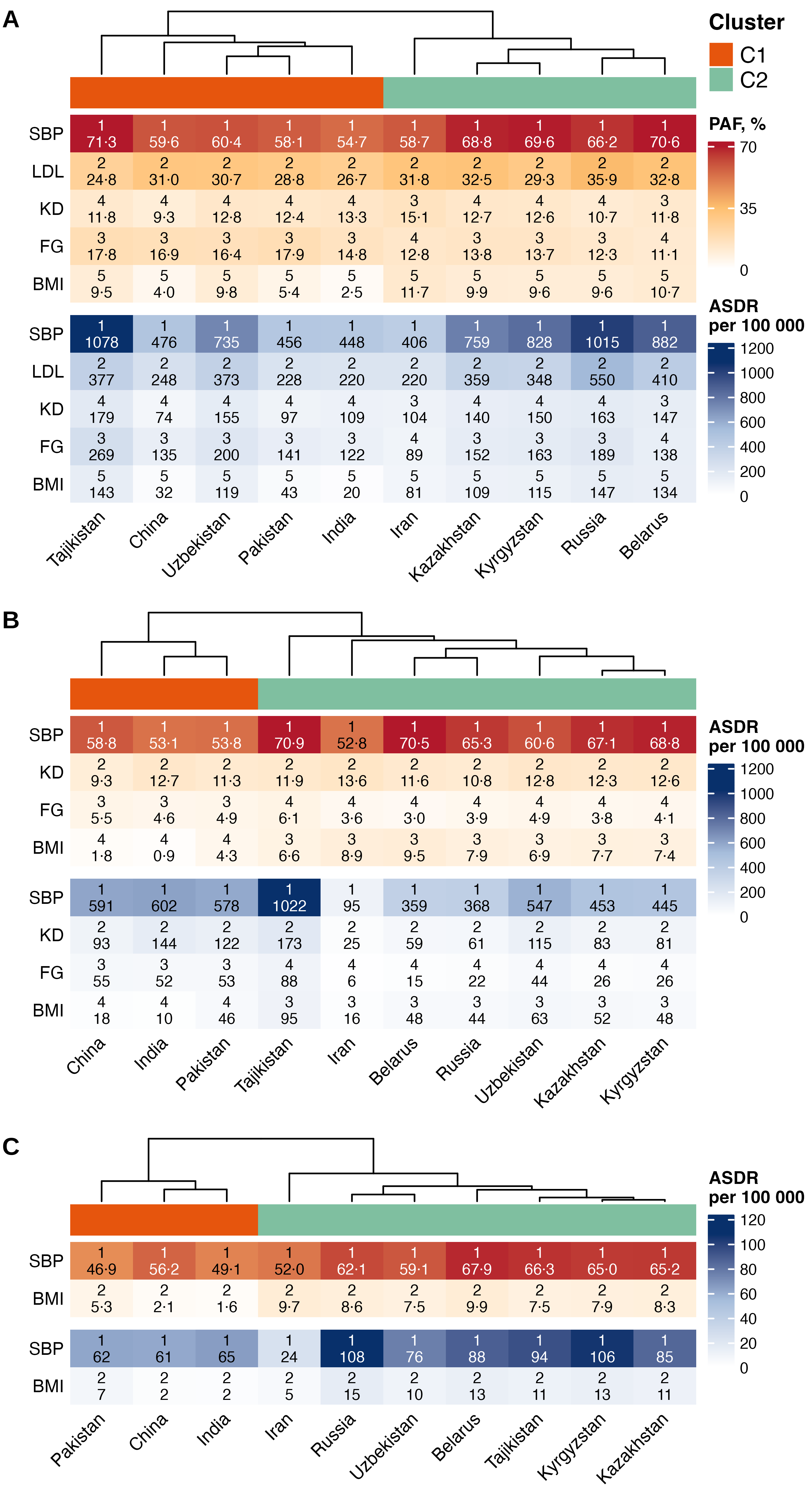

### figure S10

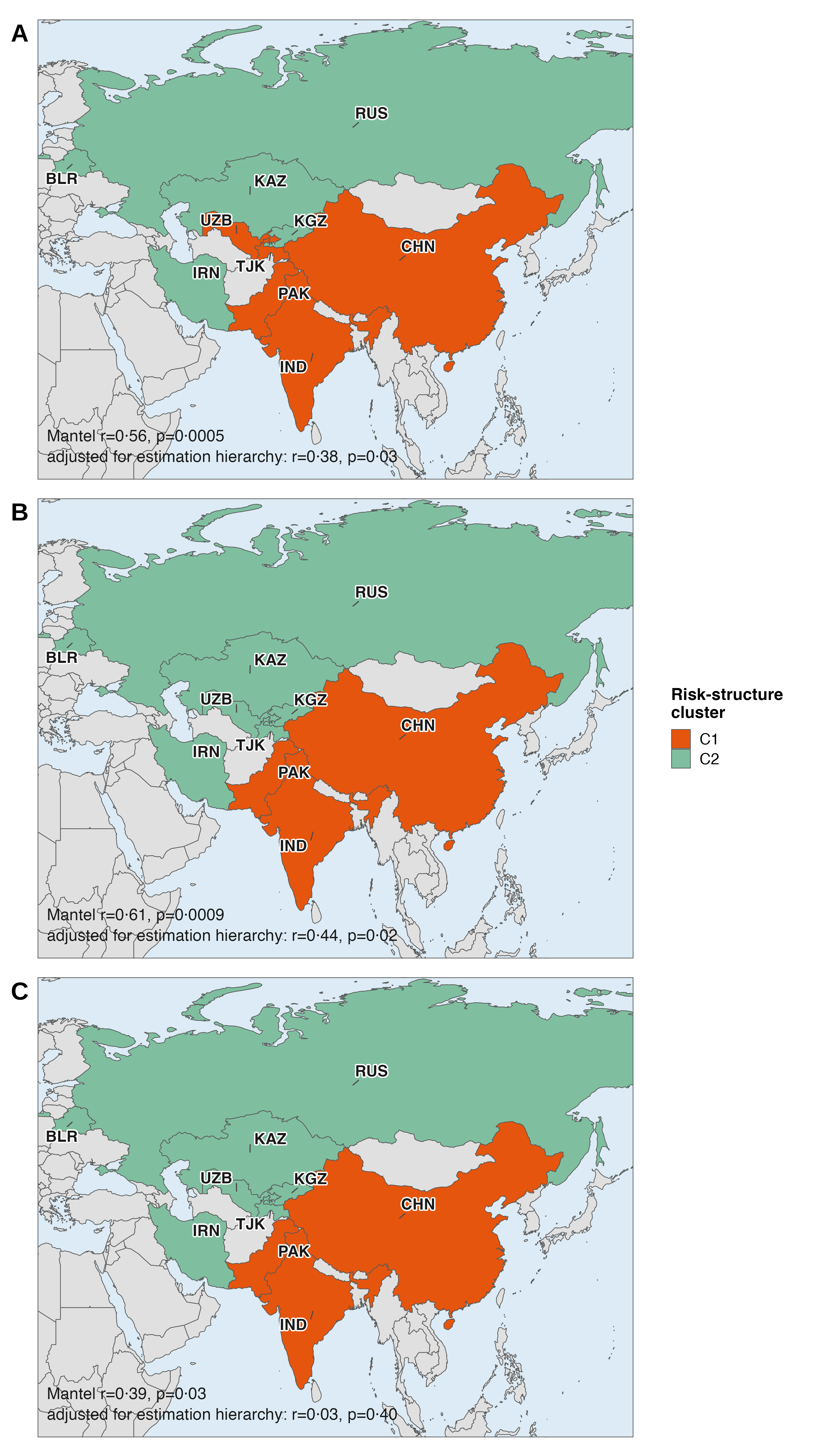

### figure S11

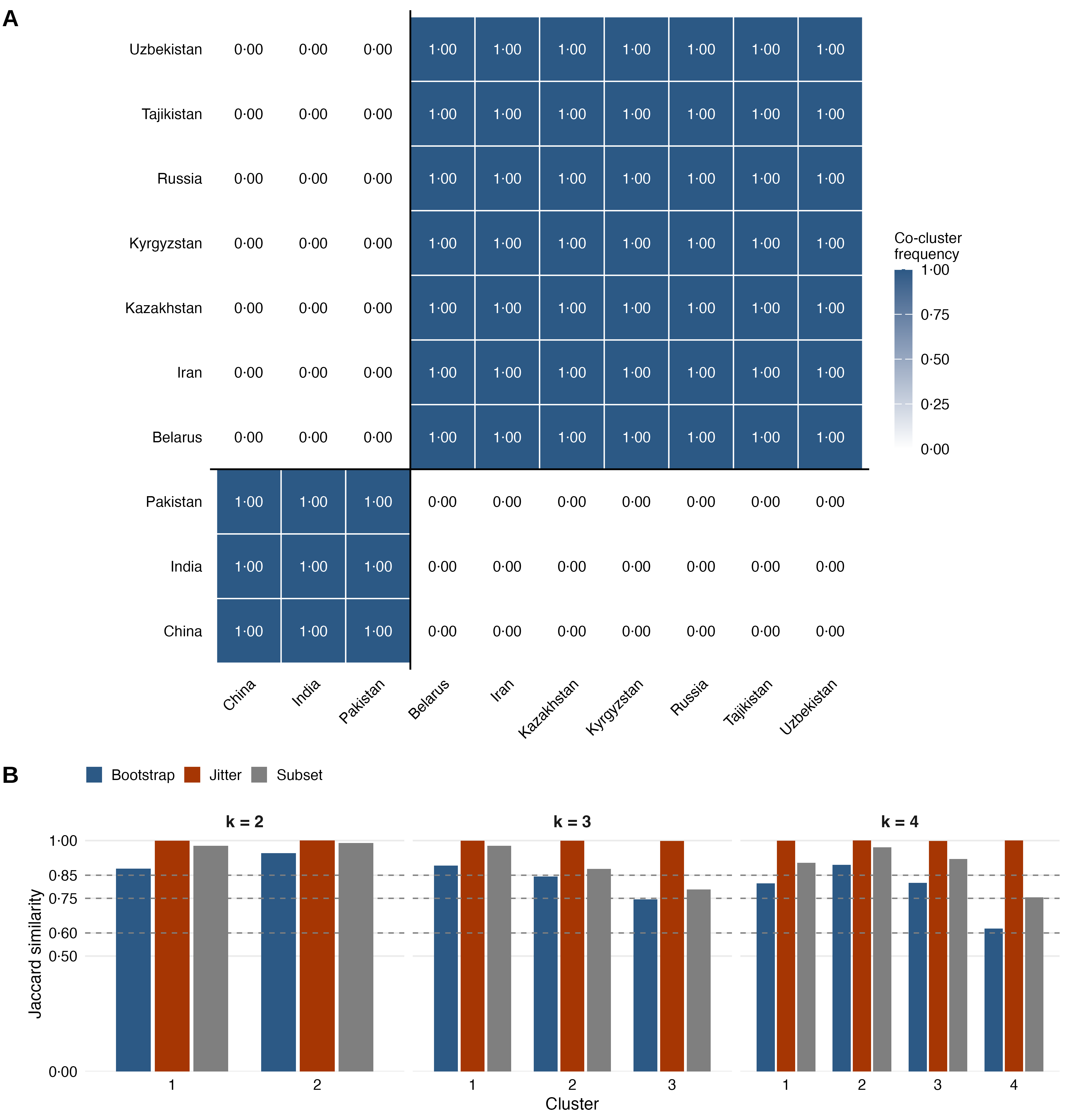

### figure S12

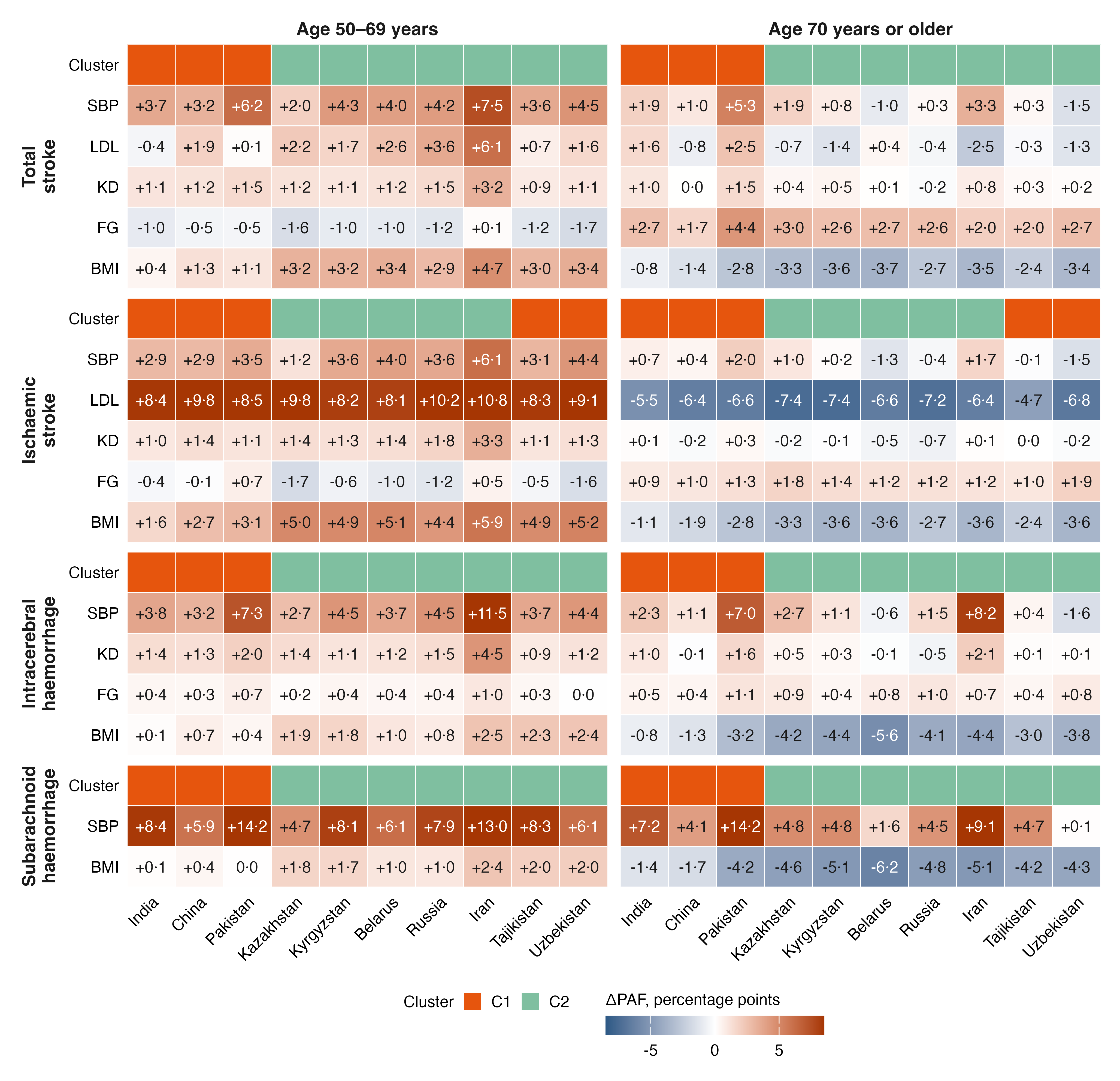

### figure S13

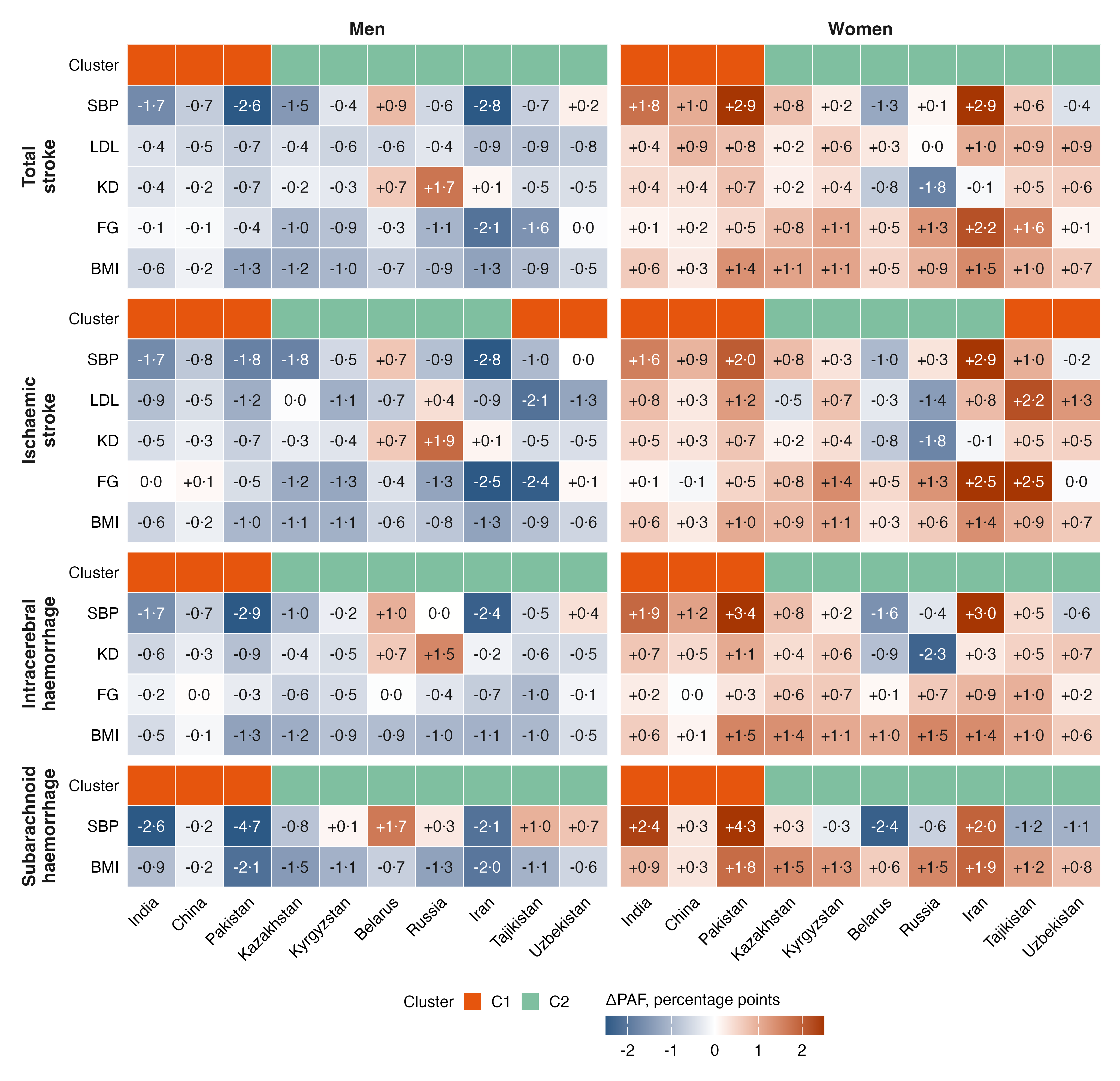

### figure S14

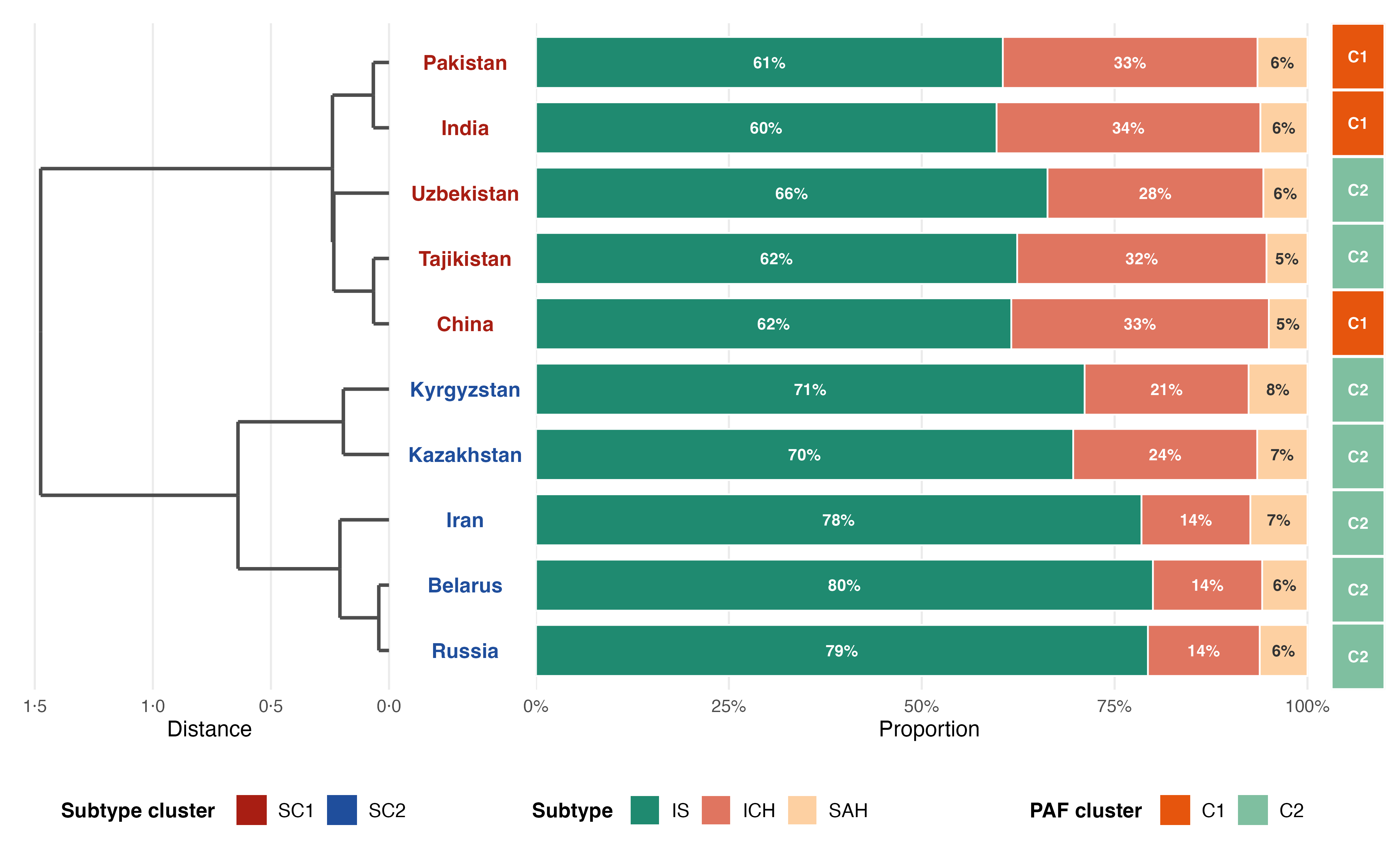

### figure S15

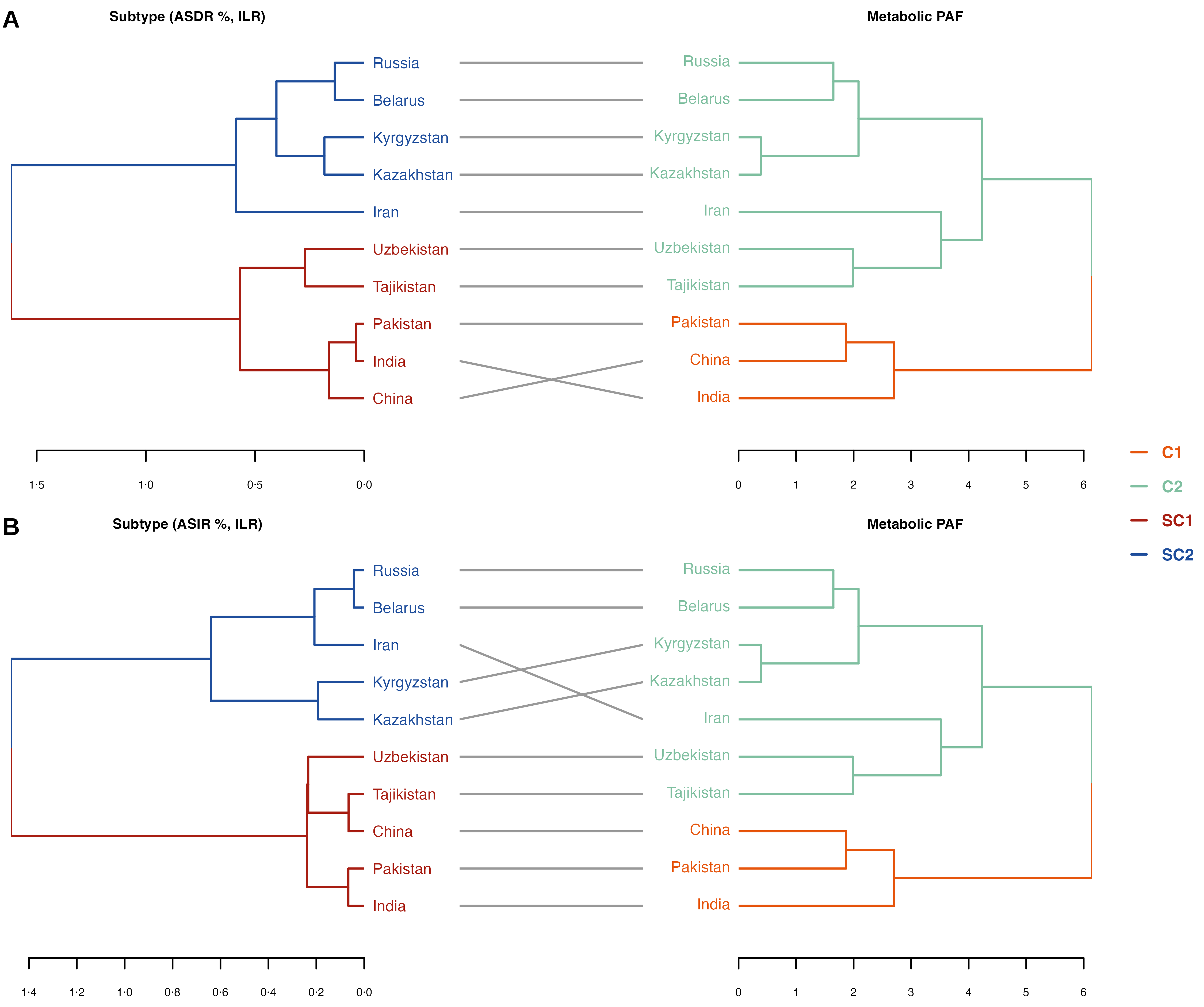

### figure S16

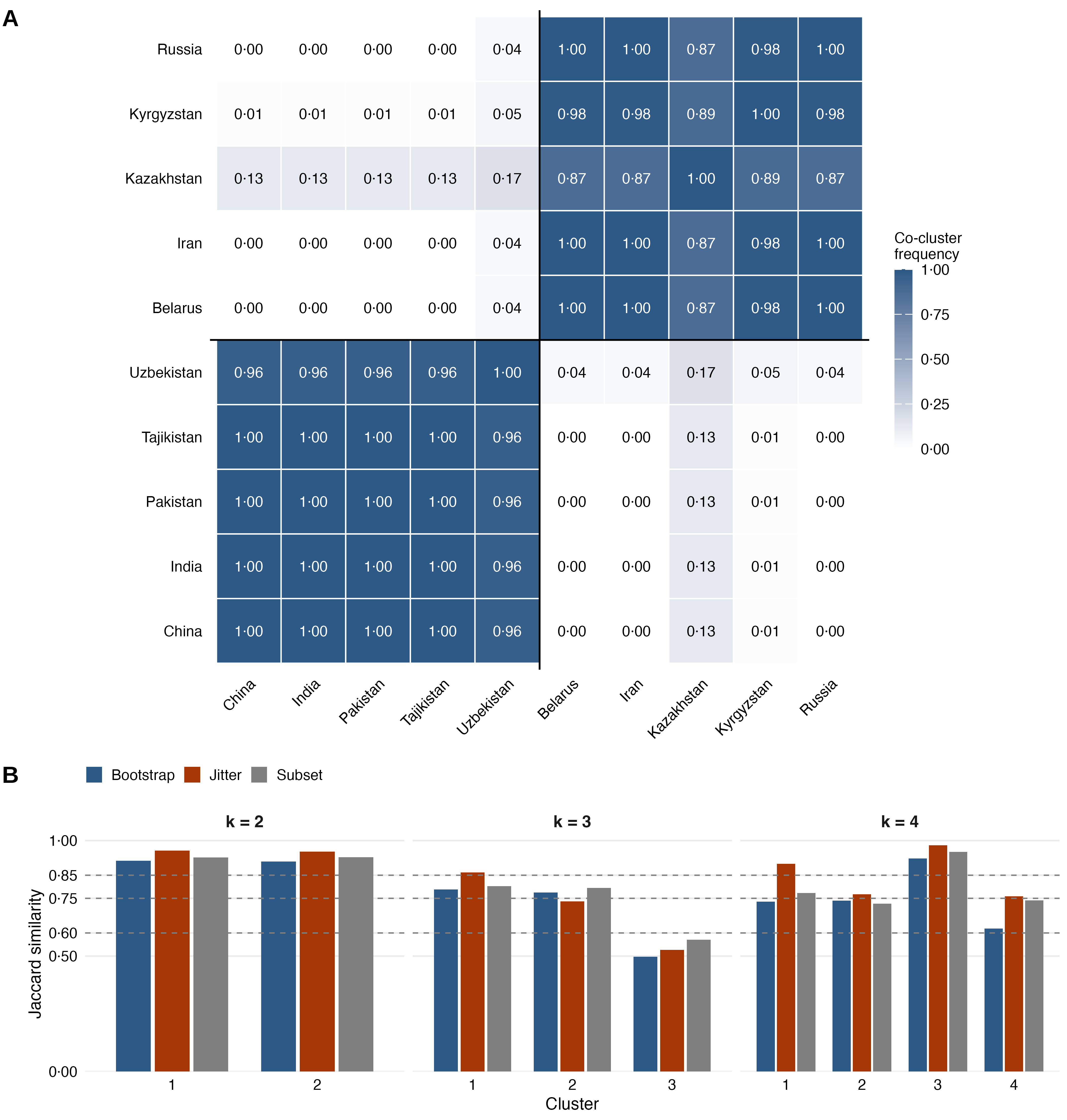

### figure S17

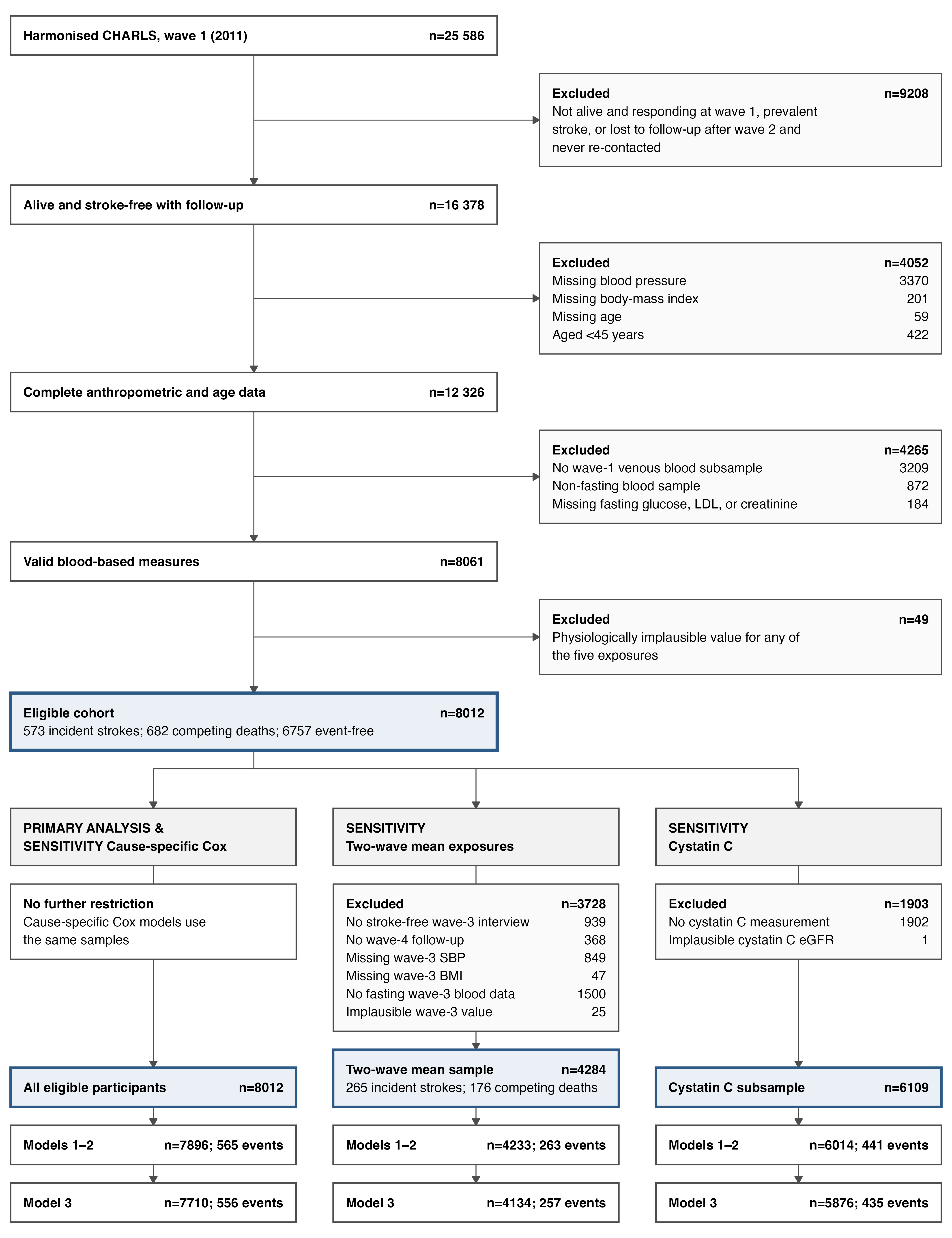

### figure S18

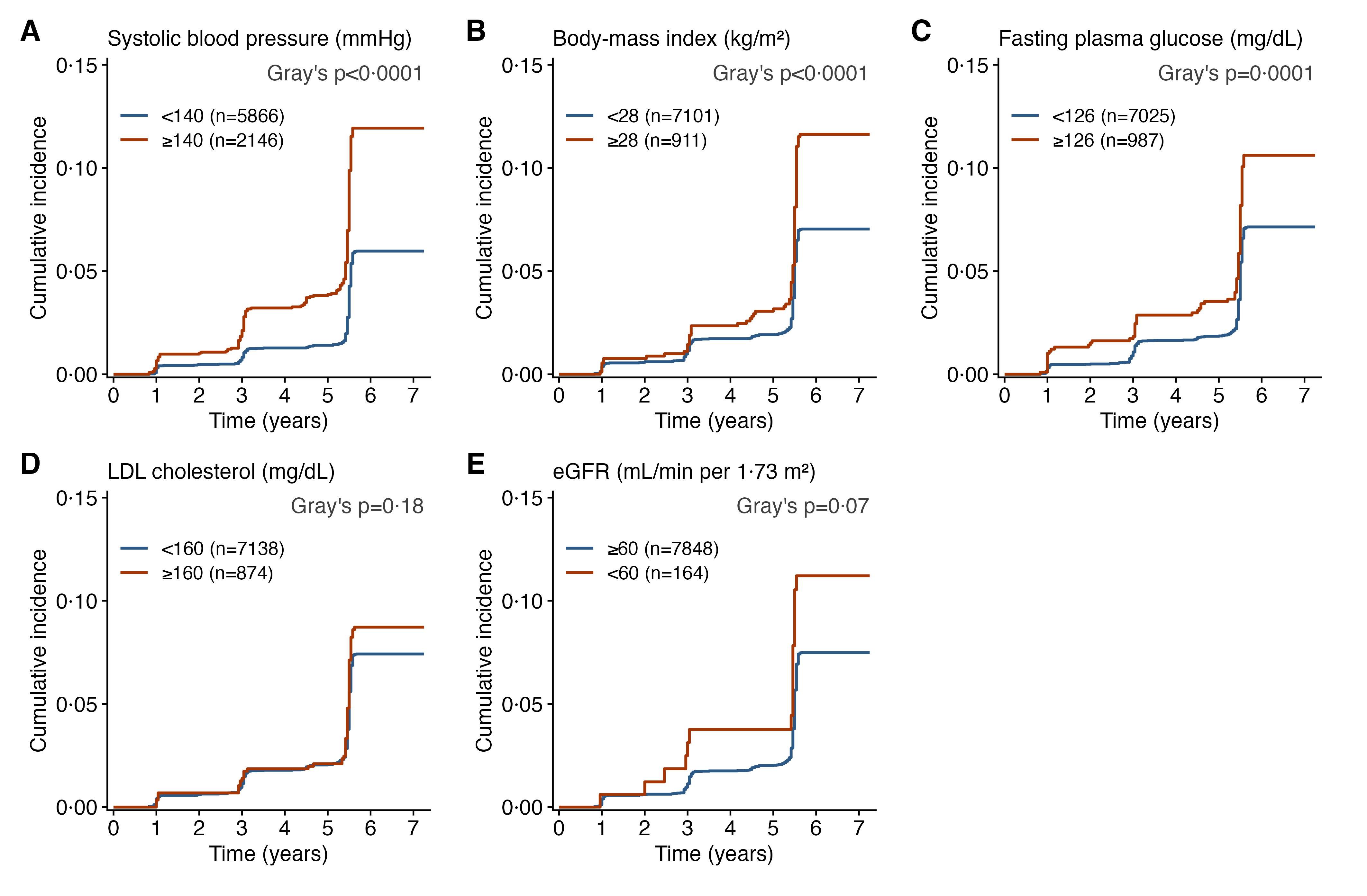

### figure S19

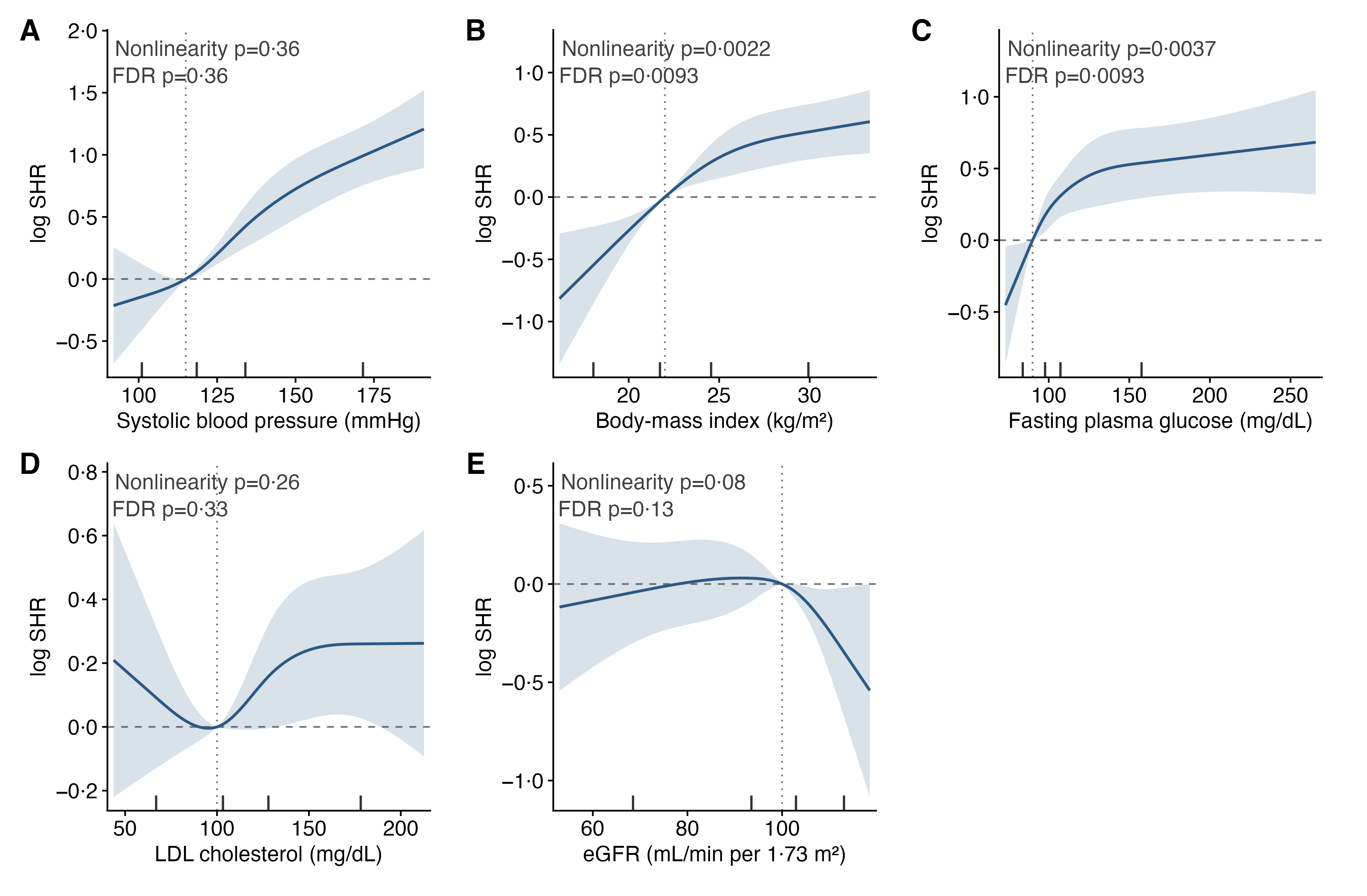

### figure S20

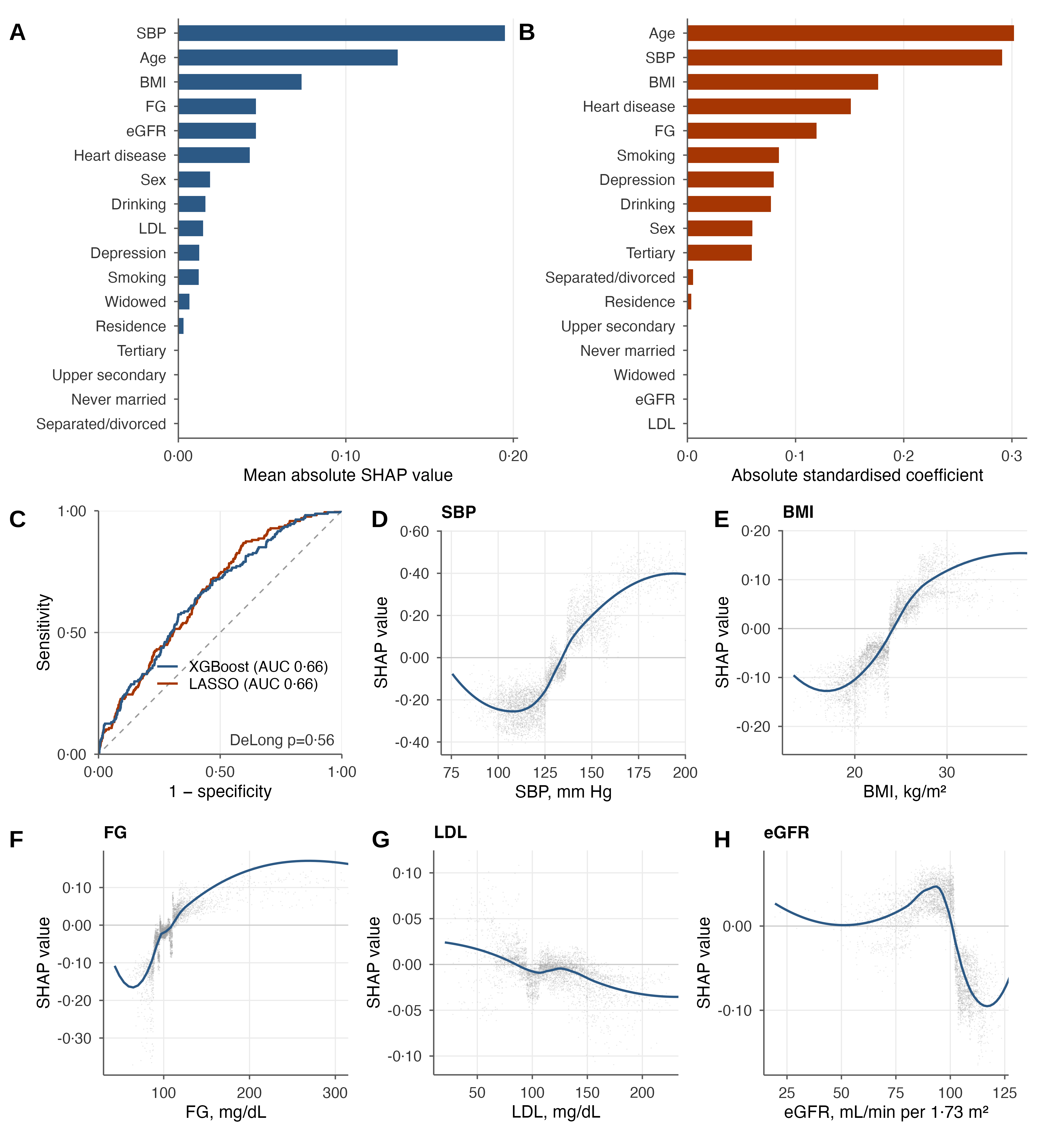
